# Linking the NETSARC+ national sarcoma database with the SNDS to evaluate adjuvant and/or neoadjuvant therapy: report on the linkage process and result (Health Data Hub’s DEEPSARC pilot project)

**DOI:** 10.1101/2025.05.02.25326859

**Authors:** Erwan Drezen, André Happe, Vincent Thevenet, Nicolas Penel, François Gouin, François Le Loarer, Gonzague Du Bouexic De Pinieux, Hugo Crochet, Claire Chemin Airiau, Françoise Ducimetiere, Simone Mathoulin Pelissier, Jean-Yves Blay, Emmanuel Oger

## Abstract

**Background:** DEEPSARC, one of the first project running on the Health Data Hub aimed to identify real-life treatment regimens that could improve overall survival. The project is based on matching the national database of the sarcoma reference network with the SNDS.

**Objectives:** We aimed to report a transparent description of the linking process and its results.

**Methods:** The sarcoma database encompasses 33,548 patients matching the selection criteria divided in three subsets: 13,507 patients with a complete dataset gathering clinical and pathological data; 5,844 patients with clinical data alone; and 14,197 patients with pathological data alone. As no ICD-10 code reliably identifies patients with sarcoma the subpopulation extracted from the SNDS was extended to 3 million patients who underwent surgery for their cancer. An indirect record linkage process used a combination (called a signature) of so-called chaining variables to uniquely identify a pair of patients from each of the bases. Two metrics (signature robustness and overall quality) were calculated for ease of interpretation.

**Results:** The overall matching rate of 73.1% (24,539 pairs out of 33,548 observations), reaching 90.5% in the intersection of the sarcomas databases (with extended data, 12,225 pairs out of 13,507 observations).

**Conclusion:** Detailed reporting, along with dedicated metrics, contribute to the transparency of the process, as discussion and interpretation of the chaining results are crucial for the validity of the main results of the study.

## Introduction

Although clinical data sources (clinical trials, epidemiological cohorts or registries), provide valuable insights, even the best of them are limited by factors such as participant bias, attrition, and data inconsistency. These clinical data sources make it possible to collect the most relevant data for the field of research under consideration. For sarcomas, the multiplicity of histological types and subtypes, difficult to classify, leading to diagnostic errors ^1^ and a centralized double reading corrects up to 30% of diagnoses with a major impact on the choice of treatment. ^2,3^ The national database of the sarcoma reference network (Sarcoma-BCB) has recorded around 90% of patients with sarcoma, since 2010, with centralized review in the 25 expert centers of the NETSARC+ network. Even for a network like NETSARC+, collecting follow-up data for all patients is an enormous challenge. Despite significant resources, with more than a third of patients not followed up in reference centers, the survey could not be completed for almost 20% of patients.^4^

The use of a medico-administrative database such as the SNDS, “*Système National des Données de Santé*” (French National Health Data System),^5^ a database derived from medical care reimbursement data, allows, through its universality and the exhaustiveness of the data collection necessary for its primary purpose (reimbursing the insured and participating in the financing of healthcare establishments), tracking without attrition, as long as the subject remains within the health insurance fold. Other sources of data, such as civil status records, also provide information on the vital status of individual citizens.

Other highly relevant data, however, would require an unthinkable effort to collect on a large scale, and especially over the long term: all the information needed to describe a patient’s course of treatment (consultations, treatments, hospitalizations). A medico-administrative database is therefore ideal. On the other hand, data collected for medico-administrative purposes have limitations (little or no clinical data, no categorization of sarcoma extension at diagnosis, no histological data). Sarcomas are coded, in the context of hospital stays, by location with organ cancers, which does not enable them to be identified easily, correctly or exhaustively. Linking clinical data with data already collected systematically for other purposes, such as medico-administrative data, can overcome data gaps in clinical studies and enable comprehensive analysis of patient care pathways and outcomes.^6^

DEEPSARC was one of the first project running on the Health Data Hub (HDH). The aim of the study was to improve the management of sarcoma patients by identifying real-life treatment regimens that improve overall survival. The project proposed a nationwide analysis based on matching the national database of the sarcoma reference network (Sarcoma-BCB) with data from the SNDS, enabling to understand patients care pathways and healthcare consumption over 5 years of follow-up. The primary objective was to estimate whether the administration of adjuvant and/or neoadjuvant therapy for non-metastatic sarcomas is associated with a better overall survival.

One of the challenging aspects of the DEEPSARC project was to start mid 2019 in a regulatory and technical context under construction. Established by the Law of 24 July 2019 aiming the organization and transformation of the French healthcare system, the HDH becomes a public entity that provides a single gateway to assist project leaders with their administrative procedures and a secure, state-of-the-art platform with advanced data storage, computing and analysis capabilities.

We aimed to report a transparent description of the linking process realized on this platform and its results.

## Material and Methods

### Data collection

On one hand, the Sarcoma-BCB database allows to describe the population of sarcoma patients in France since 2010. It includes a set of data describing patients and tumor characteristics, surgery, relapse and survival by cross comparison of the clinical database (NETSARC) and of the pathological review database for soft tissues and viscera sarcoma (RREPS) and for bone sarcoma (RESOS). A complete description of the 3 data dictionaries of these 3 subsets of the database can be found here: https://netsarc.sarcomabcb.org/public/dictionary/display for NETSARC; https://rreps.sarcomabcb.org/public/dictionary/display for RREPS; https://resos.sarcomabcb.org/public/dictionary/display for RESOS.

The selection criteria for the DEEPSARC project in the sarcoma database were a diagnosis of sarcoma according to the 2013 WHO classification of connective tissue tumors, established between 01/01/2010 and 31/12/2017 and surgery of the primary tumour. As the 3 subsets of the database were not structured to meet the same initial objectives, discrepancies in the data may appear if the patient exists in two of them. These discrepancies were dealt with (using algorithms) by a data manager when linking patients from two subsets in order to prepare the database. It is also important to note that the pathology double-reading could have been performed on the biopsy. In this case, only biopsy-related data are collected in the database while surgery-related data are not and the surgery procedure is not mentioned . As a result, some patients were selected without knowledge whether they underwent a surgery or not (and some of them were subsequently excluded during the record linkage process when the surgery procedure was not found in the SNDS data).

The completeness of the data in the Sarcoma-BCB database is not uniform for all patients and varies depending on the patient’s access to double reading pathology organization and discussion in sarcoma multidisciplinary team meetings (MDTM). The 33,548 patients are divided in three subsets according to the availability of diagnostic and/or clinical data in the database: 13,507 patients with a complete dataset gathering clinical and pathological data which represent 40,3 % of the whole population; 5,844 patients with clinical data alone which represent 17,4 % of the whole population; 14,197 patients with pathological data alone which represent 42,3 % of the whole population (this latter proportion is significant because it was impossible to select only patients who had undergone surgery). By convention this database will be called the « source «; the clinical part of it will be called NETSARC while the pathological part of it will be called RREPS (RREPS+RESOS).

On the other hand, the SNDS is built on individual data, linked by pseudonymized record identification from 3 databases: the SNIIRAM containing health insurance data derived from the processing of healthcare reimbursements, mainly for ambulatory care; the PMSI containing data derived from the healthcare activity delivered in private and public establishments; and the CepiDC database containing data on causes of death derived from the treatment of certificates of death. The SNDS covers around 99% of the French population (which means more than 67 million people) since 2006 and it will allow a follow-up over a 20-year sliding window. A comprehensive description of the SNDS database dictionary can be found here https://health-data-hub.shinyapps.io/dico-snds/.

It has been shown that using ICD-10 code was not a reliable method to identify patients diagnosed with sarcoma due to a lack of consistency in ICD coding for the diagnosis limiting the ability to conduct real-world observational research of this rare disease.^7^ For this reason, the criteria for extracting the subpopulation of the SNDS have been extended to patients who underwent surgery for their cancer between 2010 and 2018, a detailed description is provided in Supplementary Material/section A. The CNIL expressed concerns about the size of the subpopulation to be extracted and recommended that the matching step and analysis phase would be carried out in separate environments in order to reduce the risk to expose a 3 million patients’ population on the new HDH technological platform. By convention this subset of the SNDS will be called the « target ».

### Principles of the record linkage process

In order to achieve the objectives of the DEEPSARC project, an initial step of data linkage was needed between the « source » and the « target » databases to enrich the Sarcoma-BCB database with the complete care pathway of the patients. Data or record linkage has been defined as “a process of pairing records from two files and trying to select the pairs that belong to the same entity.^8^” In our case, as no common unique patient identifier was available, an indirect record linkage process was used retaining several non-directly identifying variables eventually at hand in the « source » and the « target » databases.^9^

The underlying principle is that a combination of such variables (called chaining variables) makes it possible to uniquely identify a patient in each of the bases, such a combination of chaining variables is called a signature (S). However, each chaining variable does not hold the same discriminatory power since for example, a sex code generally separates all patients into two categories, whereas a year of birth will divide them into groups of smaller sizes.

The algorithm can be described using the set theory. For each patient P in the « source » base having a given signature composed of N chaining variables (for example sex, birthdate, date of a cancer surgery, town of residency), it searches among all the patients in the « target » base for the one who has the same signature, or at least the closest subpart of the signature. If a unique patient holding the source signature (or a part of it) can be found in the target, a linked pair has been constituted.

As the algorithm explore the whole research space (i.e. the cartesian product between patients from the source and target databases), a quality index can be estimated called the robustness (R). The robustness R of a linked pair is the number of chaining variables that can be dropped from the signature without losing the uniqueness of the linked pair. For example, a robustness R=2 indicates that we could remove any of 2 variables from the signature without losing the unicity of the link. On the other hand, a robustness R=0 means that there is no possibility to remove any chaining variable from the signature otherwise the uniqueness of the link would be lost.

### Data preparation for record linkage

#### Step 1: Application of the chaining variables

In order to prepare the chaining process, a selection of discriminating chaining variables is carried out within the « source » and « target » database. When it is possible the chaining variables are associated with a date in order to make them more discriminating, for example a variable C « Type of Tumor », whose modalities are « Soft tissue », « Bone », « Viscera » will be associated with the date of surgery. Furthermore, a margin of error allowed on the dates can be specified in order to resist to discrepancies in the process of collecting data in both systems. For example, a variable M « Micro-biopsy » which represents the presence or absence of a sampling of tumor in the care pathway of the patient could be associated with the date of the sampling itself in one system and with the date of the production of the result of the tissue fragments in the other database. For the death date there is no such a variable in « source » database, only a date of last contact that can be combined with a vital status (dead or alive) which means that there could be a variable delay between this combination of variables in the « source » and the real death date in the « target ». For the date of surgery, the « source » database collects the exact date while in the « target » this information is linked to the beginning of the hospital stay, and therefore the date of surgery could be potentially posterior to it.

This preparation led to a « linkage management book » describing the way each chaining variable entering in the signature is calculated (see Table 1 in Supplementary Material/section B). The details of the code used is in Supplementary Material/section B.

**Table 1.**
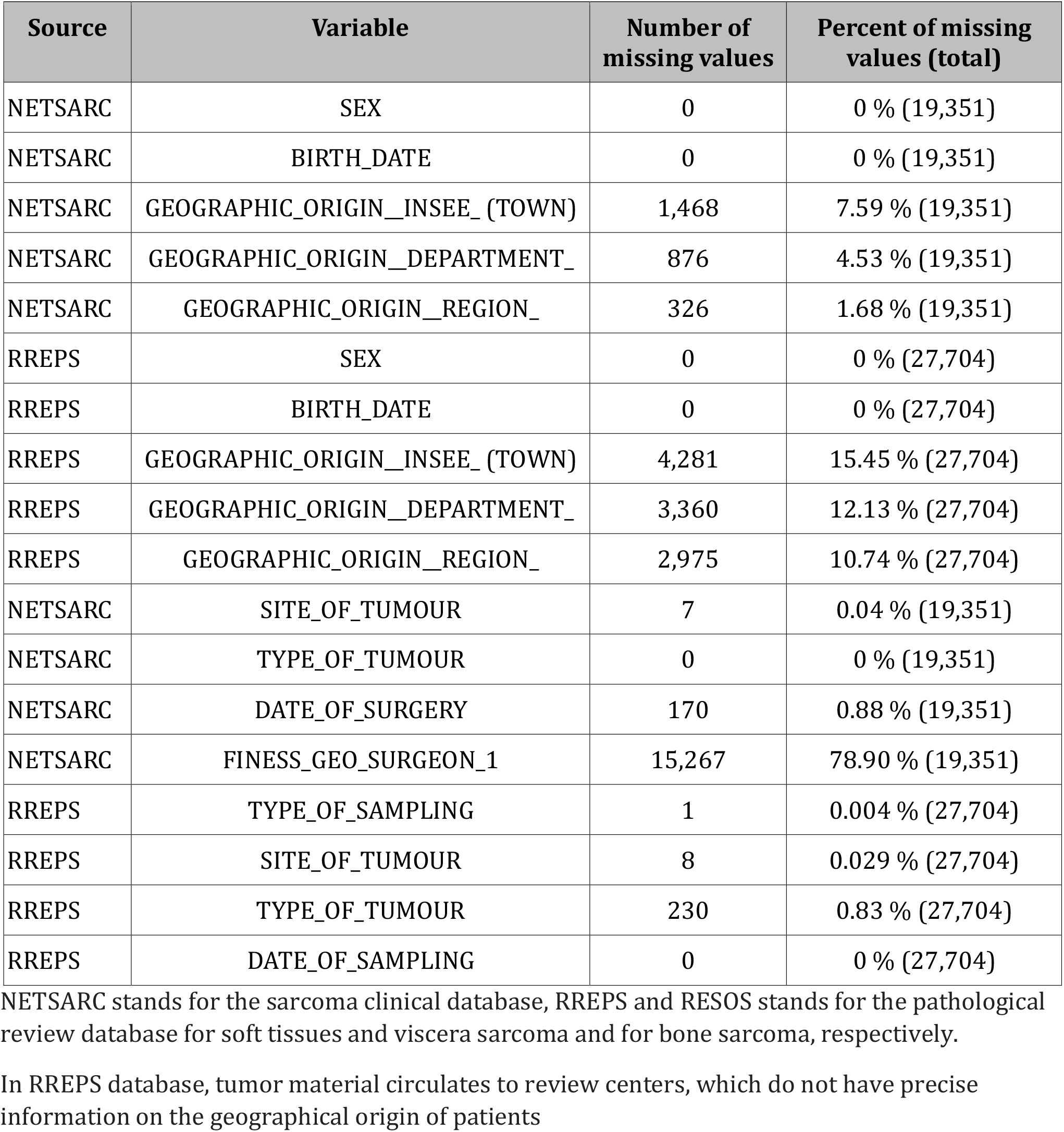
Rates of missing values for the expected chaining variables in the source database

| Source | Variable | Number of missing values | Percent of missing values (total) |
| --- | --- | --- | --- |
| NETSARC | SEX | 0 | 0 % (19,351) |
| NETSARC | BIRTH_DATE | 0 | 0 % (19,351) |
| NETSARC | GEOGRAPHIC_ORIGIN_INSEE_ (TOWN) | 1,468 | 7.59 % (19,351) |
| NETSARC | GEOGRAPHIC_ORIGIN_DEPARTMENT_ | 876 | 4.53 % (19,351) |
| NETSARC | GEOGRAPHIC_ORIGIN_REGION_ | 326 | 1.68 % (19,351) |
| RREPS | SEX | 0 | 0 % (27,704) |
| RREPS | BIRTH_DATE | 0 | 0 % (27,704) |
| RREPS | GEOGRAPHIC_ORIGIN_INSEE_ (TOWN) | 4,281 | 15.45 % (27,704) |
| RREPS | GEOGRAPHIC_ORIGIN_DEPARTMENT_ | 3,360 | 12.13 % (27,704) |
| RREPS | GEOGRAPHIC_ORIGIN_REGION_ | 2,975 | 10.74 % (27,704) |
| NETSARC | SITE_OF_TUMOUR | 7 | 0.04 % (19,351) |
| NETSARC | TYPE_OF_TUMOUR | 0 | 0 % (19,351) |
| NETSARC | DATE_OF_SURGERY | 170 | 0.88 % (19,351) |
| NETSARC | FINESS_GEO_SURGEON_1 | 15,267 | 78.90 % (19,351) |
| RREPS | TYPE_OF_SAMPLING | 1 | 0.004 % (27,704) |
| RREPS | SITE_OF_TUMOUR | 8 | 0.029 % (27,704) |
| RREPS | TYPE_OF_TUMOUR | 230 | 0.83 % (27,704) |
| RREPS | DATE_OF_SAMPLING | 0 | 0 % (27,704) |

NETSARC stands for the sarcoma clinical database, RREPS and RESOS stands for the pathological review database for soft tissues and viscera sarcoma and for bone sarcoma, respectively.
In RREPS database, tumor material circulates to review centers, which do not have precise information on the geographical origin of patients

At this stage the chaining process can be launched in order to produce, ideally, a unique linked pair for each patient of the « source » database. In practice, 3 situations result from the launch of the chaining algorithm: (1) no candidate can be found in the « target » database for a given patient in the « source » database; (2) a unique candidate has been found in the « target » database for a given patient in the « source » database; (3) two or more candidates have been found in the « target » database for a given patient in the « source » database. The matching rate will be calculated by dividing the final number of unique pairs by the total number of patients in the « source » database.

#### Step 2: Extension to the checking variables

If several candidates are eligible for a given patient in the « source » database, one could try to improve the matching rate by integrating more information into the chaining process in order to remove ambiguity among the various candidates in the « target » database. This disambiguation process relies on so-called checking variables. For example, the NETSARC database collects decisions taken in multidisciplinary team meeting, mainly indications of appropriate treatment options for the patient like chemotherapy, radiotherapy or tumor re-excision. Knowing this planned treatment in the near future could help to choose between 2 candidates by searching such an event in their care pathway.

Another opportunity is to use diagnosis recorded during the surgery stay, even though ICD-10 codes where not considered sufficiently reliable at first to identify sarcoma.^7^ A limited mapping table between 2,000 ICD-10 codes and a combination of information about the type (Soft tissue, Bone, Viscera) and the anatomical site of the tumor was produced in order to separate the good from the bad when 2 ambiguous candidates where found after step1.

The details of the code are in Supplementary Material/section C. Table 2 in Supplementary Material/section C presents the checking variables used in this step.

**Table 2.**
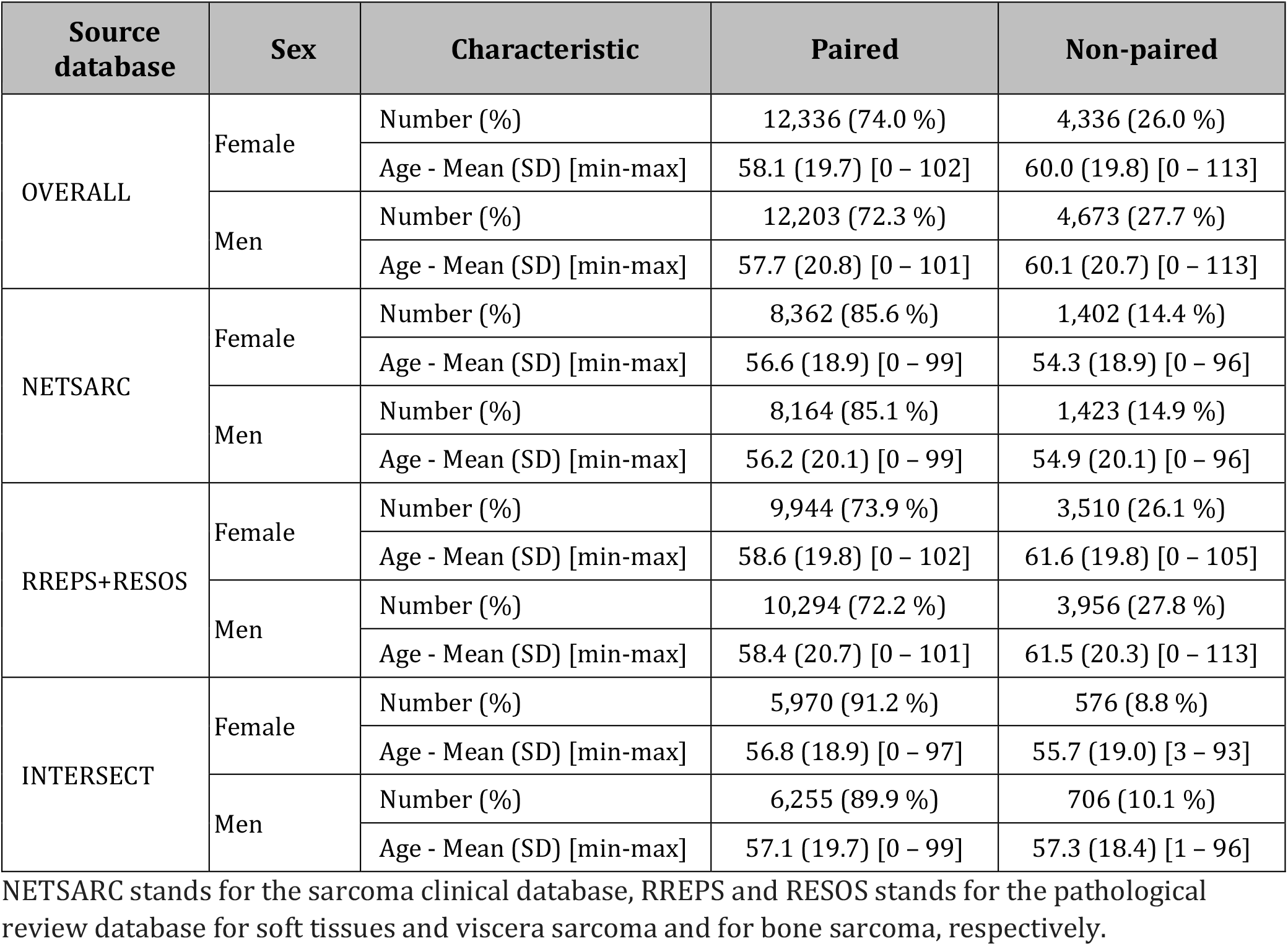
Characteristics of the paired and non-paired patients by origin of the « source » database

| Source database | Sex | Characteristic | Paired | Non-paired |
| --- | --- | --- | --- | --- |
| OVERALL | Female | Number (%) | 12,336 (74.0 %) | 4,336 (26.0 %) |
|  |  | Age - Mean (SD) [min-max] | 58.1 (19.7) [0 – 102] | 60.0 (19.8) [0 – 113] |
|  | Men | Number (%) | 12,203 (72.3 %) | 4,673 (27.7 %) |
|  |  | Age - Mean (SD) [min-max] | 57.7 (20.8) [0 – 101] | 60.1 (20.7) [0 – 113] |
| NETSARC | Female | Number (%) | 8,362 (85.6 %) | 1,402 (14.4 %) |
|  |  | Age - Mean (SD) [min-max] | 56.6 (18.9) [0 – 99] | 54.3 (18.9) [0 – 96] |
|  | Men | Number (%) | 8,164 (85.1 %) | 1,423 (14.9 %) |
|  |  | Age - Mean (SD) [min-max] | 56.2 (20.1) [0 – 99] | 54.9 (20.1) [0 – 96] |
| RREPS+RESOS | Female | Number (%) | 9,944 (73.9 %) | 3,510 (26.1 %) |
|  |  | Age - Mean (SD) [min-max] | 58.6 (19.8) [0 – 102] | 61.6 (19.8) [0 – 105] |
|  | Men | Number (%) | 10,294 (72.2 %) | 3,956 (27.8 %) |
|  |  | Age - Mean (SD) [min-max] | 58.4 (20.7) [0 – 101] | 61.5 (20.3) [0 – 113] |
| INTERSECT | Female | Number (%) | 5,970 (91.2 %) | 576 (8.8 %) |
|  |  | Age - Mean (SD) [min-max] | 56.8 (18.9) [0 – 97] | 55.7 (19.0) [3 – 93] |
|  | Men | Number (%) | 6,255 (89.9 %) | 706 (10.1 %) |
|  |  | Age - Mean (SD) [min-max] | 57.1 (19.7) [0 – 99] | 57.3 (18.4) [1 – 96] |

NETSARC stands for the sarcoma clinical database, RREPS and RESOS stands for the pathological review database for soft tissues and viscera sarcoma and for bone sarcoma, respectively.

However, those variables could not be used directly as chaining variables since they would have brought too many false positives in the first step of the chaining process, either because the indication for treatment is not necessarily followed by the therapeutic procedure initially planned or because the margin of error on dates on such events could be high or just because the effort to produce a wider mapping table was out of reach.

Finally, a rule of thumb was defined to determine, *a priori*, which of the patients in the « source » database with several candidates in the « target » database will be verified with those checking variables. Two criteria are applied in order to limit this step to patients with a strong concordance with their candidates: (1) the signature of the patient at the end of step 1 must contain at least 5 variables, and (2) the signature of the patient at the end of step 1 must have no more than 2 missing variables.

The step 2 is launched for each patient meeting the criteria in the « source » database, the checking variables are compared for each candidate in the « target » database filtered in the step 1. If a candidate shows at least one checking variables matching with the « source » patient and if no other candidate does then the pair is considered as potentially unique at the end of step 2 and improves the matching rate.

#### Step 3: Elimination of questionable pairs

At this stage, all the operations of the algorithm were unsupervised. As stated earlier, sarcoma need a skillful clinician expertise and the pairing process could not escape the rule. A checking of several care pathways was carried out in order to identify signatures of unique pairs that could seem doubtful to clinicians, the aim was to identify a general rule of safety that automatically eliminates questionable pairs, even if it means accepting the loss of real positives. The criteria were applied to eliminate complete groups of patients with signatures having the following characteristics: the signature does not contain the 2 chaining variables S and s (complete lack of demographic information on sex and/or birth date in the « target database); the signature does not contain the 2 chaining variables L and l (complete lack of demographic information on localization in the « target database); the signature does not contain one out of the 2 chaining variables S or s, is missing at least 2 chaining variables and contains less than 6 chaining variables; the signature does not contain the chaining variable L, is missing at least 2 chaining variables and contains less than 8 chaining variables; the signature does not contain one out of the 2 chaining variables L or l, is missing at least 2 chaining variables and contains less than 6 chaining variables. The definitive matching rate is calculated at the end of the step 3.

### Definition of a quality metric

However, while robustness can be used to compare individuals within the same signature group, it is less relevant for comparing patients belonging to different signature classes. A metric, called quality (Q), analyze the results globally, overcoming intra-class variability and enable inter-class comparisons of signatures. This metric was defined iteratively as follows: all chained patients at the end of the step 3 are initially assigned a quality of Q = 0; Q = 1 if pairs present a robustness R = 0 with a signature of 4 or less than 4 chaining variables; Q = 2 if pairs present a robustness R = 0 with a signature of more than 4 chaining variables; Q = 3 if pairs present a robustness R = 1 with a signature of 5 or less than 5 chaining variables; Q = 4 if pairs present a robustness R = 1 with a signature of more than 5 chaining variables; Q = 5 if pairs present a robustness R >= 2. In addition, for pairs presenting a quality Q ≥ 2: Q = Q+1 if at least 2 checking variables from step 2 (d, e, c, r) are present; Q = Q +2 if at least 3 checking variables from step 2 (d, e, c, r) are present.

## Results

### Evaluation of missing data in the chaining variables

One of the first results produced was to check the quality of the information and in particular the rate of missing data for the expected contributing variables in the pairing process. Table 1 presents the rate of missing values for those variables in the source database either in the clinical (NETSARC) or pathological subsets (RREPS+RESOS). The demographic variables (sex and birth date) were completely recorded in both subsets since there were needed for creating the hash ID of the patient. However, the geographic origin of the patient was less well recorded in general than for demographic variables with more than 15% of missing data at the most precise level (town of residency) for the pathological base and 7.59% for the clinical base. The clinical base presented a better filling rate at the different levels, the pathological base alone did not allow to locate at all the patient in more than 10% of cases even at the regional level. Concerning the variables related to surgery, 170 patients did not have a date of surgery, which is surprising since this was an inclusion criterion in the study. We observed also 230 patients with a missing value for the variable « type of tumor » which is also surprising since this variable is mandatory and controls the entry of dependent variables in the eCRF. This absence immediately led to eliminate those patients from any possibility of chaining them to the « target » base. Identification of the surgeon was missing in more than 78% of cases due to the fact that the surgeon’s name has only been collected in the sarcoma database since 2016, and only for surgeons in the network. This information can therefore only make a weak contribution to the chaining process and could be at best be an element of disambiguation of equivocal pairs.

### Results of the pairing algorithm

It should be remembered that the pairing algorithm produced 3 types of results: (1) in the most favorable situation a unique pair was found between a patient from the source and the target database; (2) in the less favorable situation, there was no combination of chaining variable that could produce a candidate in the target database for a given source patient; (3) in intermediate situations, it could be found a signature that produces several candidates in the target database for a given source patient.

Figure 1 presents the flowchart of the different steps of the pairing algorithm. Figure 2 gives the definitive matching rates after the 3 steps of the pairing algorithm according to the origin of the « source » database (NETSARC for clinical data or RREPS+RESOS for pathological data).

**Figure 1.**
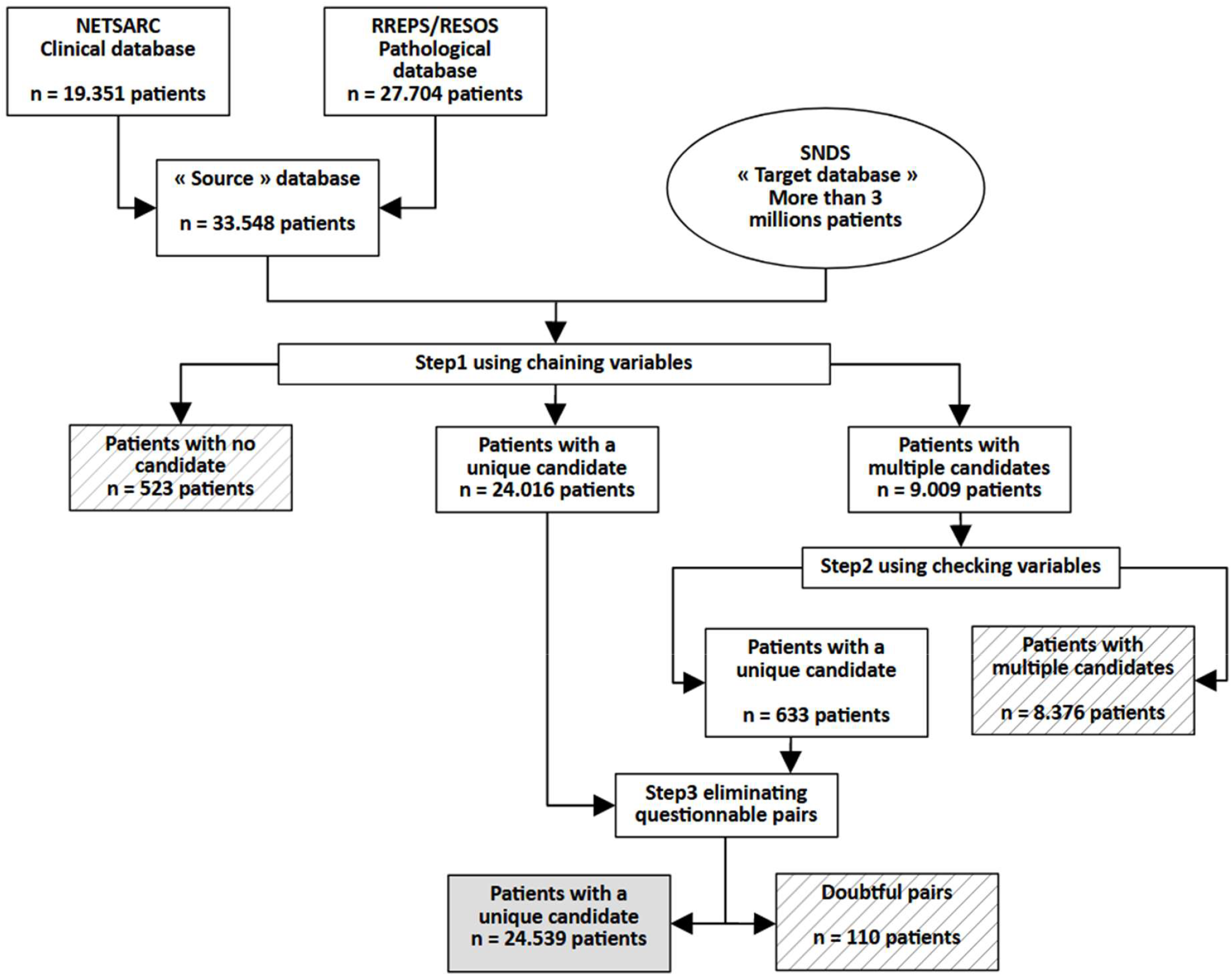
Flowchart of the different steps of the pairing algorithm

**Figure 2.**
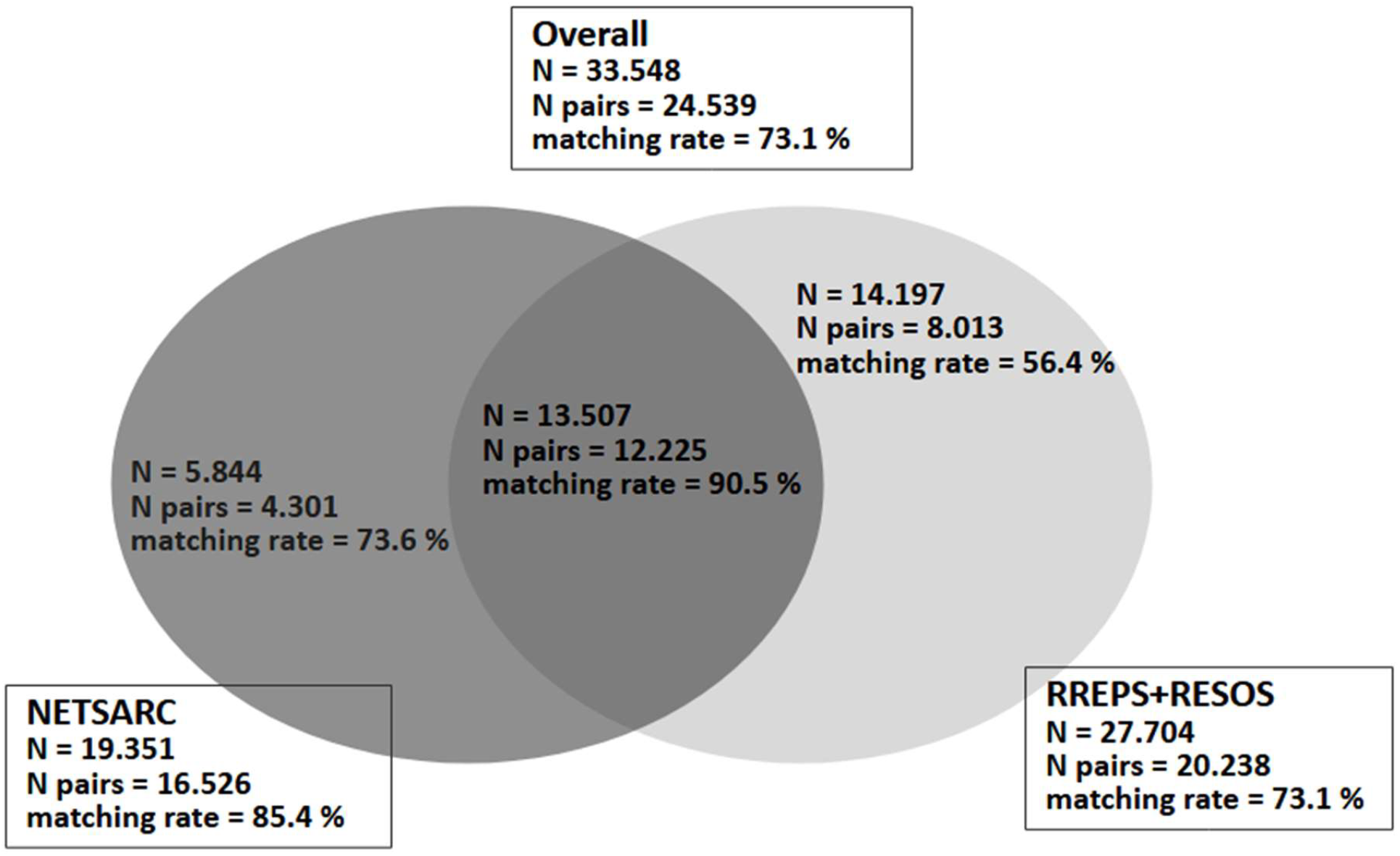
Definitive matching rates of the pairing algorithm by origin of the « source » database NETSARC stands for the sarcoma clinical database, RREPS and RESOS stands for the pathological review database for soft tissues and viscera sarcoma and for bone sarcoma, respectively.

Table 2 gives some characteristics of the 2 subgroups according to the origin of the « source » database with NETSARC for clinical data, RREPS+RESOS for pathological data, INTERSECT for the patients present in both databases and OVERALL for the whole population.

The pairing algorithm produced 24,539 unique pairs between the source and the target databases. Those patients are divided into 104 distinct signatures.

A partial view of the results for the 20 first signatures representing almost 89 % of the total pairs (n=21,835 pairs) is presented in table 3, giving for each signature, the number of matching pairs and some parameters of the distribution of the robustness variable. The extensive results are available in the Supplementary Material/section D. It is finally noticeable that the complete signature (SsDLlCFRMO) is only found for 7 patients which seems to indicate that the algorithm is quite resilient to a partial absence of information since most of the pairs are found with missing chaining variables.

**Table 3.**
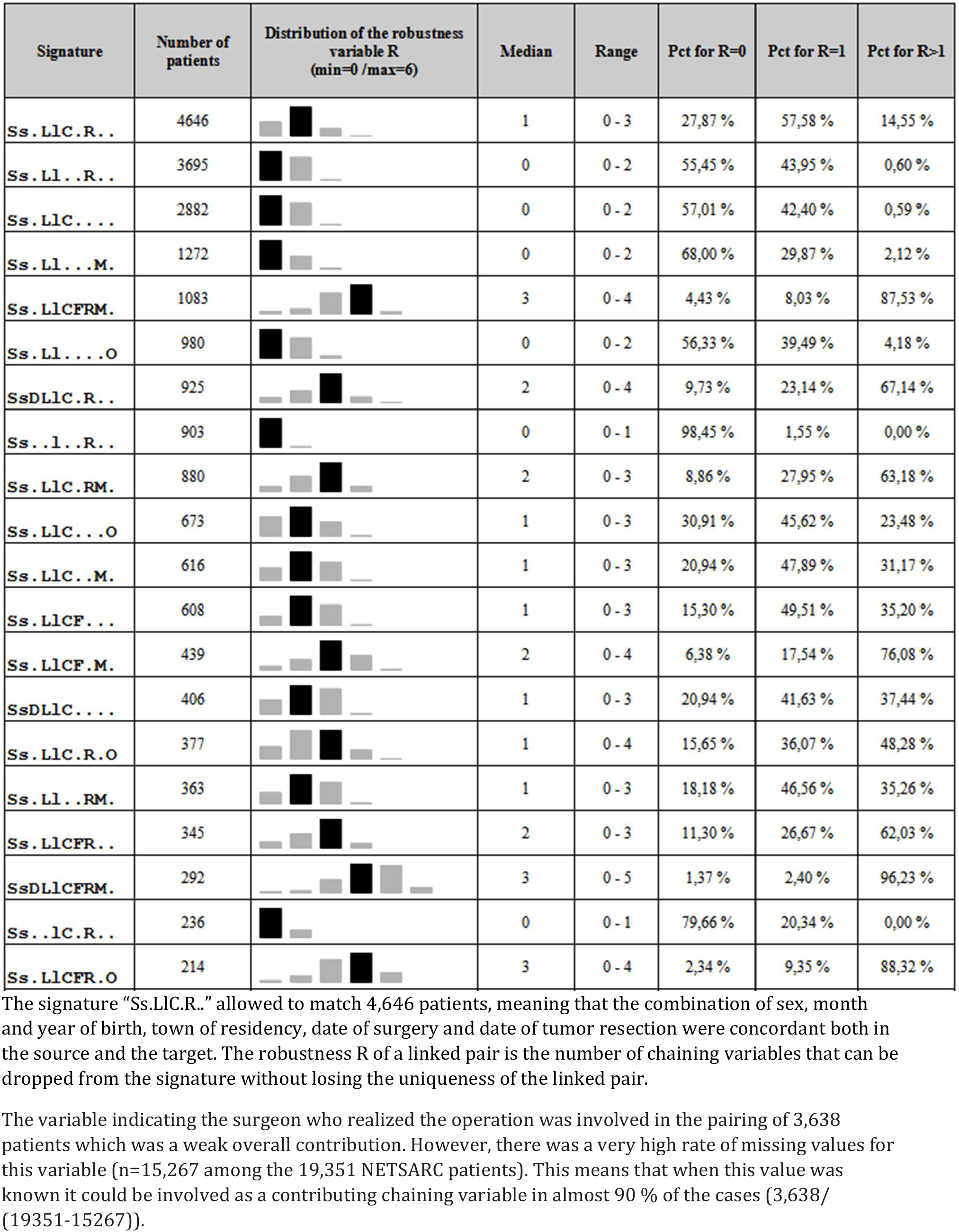
Number of unique matching pairs for the 20 first signatures (n=21,835 patients) with details of the distribution of the robustness variable

Figure 3 presents the distribution of the quality metric for the paired patients ranging from 0 to 7, the grey sector representing the non-paired patients (n = 9.009 patients).

**Figure 3.**
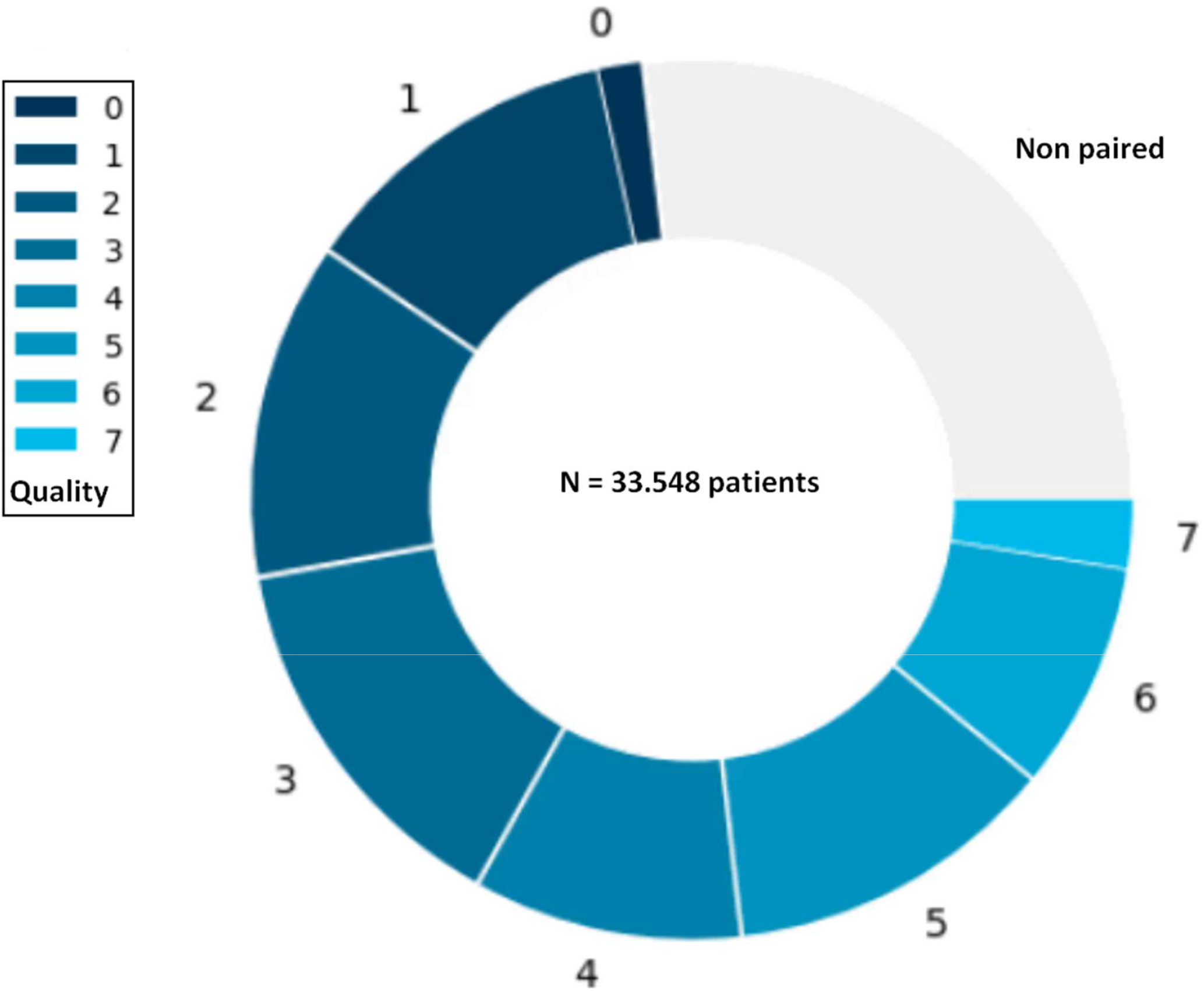
Distribution of the quality metric for the paired patients

## Discussion

The linking process for the DEEPSARC project faced several challenges: firstly, the heterogeneity of data and completeness, considering a clinical database (NETSARC) and two pathological databases (RREPS and RESOS) on the one hand, and a health insurance database on the other hand; secondly, the large amount of observation, 33,548 patients on the one hand (including 13,507 patients, 40.3%, with extended data, clinical as well as pathological), and 3 million patients who underwent surgery for their cancer between 2010 and 2018, on the other hand.

An indirect, stepwise, combinatory approach (set theory) allowed an overall matching rate of 73.1% (24,539 pairs out of 33,548 observations), reaching 90.5% in the intersection of the sarcomas databases (with extended data, 12,225 pairs out of 13,507 observations). In addition, two metric, robustness and quality allowed discussion and interpretation of the linking results.

A number of limitations need to be discussed. The first is that the result of linkage is intimately related to the quality of the data used. In our example, the two databases (NETSARC and RREPS) have a different number of candidate variables, and the quality of the data is also very different, both in terms of typology and discriminative capacity, as well as the percentage of missing data. Logically, not only the linkage rate, but also the linkage quality metrics are different, and in the expected direction. This is where the importance of producing these metrics to describe the result of linkage becomes apparent.

A second limitation is the choice of integrating an endpoint (date of death when a survival analysis is planned) as a linking variable: subjects who died within the observation window have a higher probability of being “well” linked, especially as the information will be collected more reliably for individuals who died in hospital, who could therefore be over-represented among linked subjects. Here again, the metrics delivered, and in particular the robustness among decedents, provide important elements for the discussion of potential selection bias. Among the 24,539 paired patients, 2,613 (10,88%) showed a signature involving the death date. For the very majority of cases (2,331 out of 2,613 i.e. 89,2%) the robustness of the signatures was greater than 0. This means that the death date in the pairing process brought mainly a reinforcement of the confidence of the uniqueness of the pair but was very marginally the crucial variable to establish a unique pair, since it was not possible to remove this variable in only 282 pairs.

A third limitation is selection bias related to non-linking. Even a very high linkage rate does not obviate the issue of bias in statistical analysis. It is therefore important to describe and compare linked and unlinked individuals in order to identify any association between the linkage result and the characteristics of the individuals. In our example, an individual present only in the RREPS database has a greater probability of not being linked, and the characteristics of these individuals are different from those of individuals in the NETSARC database, for example in terms of a contact with an expert center for their care.

A final limitation is a bias related to the quality of the pairs constituted, since quality is correlated with the number and availability of linking variables. It is important to be able to conduct sensitivity analyses according to the quality metric.

Several strengths are worth highlighting. First of all, the software suite used can handle databases of several million individuals. This scalability is essential when it comes to linking a clinical database with a medico-administrative database. Indeed, the combinatorial and agnostic approach compares the signature (combinations of chaining variables) of an individual in the source database with the signatures of all the individuals in the target database. A data organization trick is used to break the computational lock.

Secondly, the software suite is flexible: it allows parameterization of linking variables, such as authorizing a delay between two dates. The speed of processing enables parameters to be adjusted on the fly, and results to be compared as a function of this adjustment.

Thirdly, the software is resilient to missing data when dates are available. The first 4 signatures represent more than 50 % of the paired patients (n=12,495) and they only rely on a weak signature with 5 or 6 variables among the 10 potential chaining variables, in general an event date (or 2) along with the demographic variables. This indicates that knowing only a date (surgery date, tumor resection date or micro-biopsy date) is very discriminant for the pairing process and in more than half of the time even sufficient to find a unique pair. However, those 4 groups present generally a very low value for robustness (median=0 when only one date is known as in SS.LL..R.., SS.LLC.…or SS.LL…M., while median=1 when 2 dates are known as in Ss.LlC.R..) reinforcing the idea that the date variable is essential otherwise the uniqueness of the pair is lost.

Finally, the software suite calculates elements for understanding the linking result.^9^ This interpretability is based on the production of signatures for linked pairs, signatures for unlinked individuals, and a quality metric. Robustness is a good indicator for comparing pairs within the same signature group. Indeed, the level of confidence in matching a patient with robustness R=2 will be higher than if R=0. However, robustness is less relevant for comparing patients belonging to different signature classes. Indeed, if the signature contains, for example, 6 chaining variables, obtaining an R=2 robustness for a patient does not have the same significance as for a patient with the same robustness but whose signature contains only 3 chaining variables. Quality is a metric that can overcome intra-class variability and enable inter-class comparisons of signatures. The advantage is that it takes in account both the notion of robustness and the context, linked to the information available in the « source » and « target » databases for a given patient, thus smoothing out inter-signature variability.

In summary, all these elements contribute to an essential concept: the transparency of the process, with the production of a detailed chaining report, enabling essential discussion and interpretation of the chaining results.

## Data Availability

Access to the data by an independent expert mandated by a scientific publisher shall be carried out under the conditions set out in Deliberation No. 2018-155 of May 3, 2018, approving the French Reference Methodology (MR-004) for the processing of personal data implemented in the context of research not involving human participants, studies, and evaluations in the field of health.

## LIST OF ABBREVIATIONS

CepiDC: Nationwide database containing data on causes of death derived from the treatment of certificates of death
CNIL: National Commission for Information Technology and Civil Liberties (French data protection authority)
HDH: “Plateforme des données de Santé” Health Data Hub
ICD: International Classification of Disease
MDTM: Multidisciplinary team meeting
NETSARC+: Sarcoma reference network since 2020 (merger of the 3 pre-existing sarcoma networks since 2010: RRePS (pathological double review of soft tissue / visceral sarcoma), NetSarc (clinical management of soft tissue / visceral sarcoma) and ResOs (bone sarcoma)
PMSI: Nationwide database containing data derived from the healthcare activity delivered in private and public establishments
SNDS: *“Système National des Données de Santé”* (French National Health Data System)
SNIIRAM: Nationwide database containing health insurance data derived from the processing of healthcare reimbursements, mainly for ambulatory care

## Statements relating to ethics and integrity policies

access to the data by an independent expert mandated by a scientific publisher shall be carried out under the conditions set out in Deliberation No. 2018-155 of May 3, 2018, approving the French Reference Methodology (MR-004) for the processing of personal data implemented in the context of research not involving human participants, studies, and evaluations in the field of health.

## Funding supported

NETSARC+ (INCa-DGOS), RRePS (INCa -DGOS), RESOS (INCa-DGOS), INTERSARC+ (INCa), LabEx DEvweCAN (ANR-10-LABX-0061), LYriCAN+ (INCa-DGOS-INSERM-ITMO cancer_18003), Ligue Nationale contre le Cancer, Ligue contre le Cancer (Comité de l’Ain), Fondation ARC, and EURACAN (EU project 739521). The DEEPSARC Project was granted by Health Data Hub.

## Conflict of interest disclosure

None.

## Author contribution for each co-author by alphabetical order

Manuscript preparation: E DREZEN, A HAPPE and E OGER.

Concept and design of the linkage process: E DREZEN, A HAPPE and E OGER.

Data production (ICD-10-NETSARC mapping): C CHEMIN AIRIAU.

DEEPSARC Project Management: H CROCHET and F DUCIMETIERE.

Linkage software implementation and interpretation: E DREZEN.

Critical review: JY BLAY, G DU BOUEXIC DE PINIEUX, F GOUIN, F LE LOARER, S MATHOULIN PELISSIER, N PENEL, and V THEVENET.

Regulatory approval (DR 2020 360) by CNIL on November 20, 2020.

## Acknowledgements

We thank all contributors to data collection, analysis, or writing/editing, all physicians, all pathologists, all clinical research associate and transversal operational team of the NETSARC+ network.

## Notes

### Competing Interest Statement

The authors have declared no competing interest.

### Author Declarations

The study used de-identified individual-level data. Regulatory approval (DR 2020 360) by Commission Nationale Informatique et Libertes (CNIL) on November 20, 2020

## References

1. Ray-Coquard I, Montesco MC, Coindre JM, Dei Tos AP, Lurkin A, Ranchère-Vince D, et al. Sarcoma: concordance between initial diagnosis and centralized expert review in a population-based study within three European regions. Ann Oncol. 2012; 23(9):2442–2449.

2. Blay JY, Soibinet P, Penel N, Bompas E, Duffaud F, Stoeckle E, et al. Improved survival using specialized multidisciplinary board in sarcoma patients. Ann Oncol. 2017; 28(11):2852–2859.

3. Blay JY, Penel N, Valentin T, Anract P, Duffaud F, Dufresne A, et al. Improved nationwide survival of sarcoma patients with a network of reference centers. Ann Oncol. 2024; 35(4):351–363.

4. Fayet Y, Chevreau C, Decanter G, Dalban C, Meeus P, Carrère S, et al. No Geographical Inequalities in Survival for Sarcoma Patients in France: A Reference Networks’ Outcome? Cancers (Basel). 2022; 14(11):2620.

5. Tuppin P, Rudant J, Constantinou P, Gastaldi-Ménager C, Rachas A, de Roquefeuil L, et al. Value of a national administrative database to guide public decisions: From the système national d’information interrégimes de l’Assurance Maladie (SNIIRAM) to the système national des données de santé (SNDS) in France. Rev Epidemiol Sante Publique. 2017;65(Suppl 4):S149–S167.

6. Scailteux LM, Droitcourt C, Balusson F, Nowak E, Kerbrat S, Dupuy A, et al. French administrative health care database (SNDS): The value of its enrichment. Therapie. 2019 Apr;74(2):215–223.

7. Hess LM, Zhu YE, Sugihara T, Fang Y, Collins N, Nicol S. Challenges of Using ICD-9-CM and ICD-10-CM. Codes for Soft-Tissue Sarcoma in Databases for Health Services Research. Perspect Health Inf Manag. 2019; 16(Spring):1a.

8. Winglee M, Valliant R, Scheuren F: A case study in record linkage. Survey Methodology. 2005, 31 (1): 3–11.

9. Drézen E, Happe A, Kerbrat S, Balusson F, Oger E. New metrics for assessing linkage quality in deterministic record linkage of health databases. 2022. hal-03601245

