## Supplementary material for "Linking the NETSARC+ national sarcoma database with the SNDS to evaluate adjuvant and/or neoadjuvant therapy: report on the linkage process and result (Health Data Hub’s DEEPSARC pilot project)": Text and Tables

***Section A: Criteria for selection of the SNDS subpopulation***

The selection of the population concerns medical, or surgical, or obstetric hospital stays (PMSI MCO database) from 01/01/2010 to 12/31/2018 included. A search for surgical operations related to sarcomas is carried out by selecting the surgery DRG for disease related groups (GHM code in French for "*Groupe Homogène de Malades*"). The GHM codes will be searched in the annual tables of the PMSI MCO at the hospital stay (RSS) level (T_MCOaaB) in the fields GHM or (logical OR operator) GHM whose 3rd character is a 'C' (SUBSTR operator). To establish the completeness of the diagnoses between 01/01/2010 and 12/31/2017 it is imperative to take in account the surgeries also occurring in 2018. This criterion will be combined with an AND operator (logical AND) on the codes CIM-10 searched (see attached list) in the annual tables of the PMSI MCO at the RSS level (T_MCOaaB) in the fields DGN_PAL (main diagnosis) or (logical OR operator) DGN_REL (related diagnosis) as well as (logical OR operator) in the PMSI MCO tables at the medical or surgical unit (RUM) level (T_MCOaaUM) in the fields DGN_PAL (main diagnosis), and DGN_REL (related diagnosis). The pseudo-code writing of the extraction request is as follows:

```
(SUBSTR (T_MCOaaB. GRC_GHM,3,1) = 'C' OR SUBSTR (T_MCOaaB. GRG_GHM,3,1) = 'C')

AND     (T_MCOaaB.DGN_PAL in (liste_CIM-10) OR T_MCOaaB. DGN_REL in (liste_CIM-10)

        OR

        T_MCOaaUM.DGN_PAL in (liste_CIM-10) OR T_MCOaaUM. DGN_REL in (liste_CIM-10))

AND     (T_MCOaaC.ENT_DAT between 01/01/2010 and 31/12/2018)
```

A search for all the identities of the beneficiaries presenting the extraction characteristics is carried out to guarantee the completeness of patient pathways. Then, all the data corresponding to all these identities will be extracted from the DCIR, PMSI MCO, PMSI SSR, PMSI HAD, RIM-P, from the beneficiary repository (IR_BEN_R and IR_BEN_ARC), from the medicalized beneficiary repository (IR_IMB_R), as well as all tables relating to medical causes of death (K_xxx_R), making it possible to establish the complete care pathway for patients between 01/01/2009 and 31/12/2018.

***Section B: Rules for semantic alignment of chaining variables in both databases***

Table 1. Linkage Management Book of the DEEPSARC project

| Letter in the signature | Chaining variable name | Margin of error on date (± days) | Origin of the variable |
|---|---|---|---|
| **S** | Sex code + month and year of birth | 0 | NETSARC & RREPS |
| **s** | Sex code + year of birth | 0 | NETSARC & RREPS |
| **D** | Death date | 20 | NETSARC |
| **L** | Code of residency town | 0 | NETSARC & RREPS |
| **l** | Code of department of residency | 0 | NETSARC & RREPS |
| **C** | Surgery date + Type of tumor | 10 | NETSARC |
| **F** | Code of the surgeon | 10 | NETSARC |
| **R** | Tumor resection | 20 | RREPS |
| **M** | Micro biopsy | 20 | RREPS |
| **O** | Open biopsy | 20 | RREPS |

**Chaining variables in the source database (NETSARC + RREPS/RESOS)**

• S and s: Use of the information 'Sex' (ID=141 in NETSARC + RREPS/RESOS) and 'Birth date' (ID=140 in NETSARC + RREPS/RESOS)
Value = 1 if Sex=='Male' / 2 if Sex=='Female' + association with month and year of birth for S, only year of birth for s. This variable is calculated for the clinical and the pathological databases

• D: Use of the information 'Date of last contact' (ID=76 in NETSARC) and 'Vital status' (ID=1075 in NETSARC)
Value = the date of last contact when the vital status of the patient is 'Dead'. This variable is only available in the clinical database.

• L and l: Use of the information 'Geographic origin' combined with the level (Town/Department/Region)
Value = the geographic code of residency town (source https://insee.fr) for L or the department residency for l. No date is associated with this variable, which is calculated for the clinical and the pathological databases

• F: Use of the information 'Surgeon' (ID=1096 in NETSARC) and 'Date of surgery' (ID=1079 in NETSARC)
Value = A mapping table is built between the name of the surgeon and its FINESS code (source https://finess.esante.gouv.fr/) then the FINESS code is associated with the date of the surgery. This variable is only available in the clinical database.

• C: Use of the information 'Type of tumor' (ID=1091 in NETSARC) and 'Date of surgery' (ID=1079 in NETSARC)

Value = type of tumor corresponding to the 3 modalities of sarcoma “Soft tissue”, “Bone”, “Viscera” associated with the date of surgery. This variable is only available in the clinical database.

• R: Use of the information ‘Type of sampling’ (ID=21 in RREPS/RESOS) and ‘Date of sampling’ (ID=146 in RREPS/RESOS)
Value = The « tumor resection » corresponds to the date of sampling when the type of sampling is ’Tumor resection”. This variable is only available in the pathological database.

• M: Use of the information ‘Type of sampling’ (ID=21 in RREPS/RESOS) , ‘Date of sampling’ (ID=146 in RREPS/RESOS), ‘Type of tumor’ (ID=1091 in RREPS/RESOS) and ‘Site of tumor’ (ID=2 in RREPS/RESOS)
Value = The « Micro-biopsy » corresponds to ‘Site of tumor + Type of tumor’ if the type of sampling is ‘Micro-biopsy‘ associated with the date of sampling. This variable is only available in the pathological database.

• O: Use of the information ‘Type of sampling’ (ID=21 in RREPS/RESOS) , ‘Date of sampling’ (ID=146 in RREPS/RESOS), ‘Type of tumor’ (ID=1091 in RREPS/RESOS) and ‘Site of tumor’ (ID=2 in RREPS/RESOS)
Value = The « Open biopsy » corresponds to ‘Site of tumor + Type of tumor’ if the type of sampling is ‘Open biopsy‘ associated with the date of sampling. This variable is only available in the pathological database.

***Chaining variables in the target database (SNDS)***

• S and s: the IR_BEN_R table (as well as in its archived version) is used for the sex code (BEN_SEX_COD), the month of birth (BEN_NAI_MOI) and the year of birth (BEN_NAI_ANN)

• D: Date of death can be found in different places:

◦ in table IR_BEN_R (column BEN_DCD_DTE)

◦ in the KI_CCI_R table of medical causes of death (column BEN_DCD_DTE)

◦ as the end date of the stay when value=9 for hospitalization discharge mode (PMSI MCO/SSR/HAD)

• L and l: the patient's localization of residency is searched at two levels:

◦ town level: BEN_RES_DPT and BEN_RES_COM columns of tables IR_BEN_R and ER_PRS_F

◦ department level: column BEN_RES_DPT for the IR_BEN_R and ER_PRS_F tables, and column BDI_DEP for the PMSI/MCO tables

• F: the FINESS code of the surgeon is searched in the ETA_NUM column of the T_MCOaaB table of the PMSI

• C: As a reminder, on the « source » side, this variable includes the modalities of “Type of tumor” (“Soft tissue”, “Bone”, “Viscera”). On the SNDS side, surgeries will be searched mainly based on DRG codes (GHM) listed below, the two first characters of this GHM code, which indicates an anatomic localization, allowing a mapping with the NETSARC variable « Site of tumor ». In addition, a few ICD-10 codes (CIM-10) and CCAM procedures frequently associated with those DRG were added in order to extend the spectrum of the variable, they are listed below.

| Type | Classification | Code | Label | Found in SNDS (target) |
|---|---|---|---|---|
| Bone | GHM (DRG) | 01C041 | Non-traumatic craniotomies, age over 17, level 1 | T_MCOaaB/GRC_GHM |
| Bone | GHM (DRG) | 01C042 | Non-traumatic craniotomies, age over 17, level 2 | T_MCOaaB/GRC_GHM |
| Bone | GHM (DRG) | 01C043 | Non-traumatic craniotomies, age over 17, level 3 | T_MCOaaB/GRC_GHM |
| Bone | GHM (DRG) | 01C051 | Spine and cord surgery for neurological conditions, level 1 | T_MCOaaB/GRC_GHM |
| Bone | GHM (DRG) | 01C052 | Spine and cord procedures for neurological conditions, level 2 | T_MCOaaB/GRC_GHM |
| Bone | GHM (DRG) | 01C053 | Spine and cord procedures for neurological conditions, level 3 | T_MCOaaB/GRC_GHM |
| Bone | GHM (DRG) | 01C054 | Spine and cord procedures for neurological conditions, level 4 | T_MCOaaB/GRC_GHM |
| Bone | GHM (DRG) | 01C111 | Craniotomies for tumors, age under 18, level 1 | T_MCOaaB/GRC_GHM |
| Bone | GHM (DRG) | 01C112 | Craniotomies for tumors, age under 18, level 2 | T_MCOaaB/GRC_GHM |
| Bone | GHM (DRG) | 01M27 | Other nervous system tumors | T_MCOaaB/GRC_GHM |
| Bone | GHM (DRG) | 03C071 | Procedures on the sinuses and mastoid process, age over 17, level 1 | T_MCOaaB/GRC_GHM |
| Bone | GHM (DRG) | 03C074 | Interventions on the sinuses and mastoid process, age over 17, level 4 | T_MCOaaB/GRC_GHM |
| Bone | GHM (DRG) | 03C161 | Other ear, nose, throat or neck surgery, level 1 | T_MCOaaB/GRC_GHM |
| Bone | GHM (DRG) | 03C171 | Mouth procedures, level 1 | T_MCOaaB/GRC_GHM |
| Bone | GHM (DRG) | 03C251 | Major head and neck procedures, level 1 | T_MCOaaB/GRC_GHM |
| Bone | GHM (DRG) | 03C252 | Major head and neck procedures, level 2 | T_MCOaaB/GRC_GHM |
| Bone | GHM (DRG) | 03C253 | Major head and neck procedures, level 3 | T_MCOaaB/GRC_GHM |
| Bone | GHM (DRG) | 03C254 | Major head and neck procedures, level 4 | T_MCOaaB/GRC_GHM |
| Bone | GHM (DRG) | 03C261 | Other head and neck procedures, level 1 | T_MCOaaB/GRC_GHM |
| Bone | GHM (DRG) | 03C262 | Other head and neck procedures, level 2 | T_MCOaaB/GRC_GHM |
| Bone | GHM (DRG) | 03C263 | Other head and neck procedures, level 3 | T_MCOaaB/GRC_GHM |
| Bone | GHM (DRG) | 03C264 | Other head and neck procedures, level 4 | T_MCOaaB/GRC_GHM |
| Bone | GHM (DRG) | 03C291 | Other ear, nose or throat procedures for malignant tumors, level 1 | T_MCOaaB/GRC_GHM |
| Bone | GHM (DRG) | 03C292 | Other ear, nose or throat procedures for malignant tumors, level 2 | T_MCOaaB/GRC_GHM |
| Bone | GHM (DRG) | 03C293 | Other ear, nose or throat procedures for malignant tumors, level 3 | T_MCOaaB/GRC_GHM |
| Bone | GHM (DRG) | 04C021 | Major thoracic procedures, level 1 | T_MCOaaB/GRC_GHM |
| Bone | GHM (DRG) | 04C022 | Major thoracic procedures, level 2 | T_MCOaaB/GRC_GHM |
| Bone | GHM (DRG) | 04C023 | Major thoracic procedures, level 3 | T_MCOaaB/GRC_GHM |
| Bone | GHM (DRG) | 04C024 | Major thoracic procedures, level 4 | T_MCOaaB/GRC_GHM |
| Bone | GHM (DRG) | 04C041 | Thoracoscopic procedures, level 1 | T_MCOaaB/GRC_GHM |
| Bone | GHM (DRG) | 04C042 | Thoracoscopic procedures, level 2 | T_MCOaaB/GRC_GHM |
| Bone | GHM (DRG) | 05C111 | Other vascular surgery, level 1 | T_MCOaaB/GRC_GHM |
| Bone | GHM (DRG) | 06C151 | Other digestive tract procedures other than laparotomy, level 1 | T_MCOaaB/GRC_GHM |
| Bone | GHM (DRG) | 06M05 | Other malignant tumors of the digestive tract | T_MCOaaB/GRC_GHM |
| Bone | GHM (DRG) | 08C021 | Multiple major knee and/or hip procedures, level 1 | T_MCOaaB/GRC_GHM |
| Bone | GHM (DRG) | 08C022 | Multiple major knee and/or hip procedures, level 2 | T_MCOaaB/GRC_GHM |
| Bone | GHM (DRG) | 08C023 | Multiple major knee and/or hip procedures, level 3 | T_MCOaaB/GRC_GHM |
| Bone | GHM (DRG) | 08C024 | Multiple major knee and/or hip procedures, level 4 | T_MCOaaB/GRC_GHM |
| Bone | GHM (DRG) | 08C041 | Hip and femur procedures, age under 18, level 1 | T_MCOaaB/GRC_GHM |
| Bone | GHM (DRG) | 08C042 | Hip and femur procedures, age under 18, level 2 | T_MCOaaB/GRC_GHM |
| Bone | GHM (DRG) | 08C043 | Hip and femur procedures, age under 18, level 3 | T_MCOaaB/GRC_GHM |
| Bone | GHM (DRG) | 08C044 | Hip and femur procedures, age under 18, level 4 | T_MCOaaB/GRC_GHM |
| Bone | GHM (DRG) | 08C061 | Amputations for musculoskeletal and connective tissue disorders, level 1 | T_MCOaaB/GRC_GHM |
| Bone | GHM (DRG) | 08C062 | Amputations for musculoskeletal and connective tissue disorders, level 2 | T_MCOaaB/GRC_GHM |
| Bone | GHM (DRG) | 08C063 | Amputations for musculoskeletal and connective tissue disorders, level 3 | T_MCOaaB/GRC_GHM |
| Bone | GHM (DRG) | 08C064 | Amputations for musculoskeletal and connective tissue disorders, level 4 | T_MCOaaB/GRC_GHM |
| Bone | GHM (DRG) | 08C121 | Osteoarticular biopsies, level 1 | T_MCOaaB/GRC_GHM |
| Bone | GHM (DRG) | 08C122 | Osteoarticular biopsies, level 2 | T_MCOaaB/GRC_GHM |
| Bone | GHM (DRG) | 08C12J | Outpatient osteoarticular biopsies | T_MCOaaB/GRC_GHM |
| Bone | GHM (DRG) | 08C131 | Localized bone resections and/or removal of internal fixation hardware in the hip and femur, level 1 | T_MCOaaB/GRC_GHM |
| Bone | GHM (DRG) | 08C132 | Localized bone resections and/or removal of internal fixation devices in the hip and femur, level 2 | T_MCOaaB/GRC_GHM |
| Bone | GHM (DRG) | 08C133 | Localized bone resections and/or removal of internal fixation hardware in the hip and femur, level 3 | T_MCOaaB/GRC_GHM |
| Bone | GHM (DRG) | 08C141 | Localized bone resections and/or removal of internal fixation hardware at sites other than the hip and femur, level 1 | T_MCOaaB/GRC_GHM |
| Bone | GHM (DRG) | 08C142 | Localized bone resections and/or removal of internal fixation in sites other than hip and femur, level 2 | T_MCOaaB/GRC_GHM |

| | | | | |
|---|---|---|---|---|
| Bone | GHM (DRG) | 08C143 | Localized bone resections and/or removal of internal fixation hardware at sites other than the hip and femur, level 3 | T_MCOaaB/GRC_GHM |
| Bone | GHM (DRG) | 08C14J | Localized bone resections and/or removal of internal fixation hardware at sites other than the hip and femur, on an outpatient basis | T_MCOaaB/GRC_GHM |
| Bone | GHM (DRG) | 08C201 | Skin grafts for musculoskeletal or connective tissue disease, level 1 | T_MCOaaB/GRC_GHM |
| Bone | GHM (DRG) | 08C202 | Skin grafts for musculoskeletal or connective tissue disease, level 2 | T_MCOaaB/GRC_GHM |
| Bone | GHM (DRG) | 08C203 | Skin grafts for musculoskeletal or connective tissue disease, level 3 | T_MCOaaB/GRC_GHM |
| Bone | GHM (DRG) | 08C204 | Skin grafts for musculoskeletal or connective tissue disease, level 4 | T_MCOaaB/GRC_GHM |
| Bone | GHM (DRG) | 08C211 | Other musculoskeletal and connective tissue procedures, level 1 | T_MCOaaB/GRC_GHM |
| Bone | GHM (DRG) | 08C212 | Other musculoskeletal and connective tissue procedures, level 2 | T_MCOaaB/GRC_GHM |
| Bone | GHM (DRG) | 08C213 | Other musculoskeletal and connective tissue procedures, level 3 | T_MCOaaB/GRC_GHM |
| Bone | GHM (DRG) | 08C214 | Other musculoskeletal and connective tissue procedures, level 4 | T_MCOaaB/GRC_GHM |
| Bone | GHM (DRG) | 08C221 | Joint replacement procedures, level 1 | T_MCOaaB/GRC_GHM |
| Bone | GHM (DRG) | 08C222 | Joint replacement procedures, level 2 | T_MCOaaB/GRC_GHM |
| Bone | GHM (DRG) | 08C223 | Joint replacement procedures, level 3 | T_MCOaaB/GRC_GHM |
| Bone | GHM (DRG) | 08C224 | Revision joint replacement, level 4 | T_MCOaaB/GRC_GHM |
| Bone | GHM (DRG) | 08C241 | Knee prostheses, level 1 | T_MCOaaB/GRC_GHM |
| Bone | GHM (DRG) | 08C242 | Knee prostheses, level 2 | T_MCOaaB/GRC_GHM |
| Bone | GHM (DRG) | 08C243 | Knee replacement, level 3 | T_MCOaaB/GRC_GHM |
| Bone | GHM (DRG) | 08C244 | Knee prostheses, level 4 | T_MCOaaB/GRC_GHM |
| Bone | GHM (DRG) | 08C251 | Shoulder prostheses, level 1 | T_MCOaaB/GRC_GHM |
| Bone | GHM (DRG) | 08C252 | Shoulder prostheses, level 2 | T_MCOaaB/GRC_GHM |
| Bone | GHM (DRG) | 08C253 | Shoulder prostheses, level 3 | T_MCOaaB/GRC_GHM |
| Bone | GHM (DRG) | 08C271 | Other spinal procedures, level 1 | T_MCOaaB/GRC_GHM |
| Bone | GHM (DRG) | 08C272 | Other spinal procedures, level 2 | T_MCOaaB/GRC_GHM |
| Bone | GHM (DRG) | 08C273 | Other spinal procedures, level 3 | T_MCOaaB/GRC_GHM |
| Bone | GHM (DRG) | 08C274 | Other spinal procedures, level 4 | T_MCOaaB/GRC_GHM |
| Bone | GHM (DRG) | 08C281 | Maxillofacial procedures, level 1 | T_MCOaaB/GRC_GHM |
| Bone | GHM (DRG) | 08C282 | Maxillofacial procedures, level 2 | T_MCOaaB/GRC_GHM |
| Bone | GHM (DRG) | 08C283 | Maxillofacial procedures, level 3 | T_MCOaaB/GRC_GHM |
| Bone | GHM (DRG) | 08C284 | Maxillofacial procedures, level 4 | T_MCOaaB/GRC_GHM |
| Bone | GHM (DRG) | 08C28J | Outpatient maxillofacial procedures | T_MCOaaB/GRC_GHM |
| Bone | GHM (DRG) | 08C291 | Soft-tissue procedures for malignant tumors, level 1 | T_MCOaaB/GRC_GHM |
| Bone | GHM (DRG) | 08C292 | Soft-tissue procedures for malignant tumors, level 2 | T_MCOaaB/GRC_GHM |
| Bone | GHM (DRG) | 08C293 | Soft-tissue procedures for malignancies, level 3 | T_MCOaaB/GRC_GHM |
| Bone | GHM (DRG) | 08C29J | Outpatient soft-tissue procedures for malignant tumors | T_MCOaaB/GRC_GHM |
| Bone | GHM (DRG) | 08C311 | Leg procedures, age under 18, level 1 | T_MCOaaB/GRC_GHM |
| Bone | GHM (DRG) | 08C312 | Leg procedures, age under 18, level 2 | T_MCOaaB/GRC_GHM |
| Bone | GHM (DRG) | 08C313 | Leg procedures, age under 18, level 3 | T_MCOaaB/GRC_GHM |
| Bone | GHM (DRG) | 08C314 | Leg procedures, age under 18, level 4 | T_MCOaaB/GRC_GHM |
| Bone | GHM (DRG) | 08C321 | Leg procedures, age greater than 17, level 1 | T_MCOaaB/GRC_GHM |
| Bone | GHM (DRG) | 08C322 | Leg procedures, age greater than 17, level 2 | T_MCOaaB/GRC_GHM |
| Bone | GHM (DRG) | 08C323 | Leg procedures, age greater than 17, level 3 | T_MCOaaB/GRC_GHM |
| Bone | GHM (DRG) | 08C331 | Procedures on the ankle and hindfoot excluding fractures, level 1 | T_MCOaaB/GRC_GHM |
| Bone | GHM (DRG) | 08C351 | Procedures on the arm, elbow and shoulder, level 1 | T_MCOaaB/GRC_GHM |
| Bone | GHM (DRG) | 08C352 | Procedures on the arm, elbow and shoulder, level 2 | T_MCOaaB/GRC_GHM |
| Bone | GHM (DRG) | 08C353 | Procedures on the arm, elbow and shoulder, level 3 | T_MCOaaB/GRC_GHM |
| Bone | GHM (DRG) | 08C354 | Upper arm, elbow and shoulder procedures, level 4 | T_MCOaaB/GRC_GHM |
| Bone | GHM (DRG) | 08C35J | Outpatient arm, elbow and shoulder procedures | T_MCOaaB/GRC_GHM |
| Bone | GHM (DRG) | 08C362 | Foot procedures, age under 18, level 2 | T_MCOaaB/GRC_GHM |
| Bone | GHM (DRG) | 08C371 | Foot procedures, age over 17, level 1 | T_MCOaaB/GRC_GHM |
| Bone | GHM (DRG) | 08C372 | Foot procedures, age over 17, level 2 | T_MCOaaB/GRC_GHM |
| Bone | GHM (DRG) | 08C384 | Other knee arthroscopies, level 4 | T_MCOaaB/GRC_GHM |
| Bone | GHM (DRG) | 08C391 | Forearm procedures, level 1 | T_MCOaaB/GRC_GHM |
| Bone | GHM (DRG) | 08C392 | Forearm procedures, level 2 | T_MCOaaB/GRC_GHM |
| Bone | GHM (DRG) | 08C421 | Non-minor soft-tissue procedures, level 1 | T_MCOaaB/GRC_GHM |
| Bone | GHM (DRG) | 08C422 | Non-minor soft-tissue procedures, level 2 | T_MCOaaB/GRC_GHM |
| Bone | GHM (DRG) | 08C431 | Non-minor hand procedures, level 1 | T_MCOaaB/GRC_GHM |

| | | | | |
|---|---|---|---|---|
| Bone | GHM (DRG) | 08C432 | Non-minor hand procedures, level 2 | T_MCOaaB/GRC_GHM |
| Bone | GHM (DRG) | 08C43J | Outpatient non-minor hand procedures | T_MCOaaB/GRC_GHM |
| Bone | GHM (DRG) | 08C461 | Other soft-tissue procedures, level 1 | T_MCOaaB/GRC_GHM |
| Bone | GHM (DRG) | 08C462 | Other soft-tissue procedures, level 2 | T_MCOaaB/GRC_GHM |
| Bone | GHM (DRG) | 08C463 | Other soft-tissue procedures, level 3 | T_MCOaaB/GRC_GHM |
| Bone | GHM (DRG) | 08C464 | Other soft-tissue procedures, level 4 | T_MCOaaB/GRC_GHM |
| Bone | GHM (DRG) | 08C46J | Other soft-tissue procedures, outpatient | T_MCOaaB/GRC_GHM |
| Bone | GHM (DRG) | 08C481 | Hip prostheses for conditions other than recent trauma, level 1 | T_MCOaaB/GRC_GHM |
| Bone | GHM (DRG) | 08C482 | Hip prostheses for conditions other than recent trauma, level 2 | T_MCOaaB/GRC_GHM |
| Bone | GHM (DRG) | 08C483 | Hip prostheses for conditions other than recent trauma, level 3 | T_MCOaaB/GRC_GHM |
| Bone | GHM (DRG) | 08C484 | Hip prostheses for conditions other than recent trauma, level 4 | T_MCOaaB/GRC_GHM |
| Bone | GHM (DRG) | 08C491 | Hip and femur procedures for recent trauma, age over 17, level 1 | T_MCOaaB/GRC_GHM |
| Bone | GHM (DRG) | 08C501 | Hip and femur procedures excluding recent trauma, age over 17, level 1 | T_MCOaaB/GRC_GHM |
| Bone | GHM (DRG) | 08C502 | Hip and femur procedures, excluding recent trauma, age over 17, level 2 | T_MCOaaB/GRC_GHM |
| Bone | GHM (DRG) | 08C503 | Hip and femur procedures, excluding recent trauma, age over 17, level 3 | T_MCOaaB/GRC_GHM |
| Bone | GHM (DRG) | 08C504 | Hip and femur procedures, excluding recent trauma, age over 17, level 4 | T_MCOaaB/GRC_GHM |
| Bone | GHM (DRG) | 08C521 | Other major spinal procedures, level 1 | T_MCOaaB/GRC_GHM |
| Bone | GHM (DRG) | 08C522 | Other major spinal procedures, level 2 | T_MCOaaB/GRC_GHM |
| Bone | GHM (DRG) | 08C523 | Other major spinal surgery, level 3 | T_MCOaaB/GRC_GHM |
| Bone | GHM (DRG) | 08C524 | Other major spine procedures, level 4 | T_MCOaaB/GRC_GHM |
| Bone | GHM (DRG) | 08C541 | Knee procedures for non-traumatic conditions, level 1 | T_MCOaaB/GRC_GHM |
| Bone | GHM (DRG) | 08C542 | Knee procedures for non-traumatic conditions, level 2 | T_MCOaaB/GRC_GHM |
| Bone | GHM (DRG) | 08C562 | Ankle and hindfoot procedures for fractures, level 4 | T_MCOaaB/GRC_GHM |
| Bone | GHM (DRG) | 08C581 | Shoulder arthroscopies, level 1 | T_MCOaaB/GRC_GHM |
| Bone | GHM (DRG) | 08C591 | Wrist tenosynovectomies, level 1 | T_MCOaaB/GRC_GHM |
| Bone | GHM (DRG) | 08C601 | Wrist procedures other than tenosynovectomies, level 1 | T_MCOaaB/GRC_GHM |
| Bone | GHM (DRG) | 08C612 | Major procedures for osteoarticular infections, level 2 | T_MCOaaB/GRC_GHM |
| Bone | GHM (DRG) | 11C022 | Kidney and ureter procedures and major bladder surgery for tumor disease, level 2 | T_MCOaaB/GRC_GHM |
| Bone | GHM (DRG) | 11C024 | Kidney and ureter procedures and major bladder surgery for tumoral disease, level 4 | T_MCOaaB/GRC_GHM |
| Bone | GHM (DRG) | 13M07 | Other tumors of the female genital tract | T_MCOaaB/GRC_GHM |
| Bone | GHM (DRG) | 17C042 | Major surgery for myeloproliferative disorders or tumors of unclear or diffuse location, level 2 | T_MCOaaB/GRC_GHM |
| Bone | GHM (DRG) | 17C043 | Major procedures for myeloproliferative disorders or tumors of unclear or diffuse site, level 3 | T_MCOaaB/GRC_GHM |
| Bone | GHM (DRG) | 17C051 | Other procedures for myeloproliferative disorders or tumors of unclear or diffuse location, level 1 | T_MCOaaB/GRC_GHM |
| Bone | GHM (DRG) | 17C052 | Other procedures for myeloproliferative disorders or tumors of unclear or diffuse site, level 2 | T_MCOaaB/GRC_GHM |
| Bone | GHM (DRG) | 17C053 | Other procedures for myeloproliferative disorders or tumors of unclear or diffuse site, level 3 | T_MCOaaB/GRC_GHM |
| Bone | GHM (DRG) | 17C054 | Other procedures for myeloproliferative disorders or tumors of imprecise or diffuse site, level 4 | T_MCOaaB/GRC_GHM |
| Bone | GHM (DRG) | 17C063 | Major procedures in CMD17 (Myeloproliferative disorders and tumors of unclear or diffuse location.), level 3 | T_MCOaaB/GRC_GHM |
| Bone | GHM (DRG) | 17C064 | Major procedures in CMD17 (Myeloproliferative disorders and tumors of unclear or diffuse location.), level 4 | T_MCOaaB/GRC_GHM |
| Bone | GHM (DRG) | 17C071 | Intermediate procedures of CMD17 (Myeloproliferative disorders and tumors of unclear or diffuse location.), level 1 | T_MCOaaB/GRC_GHM |
| Bone | GHM (DRG) | 17C073 | Intermediate procedures of CMD17 (Myeloproliferative disorders and tumors of unclear or diffuse location.), level 3 | T_MCOaaB/GRC_GHM |
| Bone | GHM (DRG) | 17M17 | Other conditions and tumors of imprecise or diffuse site | T_MCOaaB/GRC_GHM |
| Bone | GHM (DRG) | 21C052 | Other procedures for injuries or complications, level 2 | T_MCOaaB/GRC_GHM |
| Bone | CIM-10 (ICD10) | C031 | Malignant neoplasm of lower gum | T_MCOaaB/DGN_PAL |
| Bone | CIM-10 (ICD10) | C310 | Malignant neoplasm of maxillary sinus | T_MCOaaB/DGN_PAL |
| Bone | CIM-10 (ICD10) | C400 | Malignant tumor of the scapula and long bones of the upper limb | T_MCOaaB/DGN_PAL |
| Bone | CIM-10 (ICD10) | C401 | Malignant tumor of the short bones of the upper limb | T_MCOaaB/DGN_PAL |
| Bone | CIM-10 (ICD10) | C402 | Malignant tumor of the long bones of the lower limb | T_MCOaaB/DGN_PAL |
| Bone | CIM-10 (ICD10) | C403 | Malignant tumor of the short bones of the lower limb | T_MCOaaB/DGN_PAL |
| Bone | CIM-10 (ICD10) | C408 | Malignant tumor of contiguous bone and articular cartilage of the limbs | T_MCOaaB/DGN_PAL |
| Bone | CIM-10 (ICD10) | C409 | Malignant tumor of the bones and articular cartilage of a limb, unspecified | T_MCOaaB/DGN_PAL |
| Bone | CIM-10 (ICD10) | C410 | Malignant neoplasm of bones of skull and face | T_MCOaaB/DGN_PAL |
| Bone | CIM-10 (ICD10) | C411 | Malignant neoplasm of mandible | T_MCOaaB/DGN_PAL |
| Bone | CIM-10 (ICD10) | C412 | Malignant neoplasm of vertebral column | T_MCOaaB/DGN_PAL |
| Bone | CIM-10 (ICD10) | C413 | Malignant neoplasm of ribs, sternum and clavicle | T_MCOaaB/DGN_PAL |
| Bone | CIM-10 (ICD10) | C414 | Malignant neoplasm of pelvic bones, sacrum and coccyx | T_MCOaaB/DGN_PAL |
| Bone | CIM-10 (ICD10) | C418 | Malignant tumor with contiguous localization of bone and articular cartilage | T_MCOaaB/DGN_PAL |

| | | | | |
|---|---|---|---|---|
| Bone | CIM-10 (ICD10) | C419 | Malignant neoplasm of bone and articular cartilage, unsp | T_MCOaaB/DGN_PAL |
| Bone | CIM-10 (ICD10) | C490 | Malig neoplm of conn and soft tissue of head, face and neck | T_MCOaaB/DGN_PAL |
| Bone | CIM-10 (ICD10) | C491 | Malignant tumors of connective tissue and other soft tissues of the upper limb, including the shoulder | T_MCOaaB/DGN_PAL |
| Bone | CIM-10 (ICD10) | C492 | Malignant tumor of the connective tissue and other soft tissues of the lower limb, including the hip | T_MCOaaB/DGN_PAL |
| Bone | CIM-10 (ICD10) | C4938 | Other malignant tumors of connective tissue and other soft tissues of the thorax | T_MCOaaB/DGN_PAL |
| Bone | CIM-10 (ICD10) | C4958 | Other malignant tumors of connective tissue and other soft tissues of the pelvis | T_MCOaaB/DGN_PAL |
| Bone | CIM-10 (ICD10) | C496 | Malignant tumor of connective tissue and other soft tissues of the trunk, unspecified | T_MCOaaB/DGN_PAL |
| Bone | CIM-10 (ICD10) | C763 | Malignant neoplasm of pelvis | T_MCOaaB/DGN_PAL |
| Bone | CIM-10 (ICD10) | C795 | Secondary malignant tumor of bone and bone marrow | T_MCOaaB/DGN_PAL |
| Bone | CIM-10 (ICD10) | D161 | Benign tumor of the short bones of the upper limb | T_MCOaaB/DGN_PAL |
| Bone | CIM-10 (ICD10) | D162 | Benign tumor of the long bones of the lower limb | T_MCOaaB/DGN_PAL |
| Bone | CIM-10 (ICD10) | D480 | Unpredictable and unknown tumor of bone and articular cartilage | T_MCOaaB/DGN_PAL |
| Bone | CCAM (procedures) | DEQP004 | Continuous electrocardiogram monitoring by oscilloscopy and/or remote monitoring, every 24 hours | T_MCOaaA/CDC_ACT |
| Bone | CCAM (procedures) | DZQM005 | Bedside transthoracic Doppler ultrasound of the heart and intrathoracic vessels | T_MCOaaA/CDC_ACT |
| Bone | CCAM (procedures) | DZQM006 | Transthoracic Doppler ultrasound of the heart and intrathoracic vessels | T_MCOaaA/CDC_ACT |
| Bone | CCAM (procedures) | MAQK003 | X-ray of shoulder girdle and/or shoulder, with 1 or 2 views | T_MCOaaA/CDC_ACT |
| Bone | CCAM (procedures) | MBFA001 | ”En bloc” resection of the tip and/or shaft of the humerus | T_MCOaaA/CDC_ACT |
| Bone | CCAM (procedures) | MBMA002 | Reconstruction of end and/or shaft of humerus after ”en bloc” resection, using graft or inert non-prosthetic material | T_MCOaaA/CDC_ACT |
| Bone | CCAM (procedures) | MBQK001 | Arm X-ray | T_MCOaaA/CDC_ACT |
| Bone | CCAM (procedures) | MEMA009 | Shoulder joint reconstruction with solid or custom prosthesis, after segmental resection | T_MCOaaA/CDC_ACT |
| Bone | CCAM (procedures) | NAFA001 | ” En bloc” resection of coxal bone or femur with monobloc hip arthrectomy | T_MCOaaA/CDC_ACT |
| Bone | CCAM (procedures) | NAFA003 | Complete “en bloc” partial or total resection of coxal bone [hemibasin] including acetabulum | T_MCOaaA/CDC_ACT |
| Bone | CCAM (procedures) | NAFA004 | Partial resection of the iliac wing or obturator frame without interrupting the continuity of the pelvic ring | T_MCOaaA/CDC_ACT |
| Bone | CCAM (procedures) | NAFA006 | Complete “en bloc” resection of the iliac wing or obturator frame interrupting the continuity of the pelvic ring | T_MCOaaA/CDC_ACT |
| Bone | CCAM (procedures) | NAMA002 | Reconstruction of the coxal bone [hemibasin] after resection of the acetabular zone, without coxal prosthesis | T_MCOaaA/CDC_ACT |
| Bone | CCAM (procedures) | NAQK007 | Pelvic girdle X-ray with 2 views | T_MCOaaA/CDC_ACT |
| Bone | CCAM (procedures) | NAQK015 | Pelvic girdle [pelvis] radiograph, 1 view | T_MCOaaA/CDC_ACT |
| Bone | CCAM (procedures) | NAQK071 | Radiograph of pelvic girdle (pelvis) at 1 angle and unilateral radiograph of coxofemoral joint at 1 or 2 angles | T_MCOaaA/CDC_ACT |
| Bone | CCAM (procedures) | NBFA007 | ”En bloc” resection of one end and/or the shaft of the femur | T_MCOaaA/CDC_ACT |
| Bone | CCAM (procedures) | NBMA001 | Reconstruction of femur using graft or inert non-prosthetic material, after “en bloc” resection of one end and/or shaft | T_MCOaaA/CDC_ACT |
| Bone | CCAM (procedures) | NBQK001 | Thigh X-ray | T_MCOaaA/CDC_ACT |
| Bone | CCAM (procedures) | NCFA001 | ”En bloc” resection of proximal end of 2 leg bones | T_MCOaaA/CDC_ACT |
| Bone | CCAM (procedures) | NCFA008 | ”En bloc” resection of one end and/or the shaft of the tibia | T_MCOaaA/CDC_ACT |
| Bone | CCAM (procedures) | NCFA009 | ”En bloc” resection of end and/or shaft of fibula | T_MCOaaA/CDC_ACT |
| Bone | CCAM (procedures) | NCMA001 | Reconstruction of tibia using graft or inert non-prosthetic material, after “en bloc” resection of end and/or shaft | T_MCOaaA/CDC_ACT |
| Bone | CCAM (procedures) | NCQK001 | X-ray of the leg | T_MCOaaA/CDC_ACT |
| Bone | CCAM (procedures) | NEMA011 | Reconstruction of the coxofemoral joint using a massive or custom-made prosthesis, after segmental loss of substance in the hip or coxal bone. | T_MCOaaA/CDC_ACT |
| Bone | CCAM (procedures) | NEQK010 | X-ray of the coxofemoral joint with 1 or 2 views | T_MCOaaA/CDC_ACT |
| Bone | CCAM (procedures) | NFKA009 | Knee joint replacement with fixed or rotating hinge prosthesis | T_MCOaaA/CDC_ACT |
| Bone | CCAM (procedures) | NFMA006 | Knee joint reconstruction with solid or custom prosthesis, after segmental loss of substance | T_MCOaaA/CDC_ACT |
| Bone | CCAM (procedures) | NFQK001 | Unilateral knee X-ray with 1 or 2 views | T_MCOaaA/CDC_ACT |
| Bone | CCAM (procedures) | NFQK003 | Knee X-ray with 3 or 4 views | T_MCOaaA/CDC_ACT |
| Bone | CCAM (procedures) | NZFA002 | Transtibial amputation | T_MCOaaA/CDC_ACT |
| Bone | CCAM (procedures) | NZFA007 | Transfemoral amputation | T_MCOaaA/CDC_ACT |
| Bone | CCAM (procedures) | NZQK001 | Unilateral or bilateral teleradiography of the entire lower limb, front view with bipodal support | T_MCOaaA/CDC_ACT |
| Bone | CCAM (procedures) | NZQK005 | Radiography of 2 segments of the lower limb | T_MCOaaA/CDC_ACT |
| Bone | CCAM (procedures) | NZQK006 | Radiography of 3 or more segments of lower limb | T_MCOaaA/CDC_ACT |
| Soft tissue | GHM (DRG) | 01C041 | Non-traumatic craniotomies, age over 17, level 1 | T_MCOaaB/GRC_GHM |
| Soft tissue | GHM (DRG) | 01C042 | Non-traumatic craniotomies, age over 17, level 2 | T_MCOaaB/GRC_GHM |
| Soft tissue | GHM (DRG) | 01C051 | Spine and cord procedures for neurological conditions, level 1 | T_MCOaaB/GRC_GHM |
| Soft tissue | GHM (DRG) | 01C052 | Spine and cord procedures for neurological conditions, level 2 | T_MCOaaB/GRC_GHM |
| Soft tissue | GHM (DRG) | 01C053 | Procedures on the spine and cord for neurological conditions, level 3 | T_MCOaaB/GRC_GHM |
| Soft tissue | GHM (DRG) | 01C08] | Interventions on the pre-cerebral vascular system, level 4 | T_MCOaaB/GRC_GHM |
| Soft tissue | GHM (DRG) | 01C081 | Interventions on cranial or peripheral nerves and other interventions on the nervous system, level 1 | T_MCOaaB/GRC_GHM |
| Soft tissue | GHM (DRG) | 01C082 | Interventions on cranial or peripheral nerves and other interventions on the nervous system, level 2 | T_MCOaaB/GRC_GHM |
| Soft tissue | GHM (DRG) | 01C083 | Interventions on cranial or peripheral nerves and other interventions on the nervous system, level 3 | T_MCOaaB/GRC_GHM |

| | | | | |
|---|---|---|---|---|
| Soft tissue | GHM (DRG) | 01C114 | Craniotomies for tumors, age under 18, level 4 | T_MCOaaB/GRC_GHM |
| Soft tissue | GHM (DRG) | 01C131 | Craniotomies for non-tumoral conditions, age under 18, level 4 | T_MCOaaB/GRC_GHM |
| Soft tissue | GHM (DRG) | 01C132 | Craniotomies for non-tumoral conditions, age under 18, level 4 | T_MCOaaB/GRC_GHM |
| Soft tissue | GHM (DRG) | 01C141 | Superficial nerve releases excluding median nerve at carpal tunnel, level 1 | T_MCOaaB/GRC_GHM |
| Soft tissue | GHM (DRG) | 01C142 | Superficial nerve releases excluding median at carpal tunnel, level 2 | T_MCOaaB/GRC_GHM |
| Soft tissue | GHM (DRG) | 01C14J | Superficial nerve releases excluding median to carpal tunnel, ambulatory | T_MCOaaB/GRC_GHM |
| Soft tissue | GHM (DRG) | 01C152 | Median to carpal tunnel releases, level 2 | T_MCOaaB/GRC_GHM |
| Soft tissue | GHM (DRG) | 01M27 | Other nervous system tumors | T_MCOaaB/GRC_GHM |
| Soft tissue | GHM (DRG) | 02C031 | Orbit procedures, level 1 | T_MCOaaB/GRC_GHM |
| Soft tissue | GHM (DRG) | 02C032 | Orbit procedures, level 2 | T_MCOaaB/GRC_GHM |
| Soft tissue | GHM (DRG) | 02C033 | Orbit procedures, level 3 | T_MCOaaB/GRC_GHM |
| Soft tissue | GHM (DRG) | 02C03J | Outpatient orbital procedures | T_MCOaaB/GRC_GHM |
| Soft tissue | GHM (DRG) | 03C061 | Sinus and mastoid process, age under 18, level 1 | T_MCOaaB/GRC_GHM |
| Soft tissue | GHM (DRG) | 03C071 | Sinus and mastoid process procedures, age over 17, level 1 | T_MCOaaB/GRC_GHM |
| Soft tissue | GHM (DRG) | 03C073 | Sinus and mastoid process procedures, age over 17, level 3 | T_MCOaaB/GRC_GHM |
| Soft tissue | GHM (DRG) | 03C132 | Procedures on tonsils and adenoids other than isolated tonsillectomies and/or adenoidectomies, age over 17, level 2 | T_MCOaaB/GRC_GHM |
| Soft tissue | GHM (DRG) | 03C171 | Mouth procedures, level 1 | T_MCOaaB/GRC_GHM |
| Soft tissue | GHM (DRG) | 03C17J | Oral procedures, outpatient | T_MCOaaB/GRC_GHM |
| Soft tissue | GHM (DRG) | 03C191 | Facial osteotomies, level 1 | T_MCOaaB/GRC_GHM |
| Soft tissue | GHM (DRG) | 03C241 | Salivary gland procedures, level 1 | T_MCOaaB/GRC_GHM |
| Soft tissue | GHM (DRG) | 03C243 | Salivary gland procedures, level 3 | T_MCOaaB/GRC_GHM |
| Soft tissue | GHM (DRG) | 03C251 | Major head and neck procedures, level 1 | T_MCOaaB/GRC_GHM |
| Soft tissue | GHM (DRG) | 03C253 | Major head and neck procedures, level 3 | T_MCOaaB/GRC_GHM |
| Soft tissue | GHM (DRG) | 03C254 | Major head and neck procedures, level 4 | T_MCOaaB/GRC_GHM |
| Soft tissue | GHM (DRG) | 03C261 | Other head and neck procedures, level 1 | T_MCOaaB/GRC_GHM |
| Soft tissue | GHM (DRG) | 03C262 | Other head and neck procedures, level 2 | T_MCOaaB/GRC_GHM |
| Soft tissue | GHM (DRG) | 03C263 | Other head and neck procedures, level 3 | T_MCOaaB/GRC_GHM |
| Soft tissue | GHM (DRG) | 03C264 | Other head and neck procedures, level 4 | T_MCOaaB/GRC_GHM |
| Soft tissue | GHM (DRG) | 03C291 | Other ear, nose or throat procedures for malignant tumors, level 1 | T_MCOaaB/GRC_GHM |
| Soft tissue | GHM (DRG) | 03C293 | Other ear, nose or throat procedures for malignant tumors, level 3 | T_MCOaaB/GRC_GHM |
| Soft tissue | GHM (DRG) | 03C29J | Other ear, nose or throat procedures for malignant tumors, outpatient | T_MCOaaB/GRC_GHM |
| Soft tissue | GHM (DRG) | 04C021 | Major thoracic procedures, level 1 | T_MCOaaB/GRC_GHM |
| Soft tissue | GHM (DRG) | 04C022 | Major thoracic procedures, level 2 | T_MCOaaB/GRC_GHM |
| Soft tissue | GHM (DRG) | 04C023 | Major thoracic procedures, level 3 | T_MCOaaB/GRC_GHM |
| Soft tissue | GHM (DRG) | 04C024 | Major thoracic procedures, level 4 | T_MCOaaB/GRC_GHM |
| Soft tissue | GHM (DRG) | 04C031 | Other surgical procedures on the respiratory system, level 1 | T_MCOaaB/GRC_GHM |
| Soft tissue | GHM (DRG) | 05C033 | Valve replacement surgery with extracorporeal circulation, without cardiac catheterization or coronary angiography, level 3 | T_MCOaaB/GRC_GHM |
| Soft tissue | GHM (DRG) | 05C052 | Coronary artery bypass grafts without cardiac catheterization or coronary angiography, level 2 | T_MCOaaB/GRC_GHM |
| Soft tissue | GHM (DRG) | 05C062 | Other cardiothoracic procedures, age over 1 year, or vascular procedures regardless of age, with extracorporeal circulation, level 2 | T_MCOaaB/GRC_GHM |
| Soft tissue | GHM (DRG) | 05C063 | Other cardiothoracic procedures, age over 1 year, or vascular procedures of any age, with extracorporeal circulation, level 3 | T_MCOaaB/GRC_GHM |
| Soft tissue | GHM (DRG) | 05C064 | Other cardiothoracic procedures, age over 1 year, or vascular procedures of any age, with extracorporeal circulation, level 4 | T_MCOaaB/GRC_GHM |
| Soft tissue | GHM (DRG) | 05C082 | Other cardiothoracic procedures, age over 1 year, or vascular procedures of any age, without extracorporeal circulation, level 2 | T_MCOaaB/GRC_GHM |
| Soft tissue | GHM (DRG) | 05C084 | Other cardiothoracic procedures, age over 1 year, or vascular procedures of any age, without extracorporeal circulation, level 4 | T_MCOaaB/GRC_GHM |
| Soft tissue | GHM (DRG) | 05C101 | Major revascularization surgery, level 1 | T_MCOaaB/GRC_GHM |
| Soft tissue | GHM (DRG) | 05C102 | Major revascularization surgery, level 2 | T_MCOaaB/GRC_GHM |
| Soft tissue | GHM (DRG) | 05C103 | Major revascularization surgery, level 3 | T_MCOaaB/GRC_GHM |
| Soft tissue | GHM (DRG) | 05C104 | Major revascularization surgery, level 4 | T_MCOaaB/GRC_GHM |
| Soft tissue | GHM (DRG) | 05C152 | Permanent pacemaker placement without acute myocardial infarction, congestive heart failure or shock, level 2 | T_MCOaaB/GRC_GHM |
| Soft tissue | GHM (DRG) | 05C171 | Vein ligatures and awakenings, level 1 | T_MCOaaB/GRC_GHM |
| Soft tissue | GHM (DRG) | 05C173 | Vein ligation and stripping, level 3 | T_MCOaaB/GRC_GHM |
| Soft tissue | GHM (DRG) | 05C181 | Other procedures on the circulatory system, level 1 | T_MCOaaB/GRC_GHM |
| Soft tissue | GHM (DRG) | 05C182 | Other circulatory procedures, level 2 | T_MCOaaB/GRC_GHM |
| Soft tissue | GHM (DRG) | 05C183 | Other circulatory procedures, level 3 | T_MCOaaB/GRC_GHM |

| | | | | |
|---|---|---|---|---|
| Soft tissue | GHM (DRG) | 05C184 | Other circulatory system procedures, level 4 | T_MCOaaB/GRC_GHM |
| Soft tissue | GHM (DRG) | 05C18J | Other outpatient circulatory procedures | T_MCOaaB/GRC_GHM |
| Soft tissue | GHM (DRG) | 06C031 | Rectal resections, level 1 | T_MCOaaB/GRC_GHM |
| Soft tissue | GHM (DRG) | 06C032 | Rectal resections, level 2 | T_MCOaaB/GRC_GHM |
| Soft tissue | GHM (DRG) | 06C033 | Rectal resections, level 3 | T_MCOaaB/GRC_GHM |
| Soft tissue | GHM (DRG) | 06C034 | Rectal resections, level 4 | T_MCOaaB/GRC_GHM |
| Soft tissue | GHM (DRG) | 06C041 | Major small bowel and colon procedures, level 1 | T_MCOaaB/GRC_GHM |
| Soft tissue | GHM (DRG) | 06C042 | Major small bowel and colon procedures, level 2 | T_MCOaaB/GRC_GHM |
| Soft tissue | GHM (DRG) | 06C043 | Major small bowel and colon procedures, level 3 | T_MCOaaB/GRC_GHM |
| Soft tissue | GHM (DRG) | 06C044 | Major small bowel and colon procedures, level 4 | T_MCOaaB/GRC_GHM |
| Soft tissue | GHM (DRG) | 06C093 | Uncomplicated appendectomies, level 3 | T_MCOaaB/GRC_GHM |
| Soft tissue | GHM (DRG) | 06C111 | Reconstructive hernia and ventricular surgery, under 18 years of age, outpatient | T_MCOaaB/GRC_GHM |
| Soft tissue | GHM (DRG) | 06C112 | Repair of hernias and ventricles, under 18 years of age, outpatient | T_MCOaaB/GRC_GHM |
| Soft tissue | GHM (DRG) | 06C121 | Inguinal and crural hernia repair, age over 17, level 1 | T_MCOaaB/GRC_GHM |
| Soft tissue | GHM (DRG) | 06C122 | Inguinal and crural hernia repair, age over 17, level 2 | T_MCOaaB/GRC_GHM |
| Soft tissue | GHM (DRG) | 06C12J | Reconstructive procedures for inguinal and crural hernias, age over 17, outpatient | T_MCOaaB/GRC_GHM |
| Soft tissue | GHM (DRG) | 06C151 | Other digestive tract procedures other than laparotomy, level 1 | T_MCOaaB/GRC_GHM |
| Soft tissue | GHM (DRG) | 06C152 | Other digestive tract procedures other than laparotomy, level 2 | T_MCOaaB/GRC_GHM |
| Soft tissue | GHM (DRG) | 06C153 | Other digestive tract procedures other than laparotomy, level 3 | T_MCOaaB/GRC_GHM |
| Soft tissue | GHM (DRG) | 06C154 | Other digestive tract procedures other than laparotomy, level 4 | T_MCOaaB/GRC_GHM |
| Soft tissue | GHM (DRG) | 06C161 | Procedures on the esophagus, stomach and duodenum for malignant tumors, age over 17, level 1 | T_MCOaaB/GRC_GHM |
| Soft tissue | GHM (DRG) | 06C162 | Procedures on the esophagus, stomach and duodenum for malignant tumors, age over 17, level 2 | T_MCOaaB/GRC_GHM |
| Soft tissue | GHM (DRG) | 06C163 | Procedures on the esophagus, stomach and duodenum for malignant tumors, age over 17, level 3 | T_MCOaaB/GRC_GHM |
| Soft tissue | GHM (DRG) | 06C164 | Operations on the esophagus, stomach and duodenum for malignant tumors, age over 17, level 4 | T_MCOaaB/GRC_GHM |
| Soft tissue | GHM (DRG) | 06C211 | Other operations on the digestive tract by laparotomy, level 1 | T_MCOaaB/GRC_GHM |
| Soft tissue | GHM (DRG) | 06C212 | Other laparotomy operations on the digestive tract, level 2 | T_MCOaaB/GRC_GHM |
| Soft tissue | GHM (DRG) | 06C213 | Other laparotomy digestive tract procedures, level 3 | T_MCOaaB/GRC_GHM |
| Soft tissue | GHM (DRG) | 06C214 | Other laparotomy digestive tract procedures, level 4 | T_MCOaaB/GRC_GHM |
| Soft tissue | GHM (DRG) | 06C25J | Reconstructive hernia procedures, excluding inguinal hernia, crural hernia, age over 17, outpatient | T_MCOaaB/GRC_GHM |
| Soft tissue | GHM (DRG) | 06M05 | Other malignant tumors of the digestive tract | T_MCOaaB/GRC_GHM |
| Soft tissue | GHM (DRG) | 07C091 | Procedures on the liver, pancreas, portal vein or vena cava for malignant tumors, level 1 | T_MCOaaB/GRC_GHM |
| Soft tissue | GHM (DRG) | 07C092 | Procedures on the liver, pancreas and portal or vena cava for malignant tumours, level 2 | T_MCOaaB/GRC_GHM |
| Soft tissue | GHM (DRG) | 07C093 | Procedures on the liver, pancreas and portal or vena cava for malignant tumors, level 3 | T_MCOaaB/GRC_GHM |
| Soft tissue | GHM (DRG) | 07C094 | Procedures on the liver, pancreas and portal or vena cava for malignant tumours, level 4 | T_MCOaaB/GRC_GHM |
| Soft tissue | GHM (DRG) | 08C022 | Multiple major knee and/or hip procedures, level 2 | T_MCOaaB/GRC_GHM |
| Soft tissue | GHM (DRG) | 08C023 | Multiple major knee and/or hip procedures, level 3 | T_MCOaaB/GRC_GHM |
| Soft tissue | GHM (DRG) | 08C024 | Multiple major knee and/or hip procedures, level 4 | T_MCOaaB/GRC_GHM |
| Soft tissue | GHM (DRG) | 08C041 | Hip and femur procedures, age under 18, level 1 | T_MCOaaB/GRC_GHM |
| Soft tissue | GHM (DRG) | 08C042 | Hip and femur procedures, age under 18, level 2 | T_MCOaaB/GRC_GHM |
| Soft tissue | GHM (DRG) | 08C043 | Hip and femur procedures, age under 18, level 3 | T_MCOaaB/GRC_GHM |
| Soft tissue | GHM (DRG) | 08C061 | Amputations for musculoskeletal and connective tissue disorders, level 1 | T_MCOaaB/GRC_GHM |
| Soft tissue | GHM (DRG) | 08C062 | Amputations for musculoskeletal and connective tissue disorders, level 2 | T_MCOaaB/GRC_GHM |
| Soft tissue | GHM (DRG) | 08C063 | Amputations for musculoskeletal and connective tissue disorders, level 3 | T_MCOaaB/GRC_GHM |
| Soft tissue | GHM (DRG) | 08C064 | Amputations for musculoskeletal and connective tissue disorders, level 4 | T_MCOaaB/GRC_GHM |
| Soft tissue | GHM (DRG) | 08C121 | Osteoarticular biopsies, level 1 | T_MCOaaB/GRC_GHM |
| Soft tissue | GHM (DRG) | 08C12J | Outpatient osteoarticular biopsies | T_MCOaaB/GRC_GHM |
| Soft tissue | GHM (DRG) | 08C134 | Localized bone resections and/or removal of internal fixation hardware in the hip and femur, level 4 | T_MCOaaB/GRC_GHM |
| Soft tissue | GHM (DRG) | 08C141 | Localized bone resections and/or removal of internal fixation hardware at sites other than the hip and femur, level 1 | T_MCOaaB/GRC_GHM |
| Soft tissue | GHM (DRG) | 08C201 | Skin grafts for musculoskeletal or connective tissue disease, level 1 | T_MCOaaB/GRC_GHM |
| Soft tissue | GHM (DRG) | 08C202 | Skin grafts for musculoskeletal or connective tissue disease, level 2 | T_MCOaaB/GRC_GHM |
| Soft tissue | GHM (DRG) | 08C203 | Skin grafts for musculoskeletal or connective tissue disease, level 3 | T_MCOaaB/GRC_GHM |
| Soft tissue | GHM (DRG) | 08C20J | Skin grafts for musculoskeletal or connective tissue disorders, outpatient | T_MCOaaB/GRC_GHM |
| Soft tissue | GHM (DRG) | 08C211 | Other musculoskeletal and connective tissue procedures, level 1 | T_MCOaaB/GRC_GHM |
| Soft tissue | GHM (DRG) | 08C212 | Other musculoskeletal and connective tissue procedures, level 2 | T_MCOaaB/GRC_GHM |
| Soft tissue | GHM (DRG) | 08C213 | Other musculoskeletal and connective tissue procedures, level 3 | T_MCOaaB/GRC_GHM |
| Soft tissue | GHM (DRG) | 08C214 | Other musculoskeletal and connective tissue procedures, level 4 | T_MCOaaB/GRC_GHM |

| | | | | |
|---|---|---|---|---|
| Soft tissue | GHM (DRG) | 08C221 | Joint replacement procedures, level 1 | T_MCOaaB/GRC_GHM |
| Soft tissue | GHM (DRG) | 08C222 | Joint replacement procedures, level 2 | T_MCOaaB/GRC_GHM |
| Soft tissue | GHM (DRG) | 08C223 | Joint replacement procedures, level 3 | T_MCOaaB/GRC_GHM |
| Soft tissue | GHM (DRG) | 08C224 | Revision joint replacement, level 4 | T_MCOaaB/GRC_GHM |
| Soft tissue | GHM (DRG) | 08C241 | Knee prostheses, level 1 | T_MCOaaB/GRC_GHM |
| Soft tissue | GHM (DRG) | 08C252 | Shoulder prostheses, level 2 | T_MCOaaB/GRC_GHM |
| Soft tissue | GHM (DRG) | 08C271 | Other spinal procedures, level 1 | T_MCOaaB/GRC_GHM |
| Soft tissue | GHM (DRG) | 08C272 | Other spinal procedures, level 2 | T_MCOaaB/GRC_GHM |
| Soft tissue | GHM (DRG) | 08C273 | Other spinal procedures, level 3 | T_MCOaaB/GRC_GHM |
| Soft tissue | GHM (DRG) | 08C274 | Other spinal procedures, level 4 | T_MCOaaB/GRC_GHM |
| Soft tissue | GHM (DRG) | 08C281 | Maxillofacial procedures, level 1 | T_MCOaaB/GRC_GHM |
| Soft tissue | GHM (DRG) | 08C282 | Maxillofacial procedures, level 2 | T_MCOaaB/GRC_GHM |
| Soft tissue | GHM (DRG) | 08C283 | Maxillofacial procedures, level 3 | T_MCOaaB/GRC_GHM |
| Soft tissue | GHM (DRG) | 08C284 | Maxillofacial procedures, level 4 | T_MCOaaB/GRC_GHM |
| Soft tissue | GHM (DRG) | 08C28J | Maxillofacial procedures, outpatient | T_MCOaaB/GRC_GHM |
| Soft tissue | GHM (DRG) | 08C291 | Soft tissue procedures for malignant tumors, level 1 | T_MCOaaB/GRC_GHM |
| Soft tissue | GHM (DRG) | 08C292 | Soft-tissue procedures for malignant tumors, level 2 | T_MCOaaB/GRC_GHM |
| Soft tissue | GHM (DRG) | 08C293 | Soft-tissue procedures for malignancies, level 3 | T_MCOaaB/GRC_GHM |
| Soft tissue | GHM (DRG) | 08C294 | Soft-tissue procedures for malignancy, level 4 | T_MCOaaB/GRC_GHM |
| Soft tissue | GHM (DRG) | 08C29J | Outpatient soft-tissue procedures for malignant tumors | T_MCOaaB/GRC_GHM |
| Soft tissue | GHM (DRG) | 08C311 | Leg procedures, age under 18, level 1 | T_MCOaaB/GRC_GHM |
| Soft tissue | GHM (DRG) | 08C321 | Leg procedures, age greater than 17, level 1 | T_MCOaaB/GRC_GHM |
| Soft tissue | GHM (DRG) | 08C322 | Leg procedures, age greater than 17, level 2 | T_MCOaaB/GRC_GHM |
| Soft tissue | GHM (DRG) | 08C323 | Leg procedures, age greater than 17, level 3 | T_MCOaaB/GRC_GHM |
| Soft tissue | GHM (DRG) | 08C331 | Ankle and hindfoot procedures excluding fractures, level 1 | T_MCOaaB/GRC_GHM |
| Soft tissue | GHM (DRG) | 08C332 | Procedures on the ankle and hindfoot excluding fractures, level 2 | T_MCOaaB/GRC_GHM |
| Soft tissue | GHM (DRG) | 08C351 | Procedures on the arm, elbow and shoulder, level 1 | T_MCOaaB/GRC_GHM |
| Soft tissue | GHM (DRG) | 08C352 | Procedures on the arm, elbow and shoulder, level 2 | T_MCOaaB/GRC_GHM |
| Soft tissue | GHM (DRG) | 08C35J | Arm, elbow and shoulder procedures, outpatient | T_MCOaaB/GRC_GHM |
| Soft tissue | GHM (DRG) | 08C361 | Foot procedures, age under 18, level 1 | T_MCOaaB/GRC_GHM |
| Soft tissue | GHM (DRG) | 08C371 | Foot procedures, age over 17, level 1 | T_MCOaaB/GRC_GHM |
| Soft tissue | GHM (DRG) | 08C373 | Foot procedures, age over 17, level 3 | T_MCOaaB/GRC_GHM |
| Soft tissue | GHM (DRG) | 08C37J | Foot procedures, age over 17, outpatient | T_MCOaaB/GRC_GHM |
| Soft tissue | GHM (DRG) | 08C38J | Other knee arthroscopies, outpatient | T_MCOaaB/GRC_GHM |
| Soft tissue | GHM (DRG) | 08C391 | Forearm procedures, level 1 | T_MCOaaB/GRC_GHM |
| Soft tissue | GHM (DRG) | 08C393 | Forearm procedures, level 3 | T_MCOaaB/GRC_GHM |
| Soft tissue | GHM (DRG) | 08C40] | Forearm procedures, outpatient | T_MCOaaB/GRC_GHM |
| Soft tissue | GHM (DRG) | 08C41J | Arthroscopies of other locations, outpatient | T_MCOaaB/GRC_GHM |
| Soft tissue | GHM (DRG) | 08C421 | Non-minor soft-tissue procedures, level 1 | T_MCOaaB/GRC_GHM |
| Soft tissue | GHM (DRG) | 08C422 | Non-minor soft-tissue procedures, level 2 | T_MCOaaB/GRC_GHM |
| Soft tissue | GHM (DRG) | 08C424 | Non-minor soft-tissue procedures, level 4 | T_MCOaaB/GRC_GHM |
| Soft tissue | GHM (DRG) | 08C42J | Outpatient non-minor soft-tissue procedures | T_MCOaaB/GRC_GHM |
| Soft tissue | GHM (DRG) | 08C431 | Non-minor hand procedures, level 1 | T_MCOaaB/GRC_GHM |
| Soft tissue | GHM (DRG) | 08C432 | Non-minor hand procedures, level 2 | T_MCOaaB/GRC_GHM |
| Soft tissue | GHM (DRG) | 08C43J | Outpatient non-minor hand procedures | T_MCOaaB/GRC_GHM |
| Soft tissue | GHM (DRG) | 08C441 | Other hand procedures, level 1 | T_MCOaaB/GRC_GHM |
| Soft tissue | GHM (DRG) | 08C44J | Other hand procedures, outpatient | T_MCOaaB/GRC_GHM |
| Soft tissue | GHM (DRG) | 08C45J | Arthroscopic menisectomy, outpatient | T_MCOaaB/GRC_GHM |
| Soft tissue | GHM (DRG) | 08C46] | Arthroscopic menisectomy, outpatient | T_MCOaaB/GRC_GHM |
| Soft tissue | GHM (DRG) | 08C461 | Other soft-tissue procedures, level 1 | T_MCOaaB/GRC_GHM |
| Soft tissue | GHM (DRG) | 08C462 | Other soft-tissue procedures, level 2 | T_MCOaaB/GRC_GHM |
| Soft tissue | GHM (DRG) | 08C463 | Other soft-tissue procedures, level 3 | T_MCOaaB/GRC_GHM |
| Soft tissue | GHM (DRG) | 08C481 | Hip prostheses for conditions other than recent trauma, level 1 | T_MCOaaB/GRC_GHM |
| Soft tissue | GHM (DRG) | 08C483 | Hip prostheses for conditions other than recent trauma, level 3 | T_MCOaaB/GRC_GHM |
| Soft tissue | GHM (DRG) | 08C484 | Hip prostheses for conditions other than recent trauma, level 4 | T_MCOaaB/GRC_GHM |
| Soft tissue | GHM (DRG) | 08C501 | Hip and femur procedures excluding recent trauma, age over 17, level 1 | T_MCOaaB/GRC_GHM |

| | | | | |
|---|---|---|---|---|
| Soft tissue | GHM (DRG) | 08C502 | Hip and femur procedures excluding recent trauma, age over 17, level 2 | T_MCOaaB/GRC_GHM |
| Soft tissue | GHM (DRG) | 08C503 | Hip and femur procedures, excluding recent trauma, age over 17, level 3 | T_MCOaaB/GRC_GHM |
| Soft tissue | GHM (DRG) | 08C504 | Hip and femur procedures, excluding recent trauma, age over 17, level 4 | T_MCOaaB/GRC_GHM |
| Soft tissue | GHM (DRG) | 08C521 | Other major spinal procedures, level 1 | T_MCOaaB/GRC_GHM |
| Soft tissue | GHM (DRG) | 08C522 | Other major spine procedures, level 2 | T_MCOaaB/GRC_GHM |
| Soft tissue | GHM (DRG) | 08C523 | Other major spine procedures, level 3 | T_MCOaaB/GRC_GHM |
| Soft tissue | GHM (DRG) | 08C524 | Other major spine procedures, level 4 | T_MCOaaB/GRC_GHM |
| Soft tissue | GHM (DRG) | 08C541 | Knee procedures for non-traumatic conditions, level 1 | T_MCOaaB/GRC_GHM |
| Soft tissue | GHM (DRG) | 08C543 | Knee procedures for non-traumatic conditions, level 3 | T_MCOaaB/GRC_GHM |
| Soft tissue | GHM (DRG) | 08C544 | Knee procedures for non-traumatic conditions, level 4 | T_MCOaaB/GRC_GHM |
| Soft tissue | GHM (DRG) | 08C54J | Outpatient knee procedures for non-traumatic conditions | T_MCOaaB/GRC_GHM |
| Soft tissue | GHM (DRG) | 08C591 | Wrist tenosynovectomies, level 1 | T_MCOaaB/GRC_GHM |
| Soft tissue | GHM (DRG) | 08C59J | Outpatient wrist tenosynovectomies | T_MCOaaB/GRC_GHM |
| Soft tissue | GHM (DRG) | 08C601 | Wrist procedures other than tenosynovectomies, level 1 | T_MCOaaB/GRC_GHM |
| Soft tissue | GHM (DRG) | 09C02J | | T_MCOaaB/GRC_GHM |
| Soft tissue | GHM (DRG) | 09C031 | Skin grafts and/or wound trimming, excluding skin ulcers and cellulitis, level 1 | T_MCOaaB/GRC_GHM |
| Soft tissue | GHM (DRG) | 09C032 | Skin grafts and/or wound trimming, excluding skin ulcers and cellulitis, level 2 | T_MCOaaB/GRC_GHM |
| Soft tissue | GHM (DRG) | 09C034 | Skin grafts and/or wound trimming, excluding skin ulcers and cellulitis, level 4 | T_MCOaaB/GRC_GHM |
| Soft tissue | GHM (DRG) | 09C03J | Greffes de peau et/ou parages de plaie à l'exception des ulcères cutanés et cellulites, en ambulatoire | T_MCOaaB/GRC_GHM |
| Soft tissue | GHM (DRG) | 09C041 | Total mastectomy for malignancy, level 1 | T_MCOaaB/GRC_GHM |
| Soft tissue | GHM (DRG) | 09C042 | Total mastectomy for malignancy, level 2 | T_MCOaaB/GRC_GHM |
| Soft tissue | GHM (DRG) | 09C043 | Total mastectomy for malignancy, level 3 | T_MCOaaB/GRC_GHM |
| Soft tissue | GHM (DRG) | 09C044 | Total mastectomy for malignancy, level 4 | T_MCOaaB/GRC_GHM |
| Soft tissue | GHM (DRG) | 09C051 | Subtotal mastectomies for malignancy, level 1 | T_MCOaaB/GRC_GHM |
| Soft tissue | GHM (DRG) | 09C052 | Subtotal mastectomies for malignancy, level 2 | T_MCOaaB/GRC_GHM |
| Soft tissue | GHM (DRG) | 09C05J | Subtotal mastectomies for malignancy, outpatient | T_MCOaaB/GRC_GHM |
| Soft tissue | GHM (DRG) | 09C061 | Breast procedures for non-malignant conditions other than biopsy and local excision procedures, level 1 | T_MCOaaB/GRC_GHM |
| Soft tissue | GHM (DRG) | 09C062 | Breast procedures for non-malignant conditions other than biopsy and local excision procedures, level 2 | T_MCOaaB/GRC_GHM |
| Soft tissue | GHM (DRG) | 09C06T | Breast procedures for non-malignant conditions other than biopsies and local excisions, very short duration | T_MCOaaB/GRC_GHM |
| Soft tissue | GHM (DRG) | 09C071 | Biopsies and local excisions for non-malignant breast conditions, level 1 | T_MCOaaB/GRC_GHM |
| Soft tissue | GHM (DRG) | 09C07J | Outpatient biopsies and local excisions for non-malignant breast diseases | T_MCOaaB/GRC_GHM |
| Soft tissue | GHM (DRG) | 09C091 | Plastic procedures other than cosmetic surgery, level 1 | T_MCOaaB/GRC_GHM |
| Soft tissue | GHM (DRG) | 09C092 | Plastic procedures other than cosmetic surgery, level 2 | T_MCOaaB/GRC_GHM |
| Soft tissue | GHM (DRG) | 09C09J | Plastic procedures other than cosmetic surgery, level 4 | T_MCOaaB/GRC_GHM |
| Soft tissue | GHM (DRG) | 09C101 | Other skin, subcutaneous tissue or breast procedures, level 1 | T_MCOaaB/GRC_GHM |
| Soft tissue | GHM (DRG) | 09C102 | Other skin, subcutaneous tissue or breast procedures, level 2 | T_MCOaaB/GRC_GHM |
| Soft tissue | GHM (DRG) | 09C103 | Other skin, subcutaneous tissue or breast procedures, level 3 | T_MCOaaB/GRC_GHM |
| Soft tissue | GHM (DRG) | 09C10J | Other skin, subcutaneous tissue or breast procedures, outpatient | T_MCOaaB/GRC_GHM |
| Soft tissue | GHM (DRG) | 09C111 | Breast reconstruction, level 1 | T_MCOaaB/GRC_GHM |
| Soft tissue | GHM (DRG) | 09C112 | Breast reconstruction, level 2 | T_MCOaaB/GRC_GHM |
| Soft tissue | GHM (DRG) | 09C113 | Breast reconstruction, level 3 | T_MCOaaB/GRC_GHM |
| Soft tissue | GHM (DRG) | 09C114 | Breast reconstruction, level 4 | T_MCOaaB/GRC_GHM |
| Soft tissue | GHM (DRG) | 09C121 | Cyst, granuloma and nail procedures, level 1 | T_MCOaaB/GRC_GHM |
| Soft tissue | GHM (DRG) | 09C123 | Cyst, granuloma and nail procedures, level 3 | T_MCOaaB/GRC_GHM |
| Soft tissue | GHM (DRG) | 09C141 | Certain lymph node treatments for skin, subcutaneous tissue or breast disorders, level 1 | T_MCOaaB/GRC_GHM |
| Soft tissue | GHM (DRG) | 09C143 | Certain lymphonodal treatments for skin, subcutaneous tissue or breast disorders, level 3 | T_MCOaaB/GRC_GHM |
| Soft tissue | GHM (DRG) | 09C151 | Procedures on the skin, subcutaneous tissue or breasts for traumatic lesions, level 1 | T_MCOaaB/GRC_GHM |
| Soft tissue | GHM (DRG) | 09C152 | Procedures on the skin, subcutaneous tissue or breasts for traumatic lesions, level 2 | T_MCOaaB/GRC_GHM |
| Soft tissue | GHM (DRG) | 09C153 | Procedures on the skin, subcutaneous tissue or breasts for traumatic lesions, level 3 | T_MCOaaB/GRC_GHM |
| Soft tissue | GHM (DRG) | 10C031 | Adrenal gland procedures, level 1 | T_MCOaaB/GRC_GHM |
| Soft tissue | GHM (DRG) | 10C032 | Adrenal gland procedures, level 2 | T_MCOaaB/GRC_GHM |
| Soft tissue | GHM (DRG) | 10C033 | Adrenal gland procedures, level 3 | T_MCOaaB/GRC_GHM |
| Soft tissue | GHM (DRG) | 10C034 | Adrenal gland procedures, level 4 | T_MCOaaB/GRC_GHM |
| Soft tissue | GHM (DRG) | 10C081 | Other procedures for endocrine, metabolic or nutritional disorders, level 1 | T_MCOaaB/GRC_GHM |
| Soft tissue | GHM (DRG) | 10C083 | Other procedures for endocrine, metabolic or nutritional disorders, level 3 | T_MCOaaB/GRC_GHM |
| Soft tissue | GHM (DRG) | 10C111 | Thyroid procedures for malignant tumors, level 1 | T_MCOaaB/GRC_GHM |

| | | | | |
|---|---|---|---|---|
| Soft tissue | GHM (DRG) | 11C021 | Kidney and ureter procedures and major bladder surgery for tumor disease, level 1 | T_MCOaaB/GRC_GHM |
| Soft tissue | GHM (DRG) | 11C022 | Kidney and ureter procedures and major bladder surgery for tumor disease, level 2 | T_MCOaaB/GRC_GHM |
| Soft tissue | GHM (DRG) | 11C023 | Kidney and ureter procedures and major bladder surgery for tumor disease, level 3 | T_MCOaaB/GRC_GHM |
| Soft tissue | GHM (DRG) | 11C024 | Kidney and ureter procedures and major bladder surgery for tumoral disease, level 4 | T_MCOaaB/GRC_GHM |
| Soft tissue | GHM (DRG) | 11C032 | Kidney and ureter procedures and major bladder surgery for non-tumoral conditions, level 2 | T_MCOaaB/GRC_GHM |
| Soft tissue | GHM (DRG) | 11C043 | Other bladder procedures excluding transurethral procedures, level 3 | T_MCOaaB/GRC_GHM |
| Soft tissue | GHM (DRG) | 11C082 | Other kidney and urinary tract procedures, level 2 | T_MCOaaB/GRC_GHM |
| Soft tissue | GHM (DRG) | 11C083 | Other kidney and urinary tract procedures, level 3 | T_MCOaaB/GRC_GHM |
| Soft tissue | GHM (DRG) | 11C132 | Transurethral or transcutaneous procedures for non-lithiasis, level 2 | T_MCOaaB/GRC_GHM |
| Soft tissue | GHM (DRG) | 11C133 | Transurethral or transcutaneous procedures for non-lithiasis, level 3 | T_MCOaaB/GRC_GHM |
| Soft tissue | GHM (DRG) | 12C031 | Penile procedures, level 1 | T_MCOaaB/GRC_GHM |
| Soft tissue | GHM (DRG) | 12C051 | Testicular procedures for malignant tumors, level 1 | T_MCOaaB/GRC_GHM |
| Soft tissue | GHM (DRG) | 12C052 | Testicular procedures for malignant tumors, level 2 | T_MCOaaB/GRC_GHM |
| Soft tissue | GHM (DRG) | 12C053 | Testicular procedures for malignant tumors, level 3 | T_MCOaaB/GRC_GHM |
| Soft tissue | GHM (DRG) | 12C071 | Testicular procedures for non-malignant conditions, age over 17, level 1 | T_MCOaaB/GRC_GHM |
| Soft tissue | GHM (DRG) | 12C073 | Testicular procedures for non-malignant conditions, age over 17, level 3 | T_MCOaaB/GRC_GHM |
| Soft tissue | GHM (DRG) | 12C07J | Testicular procedures for non-malignant conditions, age over 17, outpatient | T_MCOaaB/GRC_GHM |
| Soft tissue | GHM (DRG) | 12C091 | Other procedures for malignant tumors of the male genital tract, level 1 | T_MCOaaB/GRC_GHM |
| Soft tissue | GHM (DRG) | 12C111 | Major male pelvic procedures for malignancies, level 1 | T_MCOaaB/GRC_GHM |
| Soft tissue | GHM (DRG) | 12C112 | Major male pelvic surgery for malignancy, level 2 | T_MCOaaB/GRC_GHM |
| Soft tissue | GHM (DRG) | 13C031 | Hysterectomies, level 1 | T_MCOaaB/GRC_GHM |
| Soft tissue | GHM (DRG) | 13C032 | Hysterectomies, level 2 | T_MCOaaB/GRC_GHM |
| Soft tissue | GHM (DRG) | 13C051 | Uteroannexal procedures for malignant tumors, level 1 | T_MCOaaB/GRC_GHM |
| Soft tissue | GHM (DRG) | 13C052 | Uteroannexal procedures for malignant tumors, level 2 | T_MCOaaB/GRC_GHM |
| Soft tissue | GHM (DRG) | 13C053 | Uteroannexal procedures for malignant tumors, level 3 | T_MCOaaB/GRC_GHM |
| Soft tissue | GHM (DRG) | 13C054 | Uteroannexal procedures for malignant tumors, level 4 | T_MCOaaB/GRC_GHM |
| Soft tissue | GHM (DRG) | 13C081 | Procedures on the vulva, vagina or cervix, level 1 | T_MCOaaB/GRC_GHM |
| Soft tissue | GHM (DRG) | 13C08J | Outpatient vulvar, vaginal or cervical procedures | T_MCOaaB/GRC_GHM |
| Soft tissue | GHM (DRG) | 13C133 | Other female genital tract procedures, level 3 | T_MCOaaB/GRC_GHM |
| Soft tissue | GHM (DRG) | 13C141 | Pelvic exenteration, extended hysterectomy or vulvectomy for malignant tumors, level 1 | T_MCOaaB/GRC_GHM |
| Soft tissue | GHM (DRG) | 13C142 | Pelvic exenteration, extended hysterectomy or vulvectomy for malignant tumors, level 2 | T_MCOaaB/GRC_GHM |
| Soft tissue | GHM (DRG) | 13C143 | Pelvic exenteration, extended hysterectomy or vulvectomy for malignant tumors, level 3 | T_MCOaaB/GRC_GHM |
| Soft tissue | GHM (DRG) | 13C144 | Pelvic exenteration, extended hysterectomy or vulvectomy for malignant tumors, level 4 | T_MCOaaB/GRC_GHM |
| Soft tissue | GHM (DRG) | 13C152 | Pelvic exenteration, enlarged hysterectomy or vulvectomy for non-malignant conditions, level 2 | T_MCOaaB/GRC_GHM |
| Soft tissue | GHM (DRG) | 13M07 | Other tumors of the female genital tract | T_MCOaaB/GRC_GHM |
| Soft tissue | GHM (DRG) | 16C031 | Other procedures for disorders of the blood and blood-forming organs, level 1 | T_MCOaaB/GRC_GHM |
| Soft tissue | GHM (DRG) | 17C021 | Major surgery for lymphoma or leukemia, level 1 | T_MCOaaB/GRC_GHM |
| Soft tissue | GHM (DRG) | 17C022 | Major surgery for lymphoma or leukemia, level 2 | T_MCOaaB/GRC_GHM |
| Soft tissue | GHM (DRG) | 17C031 | | T_MCOaaB/GRC_GHM |
| Soft tissue | GHM (DRG) | 17C03J | | T_MCOaaB/GRC_GHM |
| Soft tissue | GHM (DRG) | 17C041 | Major procedures for myeloproliferative disorders or tumors of unclear or diffuse location, level 1 | T_MCOaaB/GRC_GHM |
| Soft tissue | GHM (DRG) | 17C042 | Major procedures for myeloproliferative disorders or tumors of imprecise or diffuse location, level 2 | T_MCOaaB/GRC_GHM |
| Soft tissue | GHM (DRG) | 17C043 | Major procedures for myeloproliferative disorders or tumors of imprecise or diffuse site, level 3 | T_MCOaaB/GRC_GHM |
| Soft tissue | GHM (DRG) | 17C044 | Major procedures for myeloproliferative disorders or tumors of unclear or diffuse location, level 4 | T_MCOaaB/GRC_GHM |
| Soft tissue | GHM (DRG) | 17C051 | Other procedures for myeloproliferative disorders or tumors of unclear or diffuse location, level 1 | T_MCOaaB/GRC_GHM |
| Soft tissue | GHM (DRG) | 17C052 | Other procedures for myeloproliferative disorders or tumors of unclear or diffuse site, level 2 | T_MCOaaB/GRC_GHM |
| Soft tissue | GHM (DRG) | 17C053 | Other procedures for myeloproliferative disorders or tumors of unclear or diffuse site, level 3 | T_MCOaaB/GRC_GHM |
| Soft tissue | GHM (DRG) | 17C054 | Other procedures for myeloproliferative disorders or tumors of unclear or diffuse site, level 4 | T_MCOaaB/GRC_GHM |
| Soft tissue | GHM (DRG) | 17C05J | Other outpatient procedures for myeloproliferative disorders or tumors of unclear or diffuse location | T_MCOaaB/GRC_GHM |
| Soft tissue | GHM (DRG) | 17C061 | Major procedures in CMD17 (Myeloproliferative disorders and tumors of unclear or diffuse location.), level 1 | T_MCOaaB/GRC_GHM |
| Soft tissue | GHM (DRG) | 17C062 | Major procedures in CMD17 (Myeloproliferative disorders and tumors of unclear or diffuse location.), level 2 | T_MCOaaB/GRC_GHM |
| Soft tissue | GHM (DRG) | 17C063 | Major procedures in CMD17 (Myeloproliferative disorders and tumors of unclear or diffuse location.), level 3 | T_MCOaaB/GRC_GHM |
| Soft tissue | GHM (DRG) | 17C064 | Major procedures in CMD17 (Myeloproliferative disorders and tumors of unclear or diffuse location.), level 4 | T_MCOaaB/GRC_GHM |
| Soft tissue | GHM (DRG) | 17C071 | Intermediate procedures of CMD17 (Myeloproliferative disorders and tumors of unclear or diffuse location.), level 1 | T_MCOaaB/GRC_GHM |
| Soft tissue | GHM (DRG) | 17C072 | Intermediate procedures of CMD17 (Myeloproliferative disorders and tumors of unclear or diffuse location.), level 2 | T_MCOaaB/GRC_GHM |
| Soft tissue | GHM (DRG) | 17C073 | Intermediate procedures of CMD17 (Myeloproliferative disorders and tumors of unclear or diffuse location.), level 3 | T_MCOaaB/GRC_GHM |

| | | | | |
|---|---|---|---|---|
| Soft tissue | GHM (DRG) | 17C074 | Intermediate procedures of CMD17 (Myeloproliferative disorders and tumors of unclear or diffuse location.), level 4 | T_MCOaaB/GRC_GHM |
| Soft tissue | GHM (DRG) | 17C08] | Minor outpatient CMD17 (Myeloproliferative disorders and tumors of unclear or diffuse location.) procedures | T_MCOaaB/GRC_GHM |
| Soft tissue | GHM (DRG) | 17C081 | Minor interventions for CMD17 (Myeloproliferative disorders and tumors of unclear or diffuse location.), level 1 | T_MCOaaB/GRC_GHM |
| Soft tissue | GHM (DRG) | 17C082 | Minor interventions for CMD17 (Myeloproliferative disorders and tumors of unclear or diffuse location.), level 2 | T_MCOaaB/GRC_GHM |
| Soft tissue | GHM (DRG) | 17C083 | Minor interventions for CMD17 (Myeloproliferative disorders and tumors of unclear or diffuse location.), level 3 | T_MCOaaB/GRC_GHM |
| Soft tissue | GHM (DRG) | 17C084 | Minor interventions for CMD17 (Myeloproliferative disorders and tumors of unclear or diffuse location.), level 4 | T_MCOaaB/GRC_GHM |
| Soft tissue | GHM (DRG) | 17M17 | Other conditions and tumors of imprecise or diffuse site | T_MCOaaB/GRC_GHM |
| Soft tissue | GHM (DRG) | 19C021 | Surgical procedures with a principal diagnosis of mental illness, level 1 | T_MCOaaB/GRC_GHM |
| Soft tissue | GHM (DRG) | 21C051 | Other interventions for injuries or procedural complications, level 1 | T_MCOaaB/GRC_GHM |
| Soft tissue | GHM (DRG) | 21C052 | Other procedures for injuries or procedural complications, level 2 | T_MCOaaB/GRC_GHM |
| Soft tissue | GHM (DRG) | 21C064 | Skin grafts or wound dressings for lesions other than burns, level 4 | T_MCOaaB/GRC_GHM |
| Soft tissue | GHM (DRG) | 23C023 | Surgical procedures with other reasons for healthcare use, level 3 | T_MCOaaB/GRC_GHM |
| Soft tissue | CIM-10 (ICD10) | C07 | Malignant neoplasm of parotid gland | T_MCOaaB/DGN_PAL |
| Soft tissue | CIM-10 (ICD10) | C080 | Malignant neoplasm of submandibular gland | T_MCOaaB/DGN_PAL |
| Soft tissue | CIM-10 (ICD10) | C169 | Malignant neoplasm of stomach, unspecified | T_MCOaaB/DGN_PAL |
| Soft tissue | CIM-10 (ICD10) | C170 | Malignant neoplasm of duodenum | T_MCOaaB/DGN_PAL |
| Soft tissue | CIM-10 (ICD10) | C171 | Malignant neoplasm of jejunum | T_MCOaaB/DGN_PAL |
| Soft tissue | CIM-10 (ICD10) | C179 | Malignant neoplasm of small intestine, unspecified | T_MCOaaB/DGN_PAL |
| Soft tissue | CIM-10 (ICD10) | C187 | Malignant neoplasm of sigmoid colon | T_MCOaaB/DGN_PAL |
| Soft tissue | CIM-10 (ICD10) | C20 | Malignant neoplasm of rectum | T_MCOaaB/DGN_PAL |
| Soft tissue | CIM-10 (ICD10) | C300 | Malignant neoplasm of nasal cavity | T_MCOaaB/DGN_PAL |
| Soft tissue | CIM-10 (ICD10) | C310 | Malignant neoplasm of maxillary sinus | T_MCOaaB/DGN_PAL |
| Soft tissue | CIM-10 (ICD10) | C311 | Malignant neoplasm of ethmoidal sinus | T_MCOaaB/DGN_PAL |
| Soft tissue | CIM-10 (ICD10) | C341 | Malignant tumor of the upper lobe, bronchi or lung | T_MCOaaB/DGN_PAL |
| Soft tissue | CIM-10 (ICD10) | C348 | Malignant tumor of the upper lobe, bronchi or lung | T_MCOaaB/DGN_PAL |
| Soft tissue | CIM-10 (ICD10) | C349 | Malignant tumor of bronchus or lung, unspecified | T_MCOaaB/DGN_PAL |
| Soft tissue | CIM-10 (ICD10) | C37 | Malignant neoplasm of thymus | T_MCOaaB/DGN_PAL |
| Soft tissue | CIM-10 (ICD10) | C380 | Malignant neoplasm of heart | T_MCOaaB/DGN_PAL |
| Soft tissue | CIM-10 (ICD10) | C381 | Malignant neoplasm of anterior mediastinum | T_MCOaaB/DGN_PAL |
| Soft tissue | CIM-10 (ICD10) | C383 | Malignant neoplasm of mediastinum, part unspecified | T_MCOaaB/DGN_PAL |
| Soft tissue | CIM-10 (ICD10) | C384 | Malignant neoplasm of pleura | T_MCOaaB/DGN_PAL |
| Soft tissue | CIM-10 (ICD10) | C400 | Malignant tumor of the scapula and long bones of the upper limb | T_MCOaaB/DGN_PAL |
| Soft tissue | CIM-10 (ICD10) | C401 | Malignant tumor of the short bones of the upper limb | T_MCOaaB/DGN_PAL |
| Soft tissue | CIM-10 (ICD10) | C402 | Malignant tumor of the long bones of the lower limb | T_MCOaaB/DGN_PAL |
| Soft tissue | CIM-10 (ICD10) | C403 | Malignant tumor of the short bones of the lower limb | T_MCOaaB/DGN_PAL |
| Soft tissue | CIM-10 (ICD10) | C408 | Malignant tumor of contiguous bone and articular cartilage of the limbs | T_MCOaaB/DGN_PAL |
| Soft tissue | CIM-10 (ICD10) | C410 | Malignant neoplasm of bones of skull and face | T_MCOaaB/DGN_PAL |
| Soft tissue | CIM-10 (ICD10) | C412 | Malignant neoplasm of vertebral column | T_MCOaaB/DGN_PAL |
| Soft tissue | CIM-10 (ICD10) | C413 | Malignant neoplasm of ribs, sternum and clavicle | T_MCOaaB/DGN_PAL |
| Soft tissue | CIM-10 (ICD10) | C414 | Malignant neoplasm of pelvic bones, sacrum and coccyx | T_MCOaaB/DGN_PAL |
| Soft tissue | CIM-10 (ICD10) | C418 | Malignant tumor with contiguous localization of bone and articular cartilage | T_MCOaaB/DGN_PAL |
| Soft tissue | CIM-10 (ICD10) | C419 | Malignant neoplasm of bone and articular cartilage, unsp | T_MCOaaB/DGN_PAL |
| Soft tissue | CIM-10 (ICD10) | C442 | Malignant tumor of the skin of the ear and external auditory canal | T_MCOaaB/DGN_PAL |
| Soft tissue | CIM-10 (ICD10) | C443 | Malignant tumor of the skin of the face, other and unspecified parts | T_MCOaaB/DGN_PAL |
| Soft tissue | CIM-10 (ICD10) | C444 | Malignant skin tumor of the scalp and neck | T_MCOaaB/DGN_PAL |
| Soft tissue | CIM-10 (ICD10) | C445 | Malignant tumor of the skin of the trunk | T_MCOaaB/DGN_PAL |
| Soft tissue | CIM-10 (ICD10) | C446 | Tumeur maligne de la peau du membre supérieur, y compris l'épaule | T_MCOaaB/DGN_PAL |
| Soft tissue | CIM-10 (ICD10) | C447 | Malignant skin tumour of the lower limb, including the hip | T_MCOaaB/DGN_PAL |
| Soft tissue | CIM-10 (ICD10) | C460 | Kaposi's sarcoma of skin | T_MCOaaB/DGN_PAL |
| Soft tissue | CIM-10 (ICD10) | C461 | Kaposi's sarcoma of soft tissue | T_MCOaaB/DGN_PAL |
| Soft tissue | CIM-10 (ICD10) | C470 | Malignant neoplasm of prph nerves of head, face and neck | T_MCOaaB/DGN_PAL |
| Soft tissue | CIM-10 (ICD10) | C471 | Malignant tumor of the peripheral nerves of the upper limb, including the shoulder | T_MCOaaB/DGN_PAL |
| Soft tissue | CIM-10 (ICD10) | C472 | Malignant tumor of the peripheral nerves of the lower limb, including the hip | T_MCOaaB/DGN_PAL |
| Soft tissue | CIM-10 (ICD10) | C473 | Malignant neoplasm of peripheral nerves of thorax | T_MCOaaB/DGN_PAL |
| Soft tissue | CIM-10 (ICD10) | C474 | Malignant neoplasm of peripheral nerves of abdomen | T_MCOaaB/DGN_PAL |
| Soft tissue | CIM-10 (ICD10) | C475 | Malignant neoplasm of peripheral nerves of pelvis | T_MCOaaB/DGN_PAL |

| | | | | |
|---|---|---|---|---|
| Soft tissue | CIM-10 (ICD10) | C476 | Malignant neoplasm of peripheral nerves of trunk, unsp | T_MCOaaB/DGN_PAL |
| Soft tissue | CIM-10 (ICD10) | C480 | Malignant neoplasm of retroperitoneum | T_MCOaaB/DGN_PAL |
| Soft tissue | CIM-10 (ICD10) | C481 | Malignant neoplasm of specified parts of peritoneum | T_MCOaaB/DGN_PAL |
| Soft tissue | CIM-10 (ICD10) | C482 | Malignant neoplasm of peritoneum, unspecified | T_MCOaaB/DGN_PAL |
| Soft tissue | CIM-10 (ICD10) | C488 | Malig neoplasm of ovrlp sites of retroperiton and peritoneum | T_MCOaaB/DGN_PAL |
| Soft tissue | CIM-10 (ICD10) | C490 | Malig neoplm of conn and soft tissue of head, face and neck | T_MCOaaB/DGN_PAL |
| Soft tissue | CIM-10 (ICD10) | C491 | Malignant tumors of connective tissue and other soft tissues of the upper limb, including the shoulder | T_MCOaaB/DGN_PAL |
| Soft tissue | CIM-10 (ICD10) | C492 | Malignant tumor of the connective tissue and other soft tissues of the lower limb, including the hip | T_MCOaaB/DGN_PAL |
| Soft tissue | CIM-10 (ICD10) | C4930 | Malignant tumor of the vessels of the thorax | T_MCOaaB/DGN_PAL |
| Soft tissue | CIM-10 (ICD10) | C4938 | Other malignant tumors of connective tissue and other soft tissues of the thorax | T_MCOaaB/DGN_PAL |
| Soft tissue | CIM-10 (ICD10) | C4940 | Malignant tumor of abdominal vessels | T_MCOaaB/DGN_PAL |
| Soft tissue | CIM-10 (ICD10) | C4948 | Other malignant tumours of connective tissue and other soft tissues of the abdomen | T_MCOaaB/DGN_PAL |
| Soft tissue | CIM-10 (ICD10) | C4950 | Malignant tumor of the pelvic vessels | T_MCOaaB/DGN_PAL |
| Soft tissue | CIM-10 (ICD10) | C4958 | Other malignant tumors of connective tissue and other soft tissues of the pelvis | T_MCOaaB/DGN_PAL |
| Soft tissue | CIM-10 (ICD10) | C496 | Malignant neoplasm of conn and soft tissue of trunk, unsp | T_MCOaaB/DGN_PAL |
| Soft tissue | CIM-10 (ICD10) | C498 | Malignant neoplasm of ovrlp sites of conn and soft tissue | T_MCOaaB/DGN_PAL |
| Soft tissue | CIM-10 (ICD10) | C500 | Malignant tumor of the nipple and areola | T_MCOaaB/DGN_PAL |
| Soft tissue | CIM-10 (ICD10) | C501 | Malignant tumor of the central part of the breast | T_MCOaaB/DGN_PAL |
| Soft tissue | CIM-10 (ICD10) | C502 | Malignant tumor of the superior-internal quadrant of the breast | T_MCOaaB/DGN_PAL |
| Soft tissue | CIM-10 (ICD10) | C503 | Malignant tumor of the inferomedial quadrant of the breast | T_MCOaaB/DGN_PAL |
| Soft tissue | CIM-10 (ICD10) | C504 | Malignant tumor of the superolateral quadrant of the breast | T_MCOaaB/DGN_PAL |
| Soft tissue | CIM-10 (ICD10) | C505 | Malignant tumor of the inferolateral quadrant of the breast | T_MCOaaB/DGN_PAL |
| Soft tissue | CIM-10 (ICD10) | C508 | Tumeur maligne à localisations contiguës du sein | T_MCOaaB/DGN_PAL |
| Soft tissue | CIM-10 (ICD10) | C509 | Malignant breast tumor, unspecified | T_MCOaaB/DGN_PAL |
| Soft tissue | CIM-10 (ICD10) | C539 | Malignant neoplasm of cervix uteri, unspecified | T_MCOaaB/DGN_PAL |
| Soft tissue | CIM-10 (ICD10) | C541 | Malignant neoplasm of endometrium | T_MCOaaB/DGN_PAL |
| Soft tissue | CIM-10 (ICD10) | C542 | Malignant neoplasm of myometrium | T_MCOaaB/DGN_PAL |
| Soft tissue | CIM-10 (ICD10) | C548 | Malignant neoplasm of overlapping sites of corpus uteri | T_MCOaaB/DGN_PAL |
| Soft tissue | CIM-10 (ICD10) | C549 | Malignant neoplasm of corpus uteri, unspecified | T_MCOaaB/DGN_PAL |
| Soft tissue | CIM-10 (ICD10) | C55 | Malignant neoplasm of uterus, part unspecified | T_MCOaaB/DGN_PAL |
| Soft tissue | CIM-10 (ICD10) | C571 | Malignant tumor of a broad ligament | T_MCOaaB/DGN_PAL |
| Soft tissue | CIM-10 (ICD10) | C621 | Malignant tumor of the descending testicle | T_MCOaaB/DGN_PAL |
| Soft tissue | CIM-10 (ICD10) | C629 | Malignant testicular tumor, unspecified | T_MCOaaB/DGN_PAL |
| Soft tissue | CIM-10 (ICD10) | C630 | Malignant tumor of the epididymis | T_MCOaaB/DGN_PAL |
| Soft tissue | CIM-10 (ICD10) | C631 | Malignant tumor of the spermatic cord | T_MCOaaB/DGN_PAL |
| Soft tissue | CIM-10 (ICD10) | C632 | Malignant neoplasm of scrotum | T_MCOaaB/DGN_PAL |
| Soft tissue | CIM-10 (ICD10) | C637 | Malignant neoplasm of other specified male genital organs | T_MCOaaB/DGN_PAL |
| Soft tissue | CIM-10 (ICD10) | C64 | Malignant tumor of the kidney, excluding the renal pelvis | T_MCOaaB/DGN_PAL |
| Soft tissue | CIM-10 (ICD10) | C696 | Malignant tumor of the orbit | T_MCOaaB/DGN_PAL |
| Soft tissue | CIM-10 (ICD10) | C740 | Malignant tumor of the adrenal cortex | T_MCOaaB/DGN_PAL |
| Soft tissue | CIM-10 (ICD10) | C749 | Malignant adrenal tumor, unspecified | T_MCOaaB/DGN_PAL |
| Soft tissue | CIM-10 (ICD10) | C763 | Malignant neoplasm of pelvis | T_MCOaaB/DGN_PAL |
| Soft tissue | CIM-10 (ICD10) | C764 | Malignant tumor of ill-defined location in the upper limb | T_MCOaaB/DGN_PAL |
| Soft tissue | CIM-10 (ICD10) | C765 | Malignant tumor of poorly defined site in lower limb | T_MCOaaB/DGN_PAL |
| Soft tissue | CIM-10 (ICD10) | C770 | Sec and unsp malig neoplasm of nodes of head, face and neck | T_MCOaaB/DGN_PAL |
| Soft tissue | CIM-10 (ICD10) | C773 | Sec and unsp malig neoplasm of axilla and upper limb nodes | T_MCOaaB/DGN_PAL |
| Soft tissue | CIM-10 (ICD10) | C774 | Sec and unsp malig neoplasm of inguinal and lower limb nodes | T_MCOaaB/DGN_PAL |
| Soft tissue | CIM-10 (ICD10) | C795 | Secondary malignant tumor of bone and bone marrow | T_MCOaaB/DGN_PAL |
| Soft tissue | CIM-10 (ICD10) | C798 | Secondary malignancy in other specified sites | T_MCOaaB/DGN_PAL |
| Soft tissue | CIM-10 (ICD10) | D171 | Benign lipomatous neoplasm of skin, subcu of trunk | T_MCOaaB/DGN_PAL |
| Soft tissue | CIM-10 (ICD10) | D172 | Benign lipomatous tumor of the skin and subcutaneous tissue of the limbs | T_MCOaaB/DGN_PAL |
| Soft tissue | CIM-10 (ICD10) | D180 | Hemangioma, any site | T_MCOaaB/DGN_PAL |
| Soft tissue | CIM-10 (ICD10) | D211 | Benign tumor of the connective tissue and other soft tissues of the upper limb, including the shoulder | T_MCOaaB/DGN_PAL |
| Soft tissue | CIM-10 (ICD10) | D212 | Benign tumour of the connective tissue and other soft tissues of the lower limb, including the hip | T_MCOaaB/DGN_PAL |
| Soft tissue | CIM-10 (ICD10) | D236 | Benign skin tumor of the upper limb, including the shoulder | T_MCOaaB/DGN_PAL |
| Soft tissue | CIM-10 (ICD10) | D237 | Benign skin tumour of the lower limb, including the hip | T_MCOaaB/DGN_PAL |

| | | | | |
|---|---|---|---|---|
| Soft tissue | CIM-10 (ICD10) | D24 | Benign breast tumor | T_MCOaaB/DGN_PAL |
| Soft tissue | CIM-10 (ICD10) | D361 | Benign tumor of the peripheral nerves and autonomic nervous system | T_MCOaaB/DGN_PAL |
| Soft tissue | CIM-10 (ICD10) | D370 | Unpredictable or unknown tumor of the lip, oral cavity and pharynx | T_MCOaaB/DGN_PAL |
| Soft tissue | CIM-10 (ICD10) | D401 | Unpredictable or unknown testicular tumor | T_MCOaaB/DGN_PAL |
| Soft tissue | CIM-10 (ICD10) | D407 | Unpredictable or unknown tumor of other male genital organs | T_MCOaaB/DGN_PAL |
| Soft tissue | CIM-10 (ICD10) | D480 | Neoplasm of uncertain behavior of bone/artic cartl | T_MCOaaB/DGN_PAL |
| Soft tissue | CIM-10 (ICD10) | D481 | Neoplasm of uncertain behavior of connctv/soft tiss | T_MCOaaB/DGN_PAL |
| Soft tissue | CIM-10 (ICD10) | D482 | Neoplm of uncrt behav of prph nerves and autonm nervous sys | T_MCOaaB/DGN_PAL |
| Soft tissue | CIM-10 (ICD10) | D4838 | Other tumors of the retroperitoneum with unpredictable or unknown evolution | T_MCOaaB/DGN_PAL |
| Soft tissue | CIM-10 (ICD10) | D484 | Neoplasm of uncertain behavior of peritoneum | T_MCOaaB/DGN_PAL |
| Soft tissue | CIM-10 (ICD10) | D485 | Neoplasm of uncertain behavior of skin | T_MCOaaB/DGN_PAL |
| Soft tissue | CIM-10 (ICD10) | D486 | Unpredictable and unknown breast tumor | T_MCOaaB/DGN_PAL |
| Soft tissue | CIM-10 (ICD10) | D487 | Neoplasm of uncertain behavior of other specified sites | T_MCOaaB/DGN_PAL |
| Soft tissue | CIM-10 (ICD10) | D62 | Acute posthemorrhagic anemia | T_MCOaaB/DGN_PAL |
| Soft tissue | CCAM (procedures) | CAFA005 | Partial transfixing removal of the auricle | T_MCOaaA/CDC_ACT |
| Soft tissue | CCAM (procedures) | DAFA009 | Removal of a heart tumor by thoracotomy with bypass surgery | T_MCOaaA/CDC_ACT |
| Soft tissue | CCAM (procedures) | DGCA001 | Suture of a wound of the abdominal aorta, common iliac artery and/or external iliac artery, by laparotomy | T_MCOaaA/CDC_ACT |
| Soft tissue | CCAM (procedures) | DGFA015 | Laparotomy resection-anastomosis of abdominal aorta or common iliac artery | T_MCOaaA/CDC_ACT |
| Soft tissue | CCAM (procedures) | DGKA004 | Laparotomy replacement of abdominal aorta or common iliac artery | T_MCOaaA/CDC_ACT |
| Soft tissue | CCAM (procedures) | DHCA001 | Retrohepatic or suprahepatic inferior vena cava or hepatic vein wound suture, by laparotomy | T_MCOaaA/CDC_ACT |
| Soft tissue | CCAM (procedures) | DHCA004 | Ilioiliac or iliocaval venous bypass, by laparotomy | T_MCOaaA/CDC_ACT |
| Soft tissue | CCAM (procedures) | DHFA001 | Retrohepatic and/or suprahepatic suprarenal inferior vena cava resection with reconstruction, by laparotomy | T_MCOaaA/CDC_ACT |
| Soft tissue | CCAM (procedures) | DHFA002 | Infrarenal inferior vena cava resection without reconstruction, by laparotomy | T_MCOaaA/CDC_ACT |
| Soft tissue | CCAM (procedures) | DHFA006 | Resection of infrahepatic suprarenal inferior vena cava with reconstruction, by laparotomy | T_MCOaaA/CDC_ACT |
| Soft tissue | CCAM (procedures) | HFFA009 | Atypical partial resection of the stomach wall without interrupting continuity, by laparotomy | T_MCOaaA/CDC_ACT |
| Soft tissue | CCAM (procedures) | HGFA007 | Single segmental resection of the small intestine with restoration of continuity, outside the occlusion, by laparotomy | T_MCOaaA/CDC_ACT |
| Soft tissue | CCAM (procedures) | HHFA006 | Left colectomy with liberation of the left colonic angle, with restoration of continuity, by laparotomy | T_MCOaaA/CDC_ACT |
| Soft tissue | CCAM (procedures) | HHFA009 | Right colectomy with restoration of continuity, by laparotomy | T_MCOaaA/CDC_ACT |
| Soft tissue | CCAM (procedures) | HHFA011 | Appendectomy, by laparotomy | T_MCOaaA/CDC_ACT |
| Soft tissue | CCAM (procedures) | HHFA017 | Left colectomy without release of the left colonic angle, with restoration of continuity, by laparotomy | T_MCOaaA/CDC_ACT |
| Soft tissue | CCAM (procedures) | HHFA018 | Transverse colectomy, by laparotomy | T_MCOaaA/CDC_ACT |
| Soft tissue | CCAM (procedures) | HHQX006 | Pathological examination for carcinological purposes of partial colectomy or rectosigmoidectomy specimen without mesorectal resection | T_MCOaaA/CDC_ACT |
| Soft tissue | CCAM (procedures) | HJFA002 | Rectosigmoid resection with colorectal intraperitoneal anastomosis, by laparotomy | T_MCOaaA/CDC_ACT |
| Soft tissue | CCAM (procedures) | HMFA007 | Laparotomy cholecystectomy | T_MCOaaA/CDC_ACT |
| Soft tissue | CCAM (procedures) | HNFA007 | Cephalic duodenopancreatectomy, by laparotomy | T_MCOaaA/CDC_ACT |
| Soft tissue | CCAM (procedures) | HNFA013 | Left pancreatectomy with splenectomy [Left splenopancreatectomy], by laparotomy | T_MCOaaA/CDC_ACT |
| Soft tissue | CCAM (procedures) | HPFA003 | Removal of a lesion from a peritoneal fold [meso] without bowel resection, by laparotomy | T_MCOaaA/CDC_ACT |
| Soft tissue | CCAM (procedures) | HPFA004 | Resection of the omentum [omentum] [omentectomy], by laparotomy | T_MCOaaA/CDC_ACT |
| Soft tissue | CCAM (procedures) | HPMA001 | Intra-abdominal epiplasty by releasing the greater gastric curvature with pediculization on a gastroepiploic pedicle, during a laparotomy procedure | T_MCOaaA/CDC_ACT |
| Soft tissue | CCAM (procedures) | HPQX005 | Pathological examination for carcinological purposes of tumor removal specimens from the greater omentum, peritoneum and/or peritoneal fold [meso]. | T_MCOaaA/CDC_ACT |
| Soft tissue | CCAM (procedures) | HSLD001 | enteral tube feeding at 20 to 35 kilocalories per kilogram per day [kcal/kg/day], per 24 hours | T_MCOaaA/CDC_ACT |
| Soft tissue | CCAM (procedures) | HSLF002 | Parenteral nutrition with 20 to 35 kilocalories per kilogram per day [kcal/kg/day], per 24 hours | T_MCOaaA/CDC_ACT |
| Soft tissue | CCAM (procedures) | HSLF003 | Parenteral nutrition with more than 35 kilocalories per kilogram per day [kcal/kg/day], per 24 hours | T_MCOaaA/CDC_ACT |
| Soft tissue | CCAM (procedures) | JAFA005 | Total nephrectomy extended to the renal fossa with lateral resection of the inferior vena cava, by direct approach | T_MCOaaA/CDC_ACT |
| Soft tissue | CCAM (procedures) | JAFA009 | Total nephrectomy extended to the renal fossa, by laparotomy or lumbo-abdominal approach | T_MCOaaA/CDC_ACT |
| Soft tissue | CCAM (procedures) | JAFA023 | Unilateral total nephrectomy, by laparotomy | T_MCOaaA/CDC_ACT |
| Soft tissue | CCAM (procedures) | JAFA029 | Total nephrectomy extended to the renal fossa with adrenalectomy, by laparotomy or lumbo-abdominal approach | T_MCOaaA/CDC_ACT |
| Soft tissue | CCAM (procedures) | JAQX004 | Pathological examination for carcinological purposes of total nephrectomy or nephro-ureterectomy specimen | T_MCOaaA/CDC_ACT |
| Soft tissue | CCAM (procedures) | JAQX005 | Pathological examination of partial nephrectomy specimen for carcinological purposes | T_MCOaaA/CDC_ACT |
| Soft tissue | CCAM (procedures) | JCPA002 | Freeing the ureter without intraperitonization, using a direct approach | T_MCOaaA/CDC_ACT |
| Soft tissue | CCAM (procedures) | JDLD001 | Placement of a urethrovesical catheter [Indwelling bladder catheterization]. | T_MCOaaA/CDC_ACT |
| Soft tissue | CCAM (procedures) | JFFA010 | Removal of lesion from retroperitoneal space without dissection of large vessels, by laparotomy or lumbotomy | T_MCOaaA/CDC_ACT |
| Soft tissue | CCAM (procedures) | JFFA021 | Excision of retroperitoneal space lesion with dissection of large vessels, via direct approach | T_MCOaaA/CDC_ACT |
| Soft tissue | CCAM (procedures) | JHFA001 | removal of a spermatic cord cyst in adults, via inguinal approach | T_MCOaaA/CDC_ACT |
| Soft tissue | CCAM (procedures) | JHFA005 | Inguinal orchiectomy | T_MCOaaA/CDC_ACT |

| | | | | |
|---|---|---|---|---|
| Soft tissue | CCAM (procedures) | JHFA008 | Orchiectomy extended to the spermatic cord [Orchiepididymectomy], inguinal approach | T_MCOaaA/CDC_ACT |
| Soft tissue | CCAM (procedures) | JHQX005 | Pathological examination of total orchiectomy specimen for carcinological purposes | T_MCOaaA/CDC_ACT |
| Soft tissue | CCAM (procedures) | JJFA004 | Salpingoovariectomy [Annexectomy], by laparotomy | T_MCOaaA/CDC_ACT |
| Soft tissue | CCAM (procedures) | JKFA028 | Total hysterectomy with unilateral or bilateral adnexectomy, by laparotomy | T_MCOaaA/CDC_ACT |
| Soft tissue | CCAM (procedures) | PAQX004 | Pathological examination for carcinological purposes of bone and/or cartilage tumor excision specimens | T_MCOaaA/CDC_ACT |
| Soft tissue | CCAM (procedures) | PCPA003 | Musculotendinous desinsertion | T_MCOaaA/CDC_ACT |
| Soft tissue | CCAM (procedures) | PDFA001 | Removal of fascial and/or subfascial soft-tissue lesions, without dissection of a large vascular or nerve trunk | T_MCOaaA/CDC_ACT |
| Soft tissue | CCAM (procedures) | PDFA002 | Removal of fascial and/or subfascial soft-tissue lesions, with dissection of large vascular and/or nerve trunks | T_MCOaaA/CDC_ACT |
| Soft tissue | CCAM (procedures) | PDFA003 | Removal of fascial and/or subfascial soft-tissue lesions from the root of a limb, elbow crease or popliteal fossa | T_MCOaaA/CDC_ACT |
| Soft tissue | CCAM (procedures) | PDHA001 | Direct approach biopsy of subfascial soft tissue | T_MCOaaA/CDC_ACT |
| Soft tissue | CCAM (procedures) | PDQX005 | Anatomopathological examination for carcinological purposes of fascial and/or subfascial soft tissue tumor excision specimens [aponeurotic and/or subaponeurotic]. | T_MCOaaA/CDC_ACT |
| Soft tissue | CCAM (procedures) | PZMA004 | Repair with free cutaneous, fascial, fasciocutaneous or subcutaneous, muscular, musculocutaneous, musculotendinous or bony flap with vascular anastomoses | T_MCOaaA/CDC_ACT |
| Soft tissue | CCAM (procedures) | PZQX008 | Pathological examination for carcinological purposes of partial or total limb amputation specimens | T_MCOaaA/CDC_ACT |
| Viscera | GHM (DRG) | 01C041 | Non-traumatic craniotomies, age over 17, level 1 | T_MCOaaB/GRC_GHM |
| Viscera | GHM (DRG) | 01C042 | Non-traumatic craniotomies, age over 17, level 2 | T_MCOaaB/GRC_GHM |
| Viscera | GHM (DRG) | 01C113 | Craniotomies for tumors, age under 18, level 3 | T_MCOaaB/GRC_GHM |
| Viscera | GHM (DRG) | 01M27 | Other nervous system tumors | T_MCOaaB/GRC_GHM |
| Viscera | GHM (DRG) | 03C242 | Salivary gland procedures, level 2 | T_MCOaaB/GRC_GHM |
| Viscera | GHM (DRG) | 04C021 | Major thoracic procedures, level 1 | T_MCOaaB/GRC_GHM |
| Viscera | GHM (DRG) | 04C022 | Major thoracic procedures, level 2 | T_MCOaaB/GRC_GHM |
| Viscera | GHM (DRG) | 04C023 | Major thoracic procedures, level 3 | T_MCOaaB/GRC_GHM |
| Viscera | GHM (DRG) | 04C024 | Major thoracic procedures, level 4 | T_MCOaaB/GRC_GHM |
| Viscera | GHM (DRG) | 04C041 | Thoracoscopic procedures, level 1 | T_MCOaaB/GRC_GHM |
| Viscera | GHM (DRG) | 05C062 | Other cardiothoracic procedures, age over 1 year, or vascular procedures regardless of age, with extracorporeal circulation, level 2 | T_MCOaaB/GRC_GHM |
| Viscera | GHM (DRG) | 05C063 | Other cardiothoracic procedures, age over 1 year, or vascular procedures of any age, with extracorporeal circulation, level 3 | T_MCOaaB/GRC_GHM |
| Viscera | GHM (DRG) | 05C172 | Vein ligation and stripping, level 2 | T_MCOaaB/GRC_GHM |
| Viscera | GHM (DRG) | 05C182 | Other procedures on the circulatory system, level 2 | T_MCOaaB/GRC_GHM |
| Viscera | GHM (DRG) | 06C031 | Rectal resection, level 1 | T_MCOaaB/GRC_GHM |
| Viscera | GHM (DRG) | 06C032 | Rectal resection, level 2 | T_MCOaaB/GRC_GHM |
| Viscera | GHM (DRG) | 06C033 | Rectal resection, level 3 | T_MCOaaB/GRC_GHM |
| Viscera | GHM (DRG) | 06C034 | Rectal resections, level 4 | T_MCOaaB/GRC_GHM |
| Viscera | GHM (DRG) | 06C041 | Major small bowel and colon procedures, level 1 | T_MCOaaB/GRC_GHM |
| Viscera | GHM (DRG) | 06C042 | Major small bowel and colon procedures, level 2 | T_MCOaaB/GRC_GHM |
| Viscera | GHM (DRG) | 06C043 | Major small bowel and colon procedures, level 3 | T_MCOaaB/GRC_GHM |
| Viscera | GHM (DRG) | 06C044 | Major small bowel and colon procedures, level 4 | T_MCOaaB/GRC_GHM |
| Viscera | GHM (DRG) | 06C072 | Minor small bowel and colon procedures, level 2 | T_MCOaaB/GRC_GHM |
| Viscera | GHM (DRG) | 06C10J | Reconstructive hernia and eventration procedures, under 18 years of age, outpatient | T_MCOaaB/GRC_GHM |
| Viscera | GHM (DRG) | 06C141 | Rectal and anal procedures other than rectal resections, level 1 | T_MCOaaB/GRC_GHM |
| Viscera | GHM (DRG) | 06C142 | Rectum and anus procedures other than rectal resections, level 2 | T_MCOaaB/GRC_GHM |
| Viscera | GHM (DRG) | 06C143 | Rectum and anus procedures other than rectal resections, level 3 | T_MCOaaB/GRC_GHM |
| Viscera | GHM (DRG) | 06C14J | Outpatient rectal and anal procedures other than rectal resections | T_MCOaaB/GRC_GHM |
| Viscera | GHM (DRG) | 06C151 | Other digestive tract procedures other than laparotomy, level 1 | T_MCOaaB/GRC_GHM |
| Viscera | GHM (DRG) | 06C161 | Procedures on the esophagus, stomach and duodenum for malignant tumors, age over 17, level 1 | T_MCOaaB/GRC_GHM |
| Viscera | GHM (DRG) | 06C162 | Procedures on the esophagus, stomach and duodenum for malignant tumors, age over 17, level 2 | T_MCOaaB/GRC_GHM |
| Viscera | GHM (DRG) | 06C163 | Procedures on the esophagus, stomach and duodenum for malignant tumors, age over 17, level 3 | T_MCOaaB/GRC_GHM |
| Viscera | GHM (DRG) | 06C164 | Operations on the esophagus, stomach and duodenum for malignant tumors, age over 17, level 4 | T_MCOaaB/GRC_GHM |
| Viscera | GHM (DRG) | 06C211 | Other operations on the digestive tract by laparotomy, level 1 | T_MCOaaB/GRC_GHM |
| Viscera | GHM (DRG) | 06C212 | Other laparotomy operations on the digestive tract, level 2 | T_MCOaaB/GRC_GHM |
| Viscera | GHM (DRG) | 06C221 | Procedures on the esophagus, stomach and duodenum for non-malignant conditions or ulcers, age over 17, level 1 | T_MCOaaB/GRC_GHM |
| Viscera | GHM (DRG) | 06C222 | Procedures on the esophagus, stomach and duodenum for non-malignant conditions or ulcers, age over 17, level 2 | T_MCOaaB/GRC_GHM |
| Viscera | GHM (DRG) | 06C223 | Procedures on the esophagus, stomach and duodenum for non-malignant conditions or ulcers, age over 17, level 3 | T_MCOaaB/GRC_GHM |
| Viscera | GHM (DRG) | 06C224 | Operations on oesophagus, stomach and duodenum for non-malignant conditions or ulcers, age over 17, level 4 | T_MCOaaB/GRC_GHM |
| Viscera | GHM (DRG) | 06M05 | Other malignant tumors of the digestive tract | T_MCOaaB/GRC_GHM |
| Viscera | GHM (DRG) | 07C091 | Procedures on the liver, pancreas, portal or vena cava for malignant tumors, level 1 | T_MCOaaB/GRC_GHM |

| | | | | |
|---|---|---|---|---|
| Viscera | GHM (DRG) | 07C092 | Procedures on the liver, pancreas and portal or vena cava for malignant tumors, level 2 | T_MCOaaB/GRC_GHM |
| Viscera | GHM (DRG) | 07C093 | Procedures on the liver, pancreas and portal or vena cava for malignant tumors, level 3 | T_MCOaaB/GRC_GHM |
| Viscera | GHM (DRG) | 07C094 | Procedures on the liver, pancreas and portal or vena cava for malignant tumors, level 4 | T_MCOaaB/GRC_GHM |
| Viscera | GHM (DRG) | 07C101 | Procedures on the liver, pancreas and portal or vena cava for non-malignant conditions, level 1 | T_MCOaaB/GRC_GHM |
| Viscera | GHM (DRG) | 07C103 | Operations on the liver, pancreas and portal or vena cava for non-malignant conditions, level 3 | T_MCOaaB/GRC_GHM |
| Viscera | GHM (DRG) | 08C201 | Skin grafts for musculoskeletal or connective tissue disease, level 1 | T_MCOaaB/GRC_GHM |
| Viscera | GHM (DRG) | 08C202 | Skin grafts for musculoskeletal or connective tissue diseases, level 2 | T_MCOaaB/GRC_GHM |
| Viscera | GHM (DRG) | 08C211 | Other musculoskeletal and connective tissue procedures, level 1 | T_MCOaaB/GRC_GHM |
| Viscera | GHM (DRG) | 08C212 | Other musculoskeletal and connective tissue procedures, level 2 | T_MCOaaB/GRC_GHM |
| Viscera | GHM (DRG) | 09C031 | Skin grafts and/or wound trimming, excluding skin ulcers and cellulitis, level 1 | T_MCOaaB/GRC_GHM |
| Viscera | GHM (DRG) | 09C041 | Total mastectomy for malignancy, level 1 | T_MCOaaB/GRC_GHM |
| Viscera | GHM (DRG) | 09C042 | Total mastectomy for malignancy, level 2 | T_MCOaaB/GRC_GHM |
| Viscera | GHM (DRG) | 09C043 | Total mastectomy for malignancy, level 3 | T_MCOaaB/GRC_GHM |
| Viscera | GHM (DRG) | 09C044 | Total mastectomy for malignancy, level 4 | T_MCOaaB/GRC_GHM |
| Viscera | GHM (DRG) | 09C051 | Subtotal mastectomies for malignancy, level 1 | T_MCOaaB/GRC_GHM |
| Viscera | GHM (DRG) | 09C052 | Subtotal mastectomies for malignancy, level 2 | T_MCOaaB/GRC_GHM |
| Viscera | GHM (DRG) | 09C05J | Subtotal mastectomies for malignancy, outpatient | T_MCOaaB/GRC_GHM |
| Viscera | GHM (DRG) | 09C061 | Breast procedures for non-malignant conditions other than biopsy and local excision procedures, level 1 | T_MCOaaB/GRC_GHM |
| Viscera | GHM (DRG) | 09C062 | Breast procedures for non-malignant conditions other than biopsy and local excision procedures, level 2 | T_MCOaaB/GRC_GHM |
| Viscera | GHM (DRG) | 09C06T | Breast procedures for non-malignant conditions other than biopsies and local excisions, very short duration | T_MCOaaB/GRC_GHM |
| Viscera | GHM (DRG) | 09C071 | Biopsies and local excisions for non-malignant breast conditions, level 1 | T_MCOaaB/GRC_GHM |
| Viscera | GHM (DRG) | 09C101 | Other skin, subcutaneous tissue or breast procedures, level 1 | T_MCOaaB/GRC_GHM |
| Viscera | GHM (DRG) | 09C103 | Other skin, subcutaneous tissue or breast procedures, level 3 | T_MCOaaB/GRC_GHM |
| Viscera | GHM (DRG) | 09C10J | Other skin, subcutaneous tissue or breast procedures, outpatient | T_MCOaaB/GRC_GHM |
| Viscera | GHM (DRG) | 09C111 | Breast reconstruction, level 1 | T_MCOaaB/GRC_GHM |
| Viscera | GHM (DRG) | 09C112 | Breast reconstruction, level 2 | T_MCOaaB/GRC_GHM |
| Viscera | GHM (DRG) | 09C114 | Breast reconstruction, level 4 | T_MCOaaB/GRC_GHM |
| Viscera | GHM (DRG) | 10C032 | Adrenal gland procedures, level 2 | T_MCOaaB/GRC_GHM |
| Viscera | GHM (DRG) | 10C034 | Adrenal gland procedures, level 4 | T_MCOaaB/GRC_GHM |
| Viscera | GHM (DRG) | 10C111 | Thyroid procedures for malignant tumors, level 1 | T_MCOaaB/GRC_GHM |
| Viscera | GHM (DRG) | 10C112 | Thyroid procedures for malignancy, level 2 | T_MCOaaB/GRC_GHM |
| Viscera | GHM (DRG) | 11C021 | Kidney and ureter procedures and major bladder surgery for tumor disease, level 1 | T_MCOaaB/GRC_GHM |
| Viscera | GHM (DRG) | 11C022 | Kidney and ureter procedures and major bladder surgery for tumour disease, level 2 | T_MCOaaB/GRC_GHM |
| Viscera | GHM (DRG) | 11C023 | Kidney and ureter procedures and major bladder surgery for tumor disease, level 3 | T_MCOaaB/GRC_GHM |
| Viscera | GHM (DRG) | 11C024 | Kidney and ureter procedures and major bladder surgery for tumoral disease, level 4 | T_MCOaaB/GRC_GHM |
| Viscera | GHM (DRG) | 11C042 | Other bladder procedures excluding transurethral procedures, level 2 | T_MCOaaB/GRC_GHM |
| Viscera | GHM (DRG) | 11C051 | Transurethral or transcutaneous procedures, level 1 | T_MCOaaB/GRC_GHM |
| Viscera | GHM (DRG) | 11C052 | Transurethral or transcutaneous procedures, level 2 | T_MCOaaB/GRC_GHM |
| Viscera | GHM (DRG) | 11C083 | Other kidney and urinary tract procedures, level 3 | T_MCOaaB/GRC_GHM |
| Viscera | GHM (DRG) | 11C131 | Transurethral or transcutaneous procedures for non-lithiasis, level 1 | T_MCOaaB/GRC_GHM |
| Viscera | GHM (DRG) | 11C13J | Transurethral or transcutaneous procedures for non-lithiasis, on an outpatient basis | T_MCOaaB/GRC_GHM |
| Viscera | GHM (DRG) | 12C042 | Transurethral prostatectomies, level 2 | T_MCOaaB/GRC_GHM |
| Viscera | GHM (DRG) | 12C051 | Testicular procedures for malignant tumors, level 1 | T_MCOaaB/GRC_GHM |
| Viscera | GHM (DRG) | 12C091 | Other procedures for malignant tumors of the male genital tract, level 1 | T_MCOaaB/GRC_GHM |
| Viscera | GHM (DRG) | 12C111 | Major male pelvic procedures for malignancies, level 1 | T_MCOaaB/GRC_GHM |
| Viscera | GHM (DRG) | 12C112 | Major male pelvic procedures for malignancies, level 2 | T_MCOaaB/GRC_GHM |
| Viscera | GHM (DRG) | 12C114 | Major male pelvic surgery for malignancy, level 4 | T_MCOaaB/GRC_GHM |
| Viscera | GHM (DRG) | 13C031 | Hysterectomies, level 1 | T_MCOaaB/GRC_GHM |
| Viscera | GHM (DRG) | 13C032 | Hysterectomies, level 2 | T_MCOaaB/GRC_GHM |
| Viscera | GHM (DRG) | 13C033 | Hysterectomies, level 3 | T_MCOaaB/GRC_GHM |
| Viscera | GHM (DRG) | 13C034 | Hysterectomies, level 4 | T_MCOaaB/GRC_GHM |
| Viscera | GHM (DRG) | 13C051 | Uteroannexal procedures for malignant tumors, level 1 | T_MCOaaB/GRC_GHM |
| Viscera | GHM (DRG) | 13C052 | Procedures on the uteroannexal system for malignant tumors, level 2 | T_MCOaaB/GRC_GHM |
| Viscera | GHM (DRG) | 13C071 | Uteroannexal procedures for non-malignant conditions, other than tubal interruptions, level 1 | T_MCOaaB/GRC_GHM |
| Viscera | GHM (DRG) | 13C08] | Utero-annexal procedures for non-malignant conditions, other than tubal interruptions, on an outpatient basis | T_MCOaaB/GRC_GHM |
| Viscera | GHM (DRG) | 13C081 | Procedures on the vulva, vagina or cervix, level 1 | T_MCOaaB/GRC_GHM |

| | | | | |
|---|---|---|---|---|
| Viscera | GHM (DRG) | 13C09J | Laparoscopic or laparoscopic diagnostic procedures, on an outpatient basis | T_MCOaaB/GRC_GHM |
| Viscera | GHM (DRG) | 13C113 | Dilatations and curettages, conizations for malignant tumors, level 3 | T_MCOaaB/GRC_GHM |
| Viscera | GHM (DRG) | 13C132 | Other female genital tract procedures, level 2 | T_MCOaaB/GRC_GHM |
| Viscera | GHM (DRG) | 13C141 | Pelvic exenteration, extended hysterectomy or vulvectomy for malignancy, level 1 | T_MCOaaB/GRC_GHM |
| Viscera | GHM (DRG) | 13C142 | Pelvic exenteration, enlarged hysterectomy or vulvectomy for malignancy, level 2 | T_MCOaaB/GRC_GHM |
| Viscera | GHM (DRG) | 13C143 | Pelvic exenteration, enlarged hysterectomy or vulvectomy for malignancy, level 3 | T_MCOaaB/GRC_GHM |
| Viscera | GHM (DRG) | 13C144 | Pelvic exenteration, enlarged hysterectomy or vulvectomy for malignancy, level 4 | T_MCOaaB/GRC_GHM |
| Viscera | GHM (DRG) | 13C151 | Pelvic exenteration, enlarged hysterectomy or vulvectomy for non-malignant conditions, level 1 | T_MCOaaB/GRC_GHM |
| Viscera | GHM (DRG) | 13C152 | Pelvic exenteration, enlarged hysterectomy or vulvectomy for non-malignant conditions, level 2 | T_MCOaaB/GRC_GHM |
| Viscera | GHM (DRG) | 13C181 | Myomectomies of the uterus, level 1 | T_MCOaaB/GRC_GHM |
| Viscera | GHM (DRG) | 13C201 | Exeresis or destruction of cervical lesions except conization, level 1 | T_MCOaaB/GRC_GHM |
| Viscera | GHM (DRG) | 13M07 | Other tumors of the female genital tract | T_MCOaaB/GRC_GHM |
| Viscera | GHM (DRG) | 16C023 | Spleen surgery, level 3 | T_MCOaaB/GRC_GHM |
| Viscera | GHM (DRG) | 17C021 | Major surgery for lymphoma or leukemia, level 1 | T_MCOaaB/GRC_GHM |
| Viscera | GHM (DRG) | 17C022 | Major surgery for lymphoma or leukemia, level 2 | T_MCOaaB/GRC_GHM |
| Viscera | GHM (DRG) | 17C041 | Major procedures for myeloproliferative disorders or tumors of unclear or diffuse location, level 1 | T_MCOaaB/GRC_GHM |
| Viscera | GHM (DRG) | 17C042 | Major procedures for myeloproliferative disorders or tumors of imprecise or diffuse location, level 2 | T_MCOaaB/GRC_GHM |
| Viscera | GHM (DRG) | 17C043 | Major procedures for myeloproliferative disorders or tumors of imprecise or diffuse site, level 3 | T_MCOaaB/GRC_GHM |
| Viscera | GHM (DRG) | 17C044 | Major procedures for myeloproliferative disorders or tumors of unclear or diffuse location, level 4 | T_MCOaaB/GRC_GHM |
| Viscera | GHM (DRG) | 17C051 | Other procedures for myeloproliferative disorders or tumors of unclear or diffuse location, level 1 | T_MCOaaB/GRC_GHM |
| Viscera | GHM (DRG) | 17C052 | Other procedures for myeloproliferative disorders or tumors of unclear or diffuse site, level 2 | T_MCOaaB/GRC_GHM |
| Viscera | GHM (DRG) | 17C053 | Other procedures for myeloproliferative disorders or tumors of unclear or diffuse site, level 3 | T_MCOaaB/GRC_GHM |
| Viscera | GHM (DRG) | 17C05J | Other outpatient procedures for myeloproliferative disorders or tumors of unclear or diffuse location | T_MCOaaB/GRC_GHM |
| Viscera | GHM (DRG) | 17C061 | Major procedures in CMD17 (Myeloproliferative disorders and tumors of unclear or diffuse location.), level 1 | T_MCOaaB/GRC_GHM |
| Viscera | GHM (DRG) | 17C062 | Major procedures in CMD17 (Myeloproliferative disorders and tumors of unclear or diffuse location.), level 2 | T_MCOaaB/GRC_GHM |
| Viscera | GHM (DRG) | 17C063 | Major procedures in CMD17 (Myeloproliferative disorders and tumors of unclear or diffuse location.), level 3 | T_MCOaaB/GRC_GHM |
| Viscera | GHM (DRG) | 17C064 | Major procedures in CMD17 (Myeloproliferative disorders and tumors of unclear or diffuse location.), level 4 | T_MCOaaB/GRC_GHM |
| Viscera | GHM (DRG) | 17C071 | Intermediate procedures of CMD17 (Myeloproliferative disorders and tumors of unclear or diffuse location.), level 1 | T_MCOaaB/GRC_GHM |
| Viscera | GHM (DRG) | 17C081 | Minor interventions for CMD17 (Myeloproliferative disorders and tumors of unclear or diffuse location.), level 1 | T_MCOaaB/GRC_GHM |
| Viscera | GHM (DRG) | 17M17 | Other conditions and tumors of imprecise or diffuse site | T_MCOaaB/GRC_GHM |
| Viscera | CIM-10 (ICD10) | C160 | Malignant neoplasm of cardia | T_MCOaaB/DGN_PAL |
| Viscera | CIM-10 (ICD10) | C161 | Malignant neoplasm of fundus of stomach | T_MCOaaB/DGN_PAL |
| Viscera | CIM-10 (ICD10) | C162 | Malignant neoplasm of body of stomach | T_MCOaaB/DGN_PAL |
| Viscera | CIM-10 (ICD10) | C163 | Malignant neoplasm of pyloric antrum | T_MCOaaB/DGN_PAL |
| Viscera | CIM-10 (ICD10) | C165 | Malignant neoplasm of lesser curvature of stomach, unsp | T_MCOaaB/DGN_PAL |
| Viscera | CIM-10 (ICD10) | C166 | Malignant neoplasm of greater curvature of stomach, unsp | T_MCOaaB/DGN_PAL |
| Viscera | CIM-10 (ICD10) | C168 | Malignant neoplasm of overlapping sites of stomach | T_MCOaaB/DGN_PAL |
| Viscera | CIM-10 (ICD10) | C169 | Malignant neoplasm of stomach, unspecified | T_MCOaaB/DGN_PAL |
| Viscera | CIM-10 (ICD10) | C170 | Malignant neoplasm of duodenum | T_MCOaaB/DGN_PAL |
| Viscera | CIM-10 (ICD10) | C171 | Malignant neoplasm of jejunum | T_MCOaaB/DGN_PAL |
| Viscera | CIM-10 (ICD10) | C172 | Malignant neoplasm of ileum | T_MCOaaB/DGN_PAL |
| Viscera | CIM-10 (ICD10) | C178 | Malignant neoplasm of overlapping sites of small intestine | T_MCOaaB/DGN_PAL |
| Viscera | CIM-10 (ICD10) | C179 | Malignant neoplasm of small intestine, unspecified | T_MCOaaB/DGN_PAL |
| Viscera | CIM-10 (ICD10) | C182 | Malignant neoplasm of ascending colon | T_MCOaaB/DGN_PAL |
| Viscera | CIM-10 (ICD10) | C186 | Malignant neoplasm of descending colon | T_MCOaaB/DGN_PAL |
| Viscera | CIM-10 (ICD10) | C187 | Malignant neoplasm of sigmoid colon | T_MCOaaB/DGN_PAL |
| Viscera | CIM-10 (ICD10) | C19 | Malignant neoplasm of rectosigmoid junction | T_MCOaaB/DGN_PAL |
| Viscera | CIM-10 (ICD10) | C20 | Malignant neoplasm of rectum | T_MCOaaB/DGN_PAL |
| Viscera | CIM-10 (ICD10) | C220 | Liver cell carcinoma | T_MCOaaB/DGN_PAL |
| Viscera | CIM-10 (ICD10) | C229 | Malig neoplasm of liver, not specified as primary or sec | T_MCOaaB/DGN_PAL |
| Viscera | CIM-10 (ICD10) | C250 | Malignant neoplasm of head of pancreas | T_MCOaaB/DGN_PAL |
| Viscera | CIM-10 (ICD10) | C261 | Malignant neoplasm of spleen | T_MCOaaB/DGN_PAL |
| Viscera | CIM-10 (ICD10) | C268 | Malignant tumor with contiguous digestive tract sites | T_MCOaaB/DGN_PAL |
| Viscera | CIM-10 (ICD10) | C341 | Malignant tumor of the upper lobe, bronchi or lung | T_MCOaaB/DGN_PAL |
| Viscera | CIM-10 (ICD10) | C343 | Malignant tumor of the lower lobe, bronchi or lung | T_MCOaaB/DGN_PAL |
| Viscera | CIM-10 (ICD10) | C349 | Tumeur maligne de bronche ou du poumon, sans précision | T_MCOaaB/DGN_PAL |

| | | | | |
|---|---|---|---|---|
| Viscera | CIM-10 (ICD10) | C380 | Malignant neoplasm of heart | T_MCOaaB/DGN_PAL |
| Viscera | CIM-10 (ICD10) | C383 | Malignant neoplasm of mediastinum, part unspecified | T_MCOaaB/DGN_PAL |
| Viscera | CIM-10 (ICD10) | C384 | Malignant neoplasm of pleura | T_MCOaaB/DGN_PAL |
| Viscera | CIM-10 (ICD10) | C480 | Malignant neoplasm of retroperitoneum | T_MCOaaB/DGN_PAL |
| Viscera | CIM-10 (ICD10) | C481 | Malignant neoplasm of specified parts of peritoneum | T_MCOaaB/DGN_PAL |
| Viscera | CIM-10 (ICD10) | C4938 | Other malignant tumors of connective tissue and other soft tissues of the thorax | T_MCOaaB/DGN_PAL |
| Viscera | CIM-10 (ICD10) | C4948 | Other malignant tumours of connective tissue and other soft tissues of the abdomen | T_MCOaaB/DGN_PAL |
| Viscera | CIM-10 (ICD10) | C4958 | Other malignant tumors of connective tissue and other soft tissues of the pelvis | T_MCOaaB/DGN_PAL |
| Viscera | CIM-10 (ICD10) | C500 | Malignant tumor of the nipple and areola | T_MCOaaB/DGN_PAL |
| Viscera | CIM-10 (ICD10) | C501 | Malignant tumor of the central part of the breast | T_MCOaaB/DGN_PAL |
| Viscera | CIM-10 (ICD10) | C502 | Malignant tumor of the superior-internal quadrant of the breast | T_MCOaaB/DGN_PAL |
| Viscera | CIM-10 (ICD10) | C503 | Malignant tumor of the inferomedial quadrant of the breast | T_MCOaaB/DGN_PAL |
| Viscera | CIM-10 (ICD10) | C504 | Malignant tumor of the superolateral quadrant of the breast | T_MCOaaB/DGN_PAL |
| Viscera | CIM-10 (ICD10) | C505 | Malignant tumor of the inferolateral quadrant of the breast | T_MCOaaB/DGN_PAL |
| Viscera | CIM-10 (ICD10) | C508 | Tumeur maligne à localisations contiguës du sein | T_MCOaaB/DGN_PAL |
| Viscera | CIM-10 (ICD10) | C509 | Malignant breast tumor, unspecified | T_MCOaaB/DGN_PAL |
| Viscera | CIM-10 (ICD10) | C510 | Malignant neoplasm of labium majus | T_MCOaaB/DGN_PAL |
| Viscera | CIM-10 (ICD10) | C530 | Malignant neoplasm of endocervix | T_MCOaaB/DGN_PAL |
| Viscera | CIM-10 (ICD10) | C538 | Malignant neoplasm of overlapping sites of cervix uteri | T_MCOaaB/DGN_PAL |
| Viscera | CIM-10 (ICD10) | C539 | Malignant neoplasm of cervix uteri, unspecified | T_MCOaaB/DGN_PAL |
| Viscera | CIM-10 (ICD10) | C540 | Malignant neoplasm of isthmus uteri | T_MCOaaB/DGN_PAL |
| Viscera | CIM-10 (ICD10) | C541 | Malignant neoplasm of endometrium | T_MCOaaB/DGN_PAL |
| Viscera | CIM-10 (ICD10) | C542 | Malignant neoplasm of myometrium | T_MCOaaB/DGN_PAL |
| Viscera | CIM-10 (ICD10) | C543 | Malignant neoplasm of fundus uteri | T_MCOaaB/DGN_PAL |
| Viscera | CIM-10 (ICD10) | C548 | Malignant neoplasm of overlapping sites of corpus uteri | T_MCOaaB/DGN_PAL |
| Viscera | CIM-10 (ICD10) | C549 | Malignant neoplasm of corpus uteri, unspecified | T_MCOaaB/DGN_PAL |
| Viscera | CIM-10 (ICD10) | C55 | Malignant neoplasm of uterus, part unspecified | T_MCOaaB/DGN_PAL |
| Viscera | CIM-10 (ICD10) | C56 | Malignant tumor of the ovary | T_MCOaaB/DGN_PAL |
| Viscera | CIM-10 (ICD10) | C578 | Malignant neoplasm of ovrlp sites of female genital organs | T_MCOaaB/DGN_PAL |
| Viscera | CIM-10 (ICD10) | C61 | Malignant neoplasm of prostate | T_MCOaaB/DGN_PAL |
| Viscera | CIM-10 (ICD10) | C621 | Malignant tumor of the descending testicle | T_MCOaaB/DGN_PAL |
| Viscera | CIM-10 (ICD10) | C629 | Malignant testicular tumor, unspecified | T_MCOaaB/DGN_PAL |
| Viscera | CIM-10 (ICD10) | C631 | Malignant tumor of the spermatic cord | T_MCOaaB/DGN_PAL |
| Viscera | CIM-10 (ICD10) | C64 | Malignant tumor of the kidney, excluding the renal pelvis | T_MCOaaB/DGN_PAL |
| Viscera | CIM-10 (ICD10) | C679 | Malignant neoplasm of bladder, unspecified | T_MCOaaB/DGN_PAL |
| Viscera | CIM-10 (ICD10) | C73 | Malignant neoplasm of thyroid gland | T_MCOaaB/DGN_PAL |
| Viscera | CIM-10 (ICD10) | C786 | Secondary malignant neoplasm of retroperiton and peritoneum | T_MCOaaB/DGN_PAL |
| Viscera | CIM-10 (ICD10) | D131 | Benign neoplasm of stomach | T_MCOaaB/DGN_PAL |
| Viscera | CIM-10 (ICD10) | D250 | Submucous leiomyoma of uterus | T_MCOaaB/DGN_PAL |
| Viscera | CIM-10 (ICD10) | D251 | Intramural leiomyoma of uterus | T_MCOaaB/DGN_PAL |
| Viscera | CIM-10 (ICD10) | D252 | Subserosal leiomyoma of uterus | T_MCOaaB/DGN_PAL |
| Viscera | CIM-10 (ICD10) | D259 | Leiomyoma of uterus, unspecified | T_MCOaaB/DGN_PAL |
| Viscera | CIM-10 (ICD10) | D371 | Neoplasm of uncertain behavior of stomach | T_MCOaaB/DGN_PAL |
| Viscera | CIM-10 (ICD10) | D372 | Neoplasm of uncertain behavior of small intestine | T_MCOaaB/DGN_PAL |
| Viscera | CIM-10 (ICD10) | D377 | Unpredictable or unknown tumors of other digestive organs | T_MCOaaB/DGN_PAL |
| Viscera | CIM-10 (ICD10) | D381 | Neoplasm of uncertain behavior of trachea, bronchus and lung | T_MCOaaB/DGN_PAL |
| Viscera | CIM-10 (ICD10) | D390 | Neoplasm of uncertain behavior of uterus | T_MCOaaB/DGN_PAL |
| Viscera | CIM-10 (ICD10) | D391 | Unpredictable or unknown evolution of ovarian tumor | T_MCOaaB/DGN_PAL |
| Viscera | CIM-10 (ICD10) | D486 | Unpredictable and unknown breast tumor | T_MCOaaB/DGN_PAL |
| Viscera | CIM-10 (ICD10) | D62 | Acute posthemorrhagic anemia | T_MCOaaB/DGN_PAL |
| Viscera | CCAM (procedures) | DZQJ001 | Esophageal Doppler Ultrasound of the Heart and Intrathoracic Vessels [Transesophageal Doppler Echocardiography] - Transthoracic Doppler Ultrasound of the Heart and Intrathoracic Vessels | T_MCOaaA/CDC_ACT |
| Viscera | CCAM (procedures) | DZQJ006 | Transthoracic doppler ultrasound of the heart and intrathoracic vessels | T_MCOaaA/CDC_ACT |
| Viscera | CCAM (procedures) | DZQM005 | Bedside transthoracic Doppler ultrasound of the heart and intrathoracic vessels | T_MCOaaA/CDC_ACT |
| Viscera | CCAM (procedures) | DZQX005 | Anatomopathological examination of heart tumor excision specimens for carcinological purposes | T_MCOaaA/CDC_ACT |
| Viscera | CCAM (procedures) | HEQE002 | Oeso-gastro-duodenal endoscopy | T_MCOaaA/CDC_ACT |
| Viscera | CCAM (procedures) | HFFA005 | Total gastrectomy with restoration of continuity, by laparotomy | T_MCOaaA/CDC_ACT |

| | | | | |
|---|---|---|---|---|
| Viscera | CCAM (procedures) | HFFA006 | Lower partial gastrectomy with gastrojejunal anastomosis, by laparotomy | T_MCOaaA/CDC_ACT |
| Viscera | CCAM (procedures) | HFFA009 | Atypical partial resection of the stomach wall without interruption of continuity, by laparotomy | T_MCOaaA/CDC_ACT |
| Viscera | CCAM (procedures) | HFFC001 | Atypical partial resection of the stomach wall not interrupting continuity, by laparoscopy | T_MCOaaA/CDC_ACT |
| Viscera | CCAM (procedures) | HFQX004 | Pathological examination of partial gastrectomy specimen for carcinological purposes | T_MCOaaA/CDC_ACT |
| Viscera | CCAM (procedures) | HGFA001 | Laparotomy resection of duodenojejunal angle with restoration of continuity | T_MCOaaA/CDC_ACT |
| Viscera | CCAM (procedures) | HGFA004 | Multiple segmental resection of the small intestine, by laparotomy | T_MCOaaA/CDC_ACT |
| Viscera | CCAM (procedures) | HGFA005 | Single-segment resection of the small intestine for occlusion, by laparotomy | T_MCOaaA/CDC_ACT |
| Viscera | CCAM (procedures) | HGFA007 | Single segmental resection of the small intestine with restoration of continuity, outside occlusion, by laparotomy | T_MCOaaA/CDC_ACT |
| Viscera | CCAM (procedures) | HGFC021 | Single-segment resection of the small intestine with restoration of continuity, outside the occlusion, by laparoscopy | T_MCOaaA/CDC_ACT |
| Viscera | CCAM (procedures) | HGQX008 | Pathological examination for carcinological purposes of a small bowel segmental excision specimen | T_MCOaaA/CDC_ACT |
| Viscera | CCAM (procedures) | HHFA006 | Left colectomy with liberation of the left colonic angle, with restoration of continuity, by laparotomy | T_MCOaaA/CDC_ACT |
| Viscera | CCAM (procedures) | HHFA009 | Right colectomy with restoration of continuity, by laparotomy | T_MCOaaA/CDC_ACT |
| Viscera | CCAM (procedures) | HHFA011 | Appendectomy, by laparotomy | T_MCOaaA/CDC_ACT |
| Viscera | CCAM (procedures) | HHQX006 | Pathological examination for carcinological purposes of partial colectomy or rectosigmoidectomy specimen without mesorectum resection | T_MCOaaA/CDC_ACT |
| Viscera | CCAM (procedures) | HMFA007 | Laparotomy cholecystectomy | T_MCOaaA/CDC_ACT |
| Viscera | CCAM (procedures) | HNFA007 | Cephalic duodenopancreatectomy, by laparotomy | T_MCOaaA/CDC_ACT |
| Viscera | CCAM (procedures) | HNFA013 | Left pancreatectomy with splenectomy [Left splenopancreatectomy], by laparotomy | T_MCOaaA/CDC_ACT |
| Viscera | CCAM (procedures) | HPFA004 | Omentum resection [Omentectomy], by laparotomy | T_MCOaaA/CDC_ACT |
| Viscera | CCAM (procedures) | HSLF002 | Parenteral nutrition with intake of 20 to 35 kilocalories per kilogram per day [kcal/kg/day], per 24 hours | T_MCOaaA/CDC_ACT |
| Viscera | CCAM (procedures) | JAFA029 | Total nephrectomy extended to the renal pelvis with adrenalectomy, by laparotomy or lumbo-abdominal approach | T_MCOaaA/CDC_ACT |
| Viscera | CCAM (procedures) | JCPA002 | Freeing the ureter without intraperitonealization, by direct approach | T_MCOaaA/CDC_ACT |
| Viscera | CCAM (procedures) | JDLD001 | Placement of a urethrovesical catheter [Indwelling bladder catheterization]. | T_MCOaaA/CDC_ACT |
| Viscera | CCAM (procedures) | JJFA004 | Salpingoovariectomy [Annexectomy], by laparotomy | T_MCOaaA/CDC_ACT |
| Viscera | CCAM (procedures) | JKFA015 | Total hysterectomy, by laparotomy | T_MCOaaA/CDC_ACT |
| Viscera | CCAM (procedures) | JKFA027 | Total colpohysterectomy extended to parameters, by laparotomy | T_MCOaaA/CDC_ACT |
| Viscera | CCAM (procedures) | JKFA028 | Total hysterectomy with unilateral or bilateral adnexectomy, by laparotomy | T_MCOaaA/CDC_ACT |
| Viscera | CCAM (procedures) | JKQX005 | Pathological examination for carcinological purposes of hysterectomy specimen, with adnexectomy | T_MCOaaA/CDC_ACT |

- R: this variable can be found in the SNDS via CCAM procedures. The codes retained are chosen among the group « ADC » (which stands for "*actes de chirurgie*" meaning surgery procedures) and whose description contains the French following words: "*exérèse*", "*résection*" or "*ectomie*" .
- M: this variable can be found in the SNDS via CCAM procedures.
- O: this variable can be found in the SNDS via CCAM procedures.

*Section C: Rules for semantic alignment of checking variables in both databases*

**Checking variables in the source database (NETSARC)**

• d: Use of the information 'Type of tumor' (ID=1091 in NETSARC), 'Site of tumor' (ID=2 in NETSARC) and 'Date of surgery' (ID=1079 in NETSARC)
Value = The combination of 'Site of tumor + Type of tumor' allows a mapping with the specific ICD-10 table and produces one or several ICD-10 codes which are then associated with the date of surgery. This variable is only available in the clinical database.

• c: Use of the information 'Decision 1' (ID=1119 in NETSARC) , 'Decision 2' (ID=1120 in NETSARC), 'Decision 3' (ID=1121 in NETSARC) and 'Date of RCP' (ID=1105 in NETSARC)
Value = 1 If one of the items 'Decision n' is in the following list ('Indication for chemotherapy', 'Indication for neoadjuvant chemotherapy', 'Indication for adjuvant chemotherapy', 'Indication for metastatic chemotherapy', 'Continuation of chemotherapy protocol', 'Modification of chemotherapy protocol') and then associated with the date of RCP. This variable is only available in the clinical database.

• r: Use of the information 'Decision 1' (ID=1119 in NETSARC) , 'Decision 2' (ID=1120 in NETSARC), 'Decision 3' (ID=1121 in NETSARC) and 'Date of RCP' (ID=1105 in NETSARC)
Value = 1 If one of the items 'Decision n' is in the following list ('Indication for radiotherapy', 'Indication for neoadjuvant radiotherapy', 'Indication for adjuvant radiotherapy', 'Indication for exclusive or palliative radiotherapy') and then associated with the date of RCP. This variable is only available in the clinical database.

• e: Use of the information 'Decision 1' (ID=1119 in NETSARC) , 'Decision 2' (ID=1120 in NETSARC), 'Decision 3' (ID=1121 in NETSARC) and 'Date of RCP' (ID=1105 in NETSARC)
Value = 1 If one of the items 'Decision n' is ('Indication for re-excision') and then associated with the date of RCP. This variable is only available in the clinical database.

**Checking variables in the target database (SNDS)**

• d: the annual tables T_MCOaaB and T_MCOaaC contain details about the hospital stays of the patient. We use the item T_MCOaa_B.DGN_PAL which is the main diagnosis (ICD-10 code) which is associated with the temporal window between T_MCOaaC.ENT_DAT (date of entry of the stay) and T_MCOaa_C.SOR_DAT (end date of stay). In order to detect any events occurring after the date of the MTB which could be linked to the decisions taken the annual tables T_MCOaaB and T_MCOaaC are used.

• c: for the chemotherapy events we used the following pattern: search a value of T_MCOaa_B.DGN_PAL = 'Z511' which stands for "Encounter for antineoplastic chemotherapy" combined with a value of T_MCOaa_B.DGN_REL that belongs to the site/type of tumour → ICD-10 mapping table for a given stay. If OK then the chemotherapy is associated with the temporal window between T_MCOaaC.ENT_DAT (date of entry of the stay) and T_MCOaa_C.SOR_DAT (end date of stay)

• r: for the radiotherapy events we used the following pattern: search a value of T_MCOaa_B.DGN_PAL = 'Z5100' which stands for "Encounter for radiation preparation" or T_MCOaa_B.DGN_PAL = 'Z5101' which stands for "Encounter for radiation" combined with a value of T_MCOaa_B.DGN_REL that belongs to the site/type of tumor → ICD-10 mapping table for a given stay. If OK then the radiotherapy is associated with the temporal window between T_MCOaaC.ENT_DAT (date of entry of the stay) and T_MCOaa_C.SOR_DAT (end date of stay).

Table 2. Checking variables used in step 2 of the pairing process

| Letter | Checking variable name | Margin of error on date (± days) | Origin of the variable |
|---|---|---|---|
| **d** | Main ICD-10 code of the surgery stay | 30 | NETSARC & RREPS |
| **c** | Chemotherapy planned in MDTM | Within 60 days after MDTM date | NETSARC |
| **r** | Radiotherapy planned in MDTM | Within 60 days after MDTM date | NETSARC |
| **e** | Tumor re-excision planned in MDTM | Within 60 days after MDTM date | NETSARC |

MDTM stands for Multidisciplinary team meeting

*Section D: Exhaustive crude results of the pairing algorithm*

| Signature | Number of patients | Distribution of the robustness variable R (min=0 /max=6) | Median | Range | Pct for R=0 | Pct for R=1 | Pct for R>1 |
|---|---|---|---|---|---|---|---|
| Ss.LlC.R.. | 4646 | 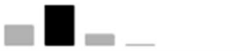 | 1 | 0 - 3 | 27,87 % | 57,58 % | 14,55 % |
| Ss.Ll..R.. | 3695 | 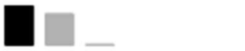 | 0 | 0 - 2 | 55,45 % | 43,95 % | 0,60 % |
| Ss.LlC.... | 2882 | 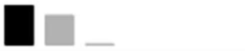 | 0 | 0 - 2 | 57,01 % | 42,40 % | 0,59 % |
| Ss.Ll...M. | 1272 | 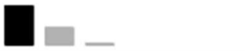 | 0 | 0 - 2 | 68,00 % | 29,87 % | 2,12 % |
| Ss.LlCFRM. | 1083 | 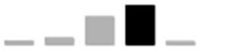 | 3 | 0 - 4 | 4,43 % | 8,03 % | 87,53 % |
| Ss.Ll....O | 980 | 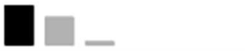 | 0 | 0 - 2 | 56,33 % | 39,49 % | 4,18 % |
| SsDLlC.R.. | 925 | 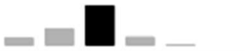 | 2 | 0 - 4 | 9,73 % | 23,14 % | 67,14 % |
| Ss..l..R.. | 903 | 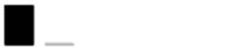 | 0 | 0 - 1 | 98,45 % | 1,55 % | 0,00 % |
| Ss.LlC.RM. | 880 | 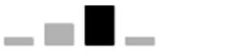 | 2 | 0 - 3 | 8,86 % | 27,95 % | 63,18 % |
| Ss.LlC...O | 673 | 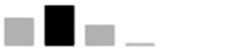 | 1 | 0 - 3 | 30,91 % | 45,62 % | 23,48 % |
| Ss.LlC..M. | 616 | 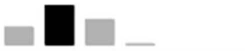 | 1 | 0 - 3 | 20,94 % | 47,89 % | 31,17 % |
| Ss.LlCF... | 608 | 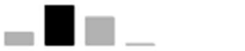 | 1 | 0 - 3 | 15,30 % | 49,51 % | 35,20 % |
| Ss.LlCF.M. | 439 | 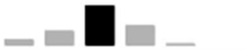 | 2 | 0 - 4 | 6,38 % | 17,54 % | 76,08 % |
| SsDLlC.... | 406 | 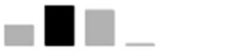 | 1 | 0 - 3 | 20,94 % | 41,63 % | 37,44 % |
| Ss.LlC.R.O | 377 | 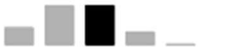 | 1 | 0 - 4 | 15,65 % | 36,07 % | 48,28 % |
| Ss.Ll..RM. | 363 | 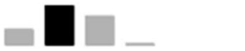 | 1 | 0 - 3 | 18,18 % | 46,56 % | 35,26 % |
| Ss.LlCFR.. | 345 | 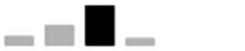 | 2 | 0 - 3 | 11,30 % | 26,67 % | 62,03 % |
| SsDLlCFRM. | 292 | 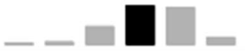 | 3 | 0 - 5 | 1,37 % | 2,40 % | 96,23 % |
| Ss..lC.R.. | 236 | 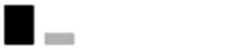 | 0 | 0 - 1 | 79,66 % | 20,34 % | 0,00 % |
| Ss.LlCFR.O | 214 | 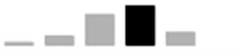 | 3 | 0 - 4 | 2,34 % | 9,35 % | 88,32 % |
| Ss..l...M. | 188 | 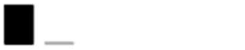 | 0 | 0 - 1 | 96,81 % | 3,19 % | 0,00 % |
| Ss.LlCF..O | 182 | 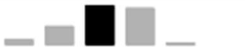 | 2 | 0 - 4 | 4,95 % | 18,13 % | 76,92 % |
| Ss.Ll..R.O | 178 | 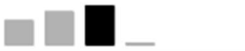 | 1 | 0 - 3 | 24,72 % | 33,71 % | 41,57 % |
| Ss..l…O | 175 | 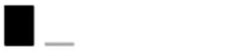 | 0 | 0 - 1 | 95,43 % | 4,57 % | 0,00 % |
| SsDLlC.RM. | 174 | 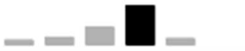 | 3 | 0 - 4 | 5,75 % | 9,77 % | 84,48 % |
| SsDLlC...O | 153 | 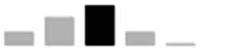 | 2 | 0 - 4 | 13,73 % | 29,41 % | 56,86 % |
| SsDLlC..M. | 145 | 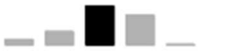 | 2 | 0 - 4 | 6,21 % | 15,17 % | 78,62 % |
| Ss...C.... | 134 | 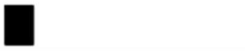 | 0 | 0 - 0 | 100,00 % | 0,00 % | 0,00 % |
| SsDLlCF.M. | 104 | 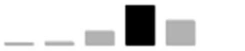 | 3 | 0 - 4 | 1,92 % | 2,88 % | 95,19 % |

| Signature | Number of patients | Distribution of the robustness variable R (min=0 /max=6) | Median | Range | Pct for R=0 | Pct for R=1 | Pct for R>1 |
|---|---|---|---|---|---|---|---|
| SsDLlCF... | 101 | | 2 | 0 - 3 | 5,94 % | 17,82 % | 76,24 % |
| SsDLlC.R.O | 100 | | 2 | 0 - 4 | 7,00 % | 14,00 % | 79,00 % |
| Ss..lC.... | 96 | | 0 | 0 - 1 | 96,88 % | 3,13 % | 0,00 % |
| SsDLlCFR.O | 90 | | 3 | 0 - 5 | 2,22 % | 7,78 % | 90,00 % |
| Ss.....R.. | 76 | | 0 | 0 - 0 | 100,00 % | 0,00 % | 0,00 % |
| SsDLlCFR.. | 66 | | 3 | 0 - 4 | 6,06 % | 7,58 % | 86,36 % |
| SsDLlCF..O | 52 | | 3 | 0 - 4 | 9,62 % | 3,85 % | 86,54 % |
| Ss......M. | 47 | | 0 | 0 - 0 | 100,00 % | 0,00 % | 0,00 % |
| Ss..lC...O | 46 | | 0 | 0 - 2 | 78,26 % | 15,22 % | 6,52 % |
| Ss.Ll..... | 46 | | 0 | 0 - 0 | 100,00 % | 0,00 % | 0,00 % |
| Ss..l..RM. | 42 | | 0 | 0 - 1 | 59,52 % | 40,48 % | 0,00 % |
| Ss.......O | 33 | | 0 | 0 - 0 | 100,00 % | 0,00 % | 0,00 % |
| Ss..lCFRM. | 32 | | 2 | 0 - 3 | 12,50 % | 12,50 % | 75,00 % |
| Ss.Ll...MO | 32 | | 1 | 0 - 2 | 34,38 % | 40,63 % | 25,00 % |
| Ss..lC.RM. | 28 | | 1 | 0 - 2 | 35,71 % | 50,00 % | 14,29 % |
| Ss..lC..M. | 23 | | 1 | 0 - 2 | 47,83 % | 47,83 % | 4,35 % |
| SsD.lC.R.. | 22 | | 1 | 0 - 2 | 45,45 % | 40,91 % | 13,64 % |
| Ss..l..R.O | 21 | | 0 | 0 - 2 | 80,95 % | 14,29 % | 4,76 % |
| Ss...CF... | 18 | | 0 | 0 - 1 | 66,67 % | 33,33 % | 0,00 % |
| Ss...C.R.. | 17 | | 0 | 0 - 0 | 100,00 % | 0,00 % | 0,00 % |
| Ss.LlC.RMO | 16 | | 2 | 1 - 4 | 0,00 % | 25,00 % | 75,00 % |
| Ss..lCF... | 15 | | 1 | 0 - 2 | 40,00 % | 53,33 % | 6,67 % |
| Ss.LlCFRMO | 14 | | 3 | 1 - 5 | 0,00 % | 7,14 % | 92,86 % |
| Ss..lC.R.O | 13 | | 1 | 0 - 2 | 38,46 % | 38,46 % | 23,08 % |
| Ss..lCF.M. | 13 | | 1 | 0 - 2 | 15,38 % | 38,46 % | 46,15 % |
| SsD..C.... | 13 | | 0 | 0 - 1 | 92,31 % | 7,69 % | 0,00 % |
| Ss.....RM. | 12 | | 0 | 0 - 1 | 91,67 % | 8,33 % | 0,00 % |
| Ss.LlC..MO | 12 | | 2 | 0 - 3 | 16,67 % | 16,67 % | 66,67 % |
| Ss..lCF..O | 11 | | 2 | 0 - 2 | 9,09 % | 18,18 % | 72,73 % |
| Ss..lCFR.. | 11 | | 1 | 0 - 2 | 18,18 % | 72,73 % | 9,09 % |

| Signature | Number of patients | Distribution of the robustness variable R (min=0 /max=6) | Median | Range | Pct for R=0 | Pct for R=1 | Pct for R>1 |
|---|---|---|---|---|---|---|---|
| Ss...CFR.. | 9 | 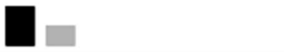 | 0 | 0 - 1 | 66,67 % | 33,33 % | 0,00 % |
| SsD.lC.... | 9 | 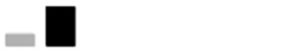 | 1 | 0 - 1 | 22,22 % | 77,78 % | 0,00 % |
| SsDLl..... | 9 | 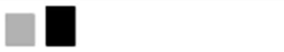 | 1 | 0 - 1 | 44,44 % | 55,56 % | 0,00 % |
| SsDLl..RM. | 9 | 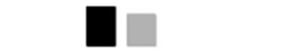 | 2 | 2 - 3 | 0,00 % | 0,00 % | 100,00 % |
| Ss..lCFR.O | 8 | 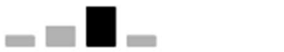 | 2 | 0 - 3 | 12,50 % | 25,00 % | 62,50 % |
| SsDLlC.RMO | 7 | 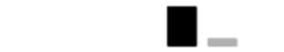 | 4 | 4 - 5 | 0,00 % | 0,00 % | 100,00 % |
| SsDLlCFRMO | 7 | 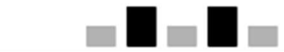 | 4 | 2 - 6 | 0,00 % | 0,00 % | 100,00 % |
| Ss.....R.O | 5 | 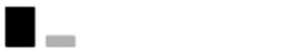 | 0 | 0 - 1 | 80,00 % | 20,00 % | 0,00 % |
| Ss...CF..O | 5 | 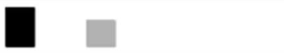 | 0 | 0 - 2 | 60,00 % | 0,00 % | 40,00 % |
| SsD.LC...O | 5 | 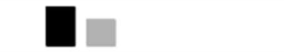 | 1 | 1 - 2 | 0,00 % | 60,00 % | 40,00 % |
| SsDLl..R.. | 5 | 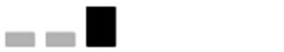 | 2 | 0 - 2 | 20,00 % | 20,00 % | 60,00 % |
| SsDLl....O | 5 | 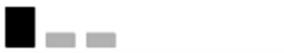 | 0 | 0 - 2 | 60,00 % | 20,00 % | 20,00 % |
| Ss...CFR.O | 4 | 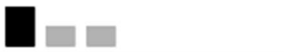 | 0 | 0 - 2 | 50,00 % | 25,00 % | 25,00 % |
| Ss.LlCF.MO | 4 | 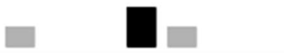 | 3 | 0 - 4 | 25,00 % | 0,00 % | 75,00 % |
| SsDLlC..MO | 4 | 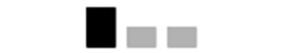 | 2 | 2 - 4 | 0,00 % | 0,00 % | 100,00 % |
| Ss...C...O | 3 | 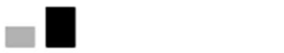 | 1 | 0 - 1 | 33,33 % | 66,67 % | 0,00 % |
| Ss...C..M. | 3 | 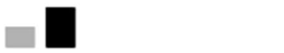 | 1 | 0 - 1 | 33,33 % | 66,67 % | 0,00 % |
| Ss.Ll..RMO | 3 | 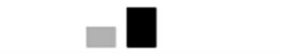 | 3 | 2 - 3 | 0,00 % | 0,00 % | 100,00 % |
| SsD..CF... | 3 | 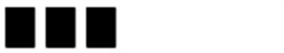 | 1 | 0 - 2 | 33,33 % | 33,33 % | 33,33 % |
| SsD..CFR.. | 3 | 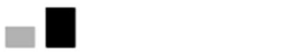 | 1 | 0 - 1 | 33,33 % | 66,67 % | 0,00 % |
| SsD.lC.RM. | 3 | 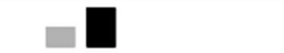 | 2 | 1 - 2 | 0,00 % | 33,33 % | 66,67 % |
| SsD.lCFR.O | 3 | 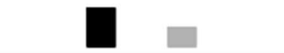 | 2 | 2 - 4 | 0,00 % | 0,00 % | 100,00 % |
| SsD.lCFRM. | 3 | 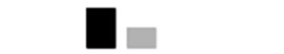 | 2 | 2 - 3 | 0,00 % | 0,00 % | 100,00 % |
| SsDLl...M. | 3 | 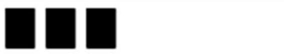 | 1 | 0 - 2 | 33,33 % | 33,33 % | 33,33 % |
| SsDLlCF.MO | 3 | 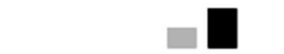 | 5 | 4 - 5 | 0,00 % | 0,00 % | 100,00 % |
| Ss...C.R.O | 2 | 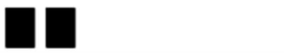 | 0 | 0 - 1 | 50,00 % | 50,00 % | 0,00 % |
| Ss...CF.M. | 2 | 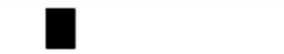 | 1 | 1 - 1 | 0,00 % | 100,00 % | 0,00 % |
| Ss...CFRM. | 2 | 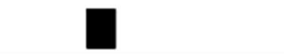 | 2 | 2 - 2 | 0,00 % | 0,00 % | 100,00 % |
| Ss..l...MO | 2 | 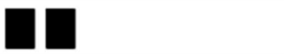 | 0 | 0 - 1 | 50,00 % | 50,00 % | 0,00 % |
| Ss..lC..MO | 2 | 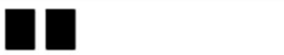 | 0 | 0 - 1 | 50,00 % | 50,00 % | 0,00 % |

| Signature | Number of patients | Distribution of the robustness variable R (min=0 /max=6) | Median | Range | Pct for R=0 | Pct for R=1 | Pct for R>1 |
|---|---|---|---|---|---|---|---|
| SsD..CF..O | 2 | | 1 | 1 - 2 | 0,00 % | 50,00 % | 50,00 % |
| SsD.lC..M. | 2 | | 1 | 1 - 2 | 0,00 % | 50,00 % | 50,00 % |
| SsD.lCFR.. | 2 | | 0 | 0 - 3 | 50,00 % | 0,00 % | 50,00 % |
| Ss...C.RM. | 1 | | 1 | 1 - 1 | 0,00 % | 100,00 % | 0,00 % |
| Ss..l..... | 1 | | 0 | 0 - 0 | 100,00 % | 0,00 % | 0,00 % |
| Ss..lCF.MO | 1 | | 2 | 2 - 2 | 0,00 % | 0,00 % | 100,00 % |
| Ss..lCFRMO | 1 | | 3 | 3 - 3 | 0,00 % | 0,00 % | 100,00 % |
| SsD..C..M. | 1 | | 2 | 2 - 2 | 0,00 % | 0,00 % | 100,00 % |
| SsD..C.R.O | 1 | | 0 | 0 - 0 | 100,00 % | 0,00 % | 0,00 % |
| SsD..C.RM. | 1 | | 2 | 2 - 2 | 0,00 % | 0,00 % | 100,00 % |
| SsD.l..RM. | 1 | | 2 | 2 - 2 | 0,00 % | 0,00 % | 100,00 % |
| SsD.lC.R.O | 1 | | 2 | 2 - 2 | 0,00 % | 0,00 % | 100,00 % |
| SsD.lCF... | 1 | | 2 | 2 - 2 | 0,00 % | 0,00 % | 100,00 % |
| SsD.lCF.M. | 1 | | 2 | 2 - 2 | 0,00 % | 0,00 % | 100,00 % |
| SsDLl..R.O | 1 | | 2 | 2 - 2 | 0,00 % | 0,00 % | 100,00 % |

All 104 groups have the two first chaining (S for Sex code + month and year of birth or s for sex code + year of birth) always present in the signatures. This means that their combination is essential in the chaining process. It can also be noted that the location of the patient (L for residency town code or l for residency department code) is present in the 27 first signatures (cumulating 93,85 % of the total unique pairs) which means that this information is also very contributive to the chaining process.
